## Supplement for "Epidemiological drivers of transmissibility and severity of SARS-CoV-2 in England"

CONTENTS

1 Model Overview 3
1.1 Model description 3
1.2 Reproduction number 4
1.3 Fitting to data 5

2 Variants of Concern 7
2.1 Seeding the epidemic with variants 7
2.2 Serial interval of variants 7
2.3 Vaccine effectiveness by strain 8
2.4 Strain relative severity 8
2.5 Cross-immunity given previous infection 8

3 Vaccination 9
3.1 Vaccine effectiveness 9
3.2 Waning of vaccine-induced immunity 15
3.3 Conditional dependencies of vaccine-immunity 15
3.4 Vaccine roll-out 15

4 Model Parameterisation and Fitting 17
4.1 Model compartments and parameters 17
4.2 Modelling of variants 18
4.3 Parallel flows 18
4.4 Equations 20
4.4.1 Force of infection 20
4.4.2 Seeding of variants 21
4.4.3 Pathway probabilities and rates 22
4.4.4 Time-varying severity parameters 25
4.4.5 Compartmental model equations 25
4.5 Observation process 34
4.5.1 Notation for distributions used in this section 34
4.5.2 Hospital admissions and new diagnoses in hospital 35
4.5.3 Hospital bed occupancy by confirmed COVID-19 cases 35
4.5.4 Hospital and community COVID-19 deaths 36
4.5.5 Serosurveys 36
4.5.6 PCR testing 37
4.5.7 Variant and Mutation data 38
4.5.8 Full likelihood 38
4.6 Reproduction number 39
4.7 Basic and effective severity 39
4.8 Effective population-level immunity against infection 41
4.9 Fixed parameters 41
4.10 Prior distributions 42
4.11 Running the model 46

5 Sensitivity Analyses 47
5.1 Vaccine efficacy sensitivity analysis 48
5.2 Cross-immunity sensitivity analysis 50
5.3 Other sensitivity analysis 51

6 Supplementary Results 53

Symbol Glossary 84

List of Figures 86

List of Tables 87

### 1 Model Overview

In this section we provide a brief description to our model along with key definitions. Full details about the fitting procedure, parameter assumptions, and model equations are provided in Section 4.

The study period considered is from the 16th of March 2020 to the 24th of February 2022. This time frame considers the roll-out of the initial vaccination programme and of the first boosters programme in England, as well as the sequential emergence and establishment of the variants of concern (VOCs), Alpha, Delta and Omicron BA.1.

#### 1.1 Model description

We adapt a previously described discrete-time (1/4 day time step) stochastic compartmental model of SARS-CoV-2 transmission (Figure S1) [1, 2, 3]. The model is an extended SEIR-type model, stratified into 17 age groups: 16 five-year age bands (0-4, 5-9, ..., 75-79) plus a group of 80+ year-olds. Mixing between age groups is informed by survey data [4].

Upon infection with SARS-CoV-2, individuals enter an exposed compartment, before becoming infectious. A proportion of infectious individuals are assumed to develop symptoms, while the rest remain asymptomatic. All asymptomatic cases and a fraction of symptomatic cases recover naturally, while the rest of the symptomatic cases develop severe disease requiring hospitalisation. Of these, a proportion die outside hospital, while the remainder are admitted to hospital.

An important feature of our model is we explicitly model hospital flows (Figure S1B), which has been previously used to track and inform the epidemic response in England in real-time [5, 6, 7, 8]. Individuals entering hospital flows are triaged for intensive care unit (ICU) admission or remaining in general beds throughout from where they can either die or recover and be discharged. Those admitted to ICU, can either die in ICU or be transferred for stepdown care in general wards, where they can either die or recover and be discharged. Hospitalised cases are either confirmed as SARS-CoV-2 cases upon admission or may be tested and confirmed later during their stay.

Model compartments were expanded to account for six vaccination strata (see Section 3 and Table S3C). These strata describe the recommended primary (two-dose) and boosters regimen common to the three vaccines predominantly used in England during the study period: Oxford-AstraZeneca ChAdOx1 nCoV-19 (AZD1222) [9], Pfizer-BioNTech COVID-19 Vaccine BNT162b2 [10], and Moderna mRNA-1273 [11] (henceforth referred to as AZ, PF, and Mod, respectively). Vaccine strata further capture delays between receiving a dose and the onset of dose-specific vaccine effectiveness (VE), as well as waning of vaccine-induced immunity following second and booster doses (see Section 3.2).

The model was further extended to account for infection flows with two actively co-circulating variants (see Section 2 and Figure S1D). In the context of this paper, we explicitly consider the Wildtype (Wuhan-like), Alpha (B.1.1.7), Delta (B.1.617.2) and Omicron BA.1 (B.1.1.529) variants. For each subsequent VOC emergence and replacement period, we fit a two-variant model, with the emerging VOC seeded at a region-specific date determined by the model fit. We model waning of infection-induced immunity assuming protection against reinfection with the same variant for an exponentially distributed duration with mean 3 years [12]. After waning, individuals are assumed to move back to the susceptible compartment. Further, we model asymmetrical cross-immunity to a new SARS-CoV-2 variant (Section 2.5) for individuals who have recovered from previous variants.

Please note that throughout our main manuscript and this supplement we use the term 'susceptible' only to refer to individuals in compartment '$S$', whereas 'uninfected' refers to those in either the '$S$' or '$R$' (recovered) compartments.

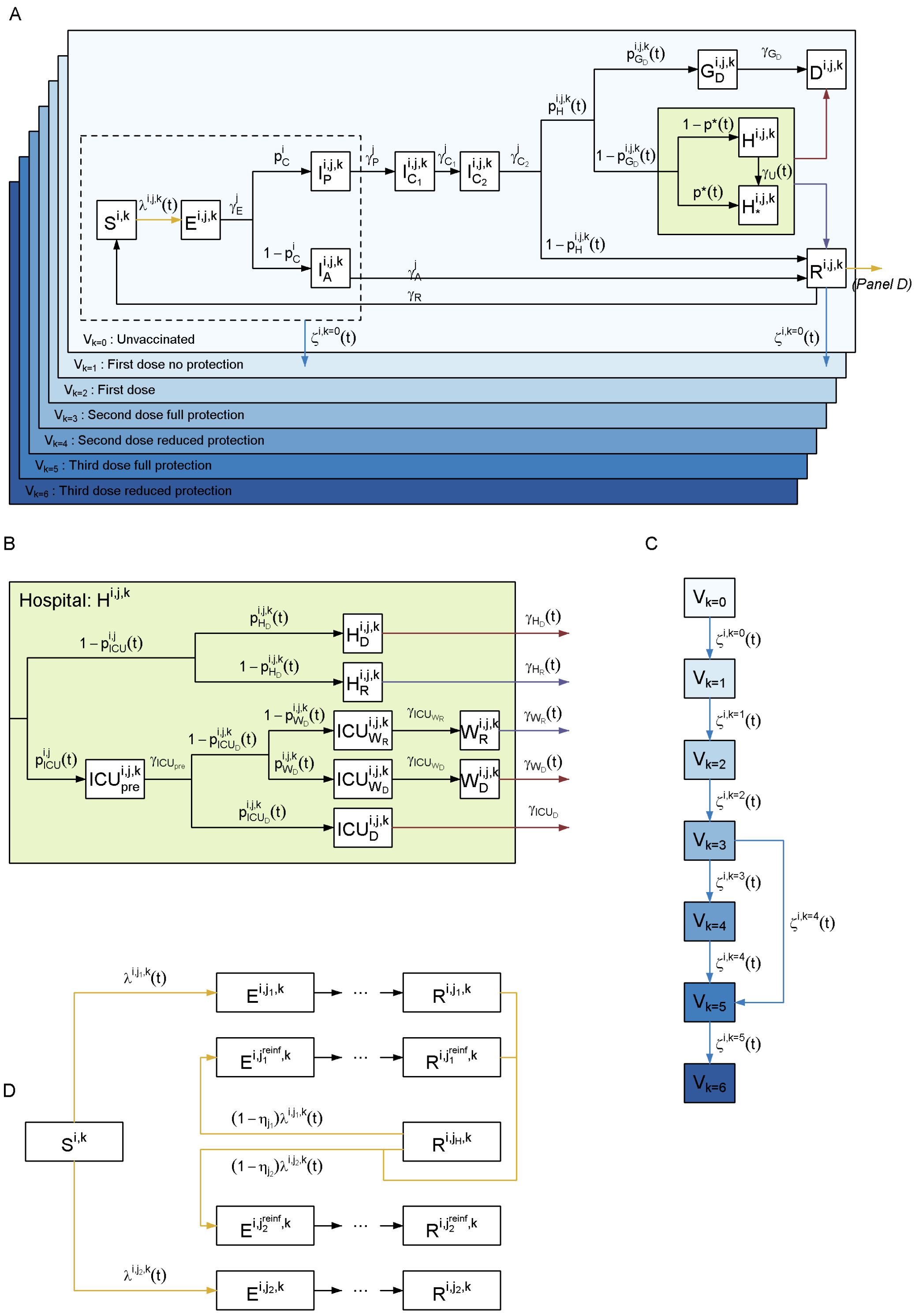

**Figure S1:** Model structure flow diagram with rates of transition between states. (A) Extended SEIR transmission model flow diagram overview. (B) Hospital flow diagram. (C) Vaccination flow diagram. (D) Multi-variant flow diagram. Model state variables and parameters are presented in detail throughout this supplement; in particular, see section 4.

### 1.2 Reproduction number

We use two definitions of the reproduction number throughout. We denote $R_t^j$ as the reproduction number for variant $j$ ($j = Wildtype, Alpha, Delta, Omicron$) in the absence of immunity at time $t$, which varies only in response to fitted $\beta$

changepoints (e.g. changes in non-pharmaceutical intervention (NPI) policies). This is defined as the average number of secondary infections that an individual infected at time $t$ with variant $j$ would generate in an entirely susceptible and unvaccinated population. In contrast, the effective reproduction number, $R_t^{j,eff}$, for variant $j$ at time $t$ is the number of secondary infections in the actual population, further accounting for immunity (infection- and vaccine-derived) and NPI policies present at that time in the population. Hence, by definition, $R_t^{j,eff} \leq R_t^j$.

#### 1.3 Fitting to data

The model is fitted to multiple data streams from each National Health Service (NHS) region in England, as summarised in Table S1, using a particle-Markov chain Monte Carlo (pMCMC) algorithm. Where age-bands are specified, we fitted to data by age. Pillar 2 testing, hospital admissions and deaths (community and hospital) were pre-processed from linelist (patient-level) data to aggregated timeseries.

For the case of hospital admissions, we counted patients on their date of first entering hospital if coming from the community within 14 days of a positive PCR test, or the date of a positive PCR test within hospital if already in hospital when diagnosed with COVID-19, and only if their hospital stay was longer than 24 hours.

Please see details of the technical procedure used to run the model for the present analysis in Section 4.11.

| Data | Description | Source | Reference |
|---|---|---|---|
| Hospital deaths by age | Daily number of deaths with COVID-19 mentioned as a cause on the death certificate and "hospital" as the place of death, in age bands 0-49, 50-54, 55-59, 60-64, 65-69, 70-74, 75-79 and 80+ | ONS | These data underlie the Gov.uk dashboard data [13] |
| Community deaths | Daily number of deaths with COVID-19 mentioned as a cause on the death certificate and any place of death that is not "hospital", in age bands 0-49, 50-54, 55-59, 60-64, 65-69, 70-74, 75-79 and 80+ | ONS | These data underlie the Gov.uk dashboard data [13] |
| ICU occupancy | Daily number of confirmed COVID-19 patients in ICU | Gov.uk Dashboard | [13] |
| General bed occupancy | Daily number of confirmed COVID-19 patients in non-ICU beds | Gov.uk Dashboard | [13] |
| Admissions | Daily number of confirmed COVID-19 patients admitted to hospital from SUS/ECDS linelist in age bands 0-9, 10-19, 20-29, 30-39, 40-49, 50-59, 60-69, 70-79 and 80+ | PHE | These data underlie the Gov.uk dashboard data [13] |
| Pillar 2 testing | Daily number of positive (cases) and negative PCR test results in age bands 15-24, 25-49, 50-64, 65-79 and 80+ | PHE | These data underlie the Gov.uk dashboard data [13] |
| REACT-1 testing | Real-time Assessment of Community Transmission (REACT) daily number of positive and negative PCR test results in age bands 5-24, 25-34, 35-44, 45-54, 55-64 and 65+ | REACT | [14] |
| ONS testing | Infection survey by the Office for National Statistics by NHS England region, reporting weekly number of positive and negative PCR test results | ONS | [15] |
| Serology | Serology survey conducted on blood donors aged 15-65. Results from the EuroImmun and Roche N assays are used, fitting to each assay separately. EuroImmun results are only used up to (and including) 14th January 2020 | PHE | These data are collected as part of [16]. |
| Vaccinations by age | Daily number of first and second-vaccine doses - reported in 5-year age groups | PHE | [13] |
| Variant and Mutation | Daily number of variant tests of symptomatic individuals with COVID-19 confirmed by PCR that are identified as "Unclassified" (assumed to be Wildtype), Alpha, Delta or Omicron BA.1 | VAM | These data underlie the Gov.uk genomic surveillance reports [17] |

**Table S1:** Data sources and definitions. All data are reported by NHS region or processed to match these regions

### 2 Variants of Concern

For model fitting, we switch from a one-variant to a two-variant model on 17th September 2020 to capture the emergence and spread of the first variant of concern to emerge in England, the Alpha variant. Subsequently, we rotate the two variant model, to capture the emergence and spread of the Delta and Omicron variants. Key differences between the modelled variants are summarised in the next subsections, these are:

1. Seeding dates.
2. Differences in transmissibility between variants
3. Differences in vaccine effectiveness for each variant for various endpoints.
4. Differences in severity between variants.
5. Asymmetric cross immunity between the variants.

#### 2.1 Seeding the epidemic with variants

We fit seeding dates for each of the variants: $t_{Wildtype}$ (which corresponds to the start date of the regional epidemic), $t_{Alpha}$, $t_{Delta}$ and $t_{Omicron}$ (see Table S12). The seeding is detailed more in section 4.4.2.

#### 2.2 Serial interval of variants

We fit a region-specific transmission advantage, $\sigma$, of the emerging variant compared to the immediately preceding one (e.g. Omicron BA.1 compared to Delta). We use a uniform prior between 0 and 3 (Table S12).

As previously described in publications from our group [1, 2, 3], we derived the expected duration in compartments with the formula for mean backwards serial interval [18]:

$$\mathbb{E}[\tau_E] + \frac{\mathbb{E}[\tau_{I_P}]^2 + \mathbb{E}[\tau_{I_P}]\mathbb{E}\left[\tau_{I_{C_1}}\right] + \mathbb{E}\left[\tau_{I_{C_1}}\right]^2}{\mathbb{E}[\tau_{I_P}] + \mathbb{E}\left[\tau_{I_{C_1}}\right]}. \tag{1}$$

Recent literature provides strong evidence that the relative rate of replacement of a new variant is sensitive to assumptions of the mean serial interval ([19]). Across multiple settings it has been estimated that the mean serial interval of SARS-CoV-2 has shortened with the emergence of variants of concern [20, 21, 22, 23]. We therefore explicitly modelled differences in the serial interval, by proportionally reducing the baseline serial interval of variants of concern compared to the Wildtype variant (table S2). We achieve this for each variant by applying the reduction to the mean durations of the $E$, $I_P$, $I_{C_1}$ and $I_A$ compartments. However we assume the same mean time ($\mathbb{E}\left[\tau_{I_{C_1}}\right] + \mathbb{E}\left[\tau_{I_{C_2}}\right] = 4$) from onset sypmtom to (possible) hospitalisation across variants. Thus $\mathbb{E}\left[\tau_{I_{C_2}}\right]$ also varies across variants.

| Variant | Mean SI (days) | % reduction | Source | Compartment | Erlang duration parameters $k_X$ | $\gamma_X^j$ | Mean |
|---|---|---|---|---|---|---|---|
| Wildtype | 5.2 | - | [2, 3] | $E$ | 2 | 0.87 | 2.31 |
| | | | | $I_A$ | 1 | 0.35 | 2.88 |
| | | | | $I_P$ | 1 | 0.60 | 1.68 |
| | | | | $I_{C_1}$ | 1 | 0.47 | 2.14 |
| | | | | $I_{C_2}$ | 1 | | 1.86 |
| Alpha | 4.9 | 6% | [21] | $E$ | 2 | 0.92 | 2.17 |
| | | | | $I_A$ | 1 | 0.37 | 2.71 |
| | | | | $I_P$ | 1 | 0.63 | 1.58 |
| | | | | $I_{C_1}$ | 1 | 0.50 | 2.01 |
| | | | | $I_{C_2}$ | 1 | 0.50 | 1.99 |
| Delta | 4.5 | 13% | [21] | $E$ | 2 | 0.99 | 2.01 |
| | | | | $I_A$ | 1 | 0.40 | 2.51 |
| | | | | $I_P$ | 1 | 0.68 | 1.46 |
| | | | | $I_{C_1}$ | 1 | 0.54 | 1.86 |
| | | | | $I_{C_2}$ | 1 | 0.47 | 2.14 |
| Omicron | 3.9 | 25% | [21] | $E$ | 2 | 1.15 | 1.73 |
| | | | | $I_A$ | 1 | 0.46 | 2.16 |
| | | | | $I_P$ | 1 | 0.79 | 1.26 |
| | | | | $I_{C_1}$ | 1 | 0.62 | 1.61 |
| | | | | $I_{C_2}$ | 1 | 0.42 | 2.39 |

**Table S2:** Serial interval assumptions for the different variants

#### 2.3 Vaccine effectiveness by strain

We assume variations in vaccine effectiveness across modelled strains. Estimates of vaccine effectiveness were fixed, informed by relevant literature from England and/or the UK (see table S4). Both the variant-specific estimated transmission advantage, $\sigma$, and vaccine effectiveness were captured in the force of infection (see Section 4.4 Section 3 and Table S4).

#### 2.4 Strain relative severity

To account for potential difference in disease severity of emerging variants of concern, we fitted multipliers for the probability of of hospitalisation ($\pi_H^j$), admission to ICU ($\pi_{ICU}^j$) and death ($\pi_D^j$), where $j$ stands for the specific strain. These probabilities where conditional upon transition to the immediately preceding severity pathway compartment (e.g. $\pi_H^j$ upon infection with strain $j$) and relative to the immediately previous strain (e.g. $\pi_H^{Omicron}$ for the probability of hospitalisation upon infection with Omicron BA.1, relative to Delta). For further details of severity pathway probabilities, see section 4.4.3.

#### 2.5 Cross-immunity given previous infection

The level of cross-protection from prior infection is difficult to quantify. In-vitro antibody neutralisation studies have reported that emerging variants are neutralised to a lesser extent by antibodies from previous infections with preceding variants [24]. We model asymmetric cross immunity across variants, assuming that infection with with emerging strain confers perfect immunity against infection with the strain being replaced. We further assume that infection with prior infection is only partially protective against the new strain (see Table S11).

For reinfections (e.g. newly infected by Omicron following prior infection with an earlier variant), we assume that, if the second infection is symptomatic, the probability of hospitalisation is reduced compared to individuals with no prior infection history. In turn, we consider that if the infection leads to hospitalisation, the probability of death is also comparatively reduced.

The probabilities of infection, hospitalisation or death by either strain "reset" to baseline assumptions once an individual's infection-induced protection wanes and they re-enter the susceptible compartment, $S^{i,k}$. We assume an exponential distribution for the time to natural immunity waning, with a mean of 3 years.

### 3 Vaccination

We modelled vaccination considering the AZ, PF and Model vaccines, all three approved for use in England by the Medicines and Healthcare Products Regulatory Agency [9, 10, 11]. The model time horizon considered was from March 16, 2020 to February 24, 2022, before the rollout of fourth doses (second "boosters") or the use of current multi-valent vaccines. As such, neither of these vaccines were considered in this study. We thus explicitly modelled six distinct vaccination strata ($V_k$, for $k \in \{1,2,3,...,6\}$) representing the stages of VE detailed in Table S3 and illustrated in Figure S2.

| Vaccination stratum | Number of doses | Vaccine effectiveness | Description | References |
|---|---|---|---|---|
| $V_0$ | 0 | None | Non-vaccinated individuals. | Section 3.4 |
| $V_1$ | 1 | None | Individuals who have had their first-dose but are in a delay period of average 3 weeks until effectiveness kicks in; transition from $V_1$ is randomly drawn from an exponential distribution with mean waning time of 24 weeks. | Section 3.4 |
| $V_2$ | 1 | First dose effectiveness | Individuals with first-dose VE 3 weeks from date of first vaccination. | [25, 26] |
| $V_3$ | 2 | Full second dose effectiveness | Individuals are fully protected 1 week from date of second vaccination. | [27, 25, 26] |
| $V_4$ | 2 | Reduced second dose effectiveness | Individuals with reduced second dose vaccine protection; transition from $V_3$ is randomly drawn from an exponential distribution with mean waning time of 24 weeks. | Section 3.2 |
| $V_5$ | 3 | Booster dose effectiveness | Individuals are fully protected 1 week from date of third ("booster") vaccination. | [27] |
| $V_6$ | 3 | Reduced "booster" dose effectiveness | Individuals with reduced "booster" dose vaccine protection; transition from $V_5$ is randomly drawn from an exponential distribution with mean waning time of 24 weeks. | Section 3.2 |

**Table S3:** Vaccination strata considered for individuals, corresponding schedule, and vaccine effectiveness at each stage.

Individuals in our model move out of an unvaccinated ($V_0$) stratum at a rate determined by vaccine roll-out data and the prioritisation strategy adopted by the UK government (Section 3.4). We only allow vaccination of individuals who are not symptomatic and not hospitalised, i.e. only individuals in the following compartments can be vaccinated: susceptible ($S$), exposed ($E$), infected asymptomatic ($I_A$), infected pre-symptomatic ($I_P$) or recovered ($R$). Whilst individuals in other compartments are also stratified by vaccination strata, we do not allow movement between vaccine strata ($V_k$) for them.

As previously described for our model [2, 3], in addition to the exponentially-distributed delay of mean 21 days between the first dose and onset of VE, we assume a fixed 7-day delay between second and "booster" doses and onset of their VE. These assumptions were based on studies of immunogenicity after SARS-CoV-2 vaccination [25, 26]. This is illustrated in Figure S2.

While we generally assume that individuals can only move between successive vaccine strata, we assume that individuals in $V_3$ (full second dose effectiveness) can receive their booster dose and move to $V_5$ before their effectiveness has waned, thereby skipping $V_4$. The pool of individuals eligible for a booster dose is therefore those in $V_3$ and $V_4$ (with no prioritisation of one over the other).

#### 3.1 Vaccine effectiveness

The assumed values for vaccine effectiveness (VE) are derived from both vaccine efficacy measured in clinical trials and vaccine effectiveness studies (Table S4). Where possible, data from the UK have been used and represent effectiveness of dosing schedules with an 12 week gap between doses. We assumed that there are no significant differences in vaccine effectiveness by age, sex, or underlying health conditions [28, 29]. We assume VE against Wildtype is the same as against Alpha. As previously describe for our model [2, 3], we assume that vaccine protection against symptomatic disease

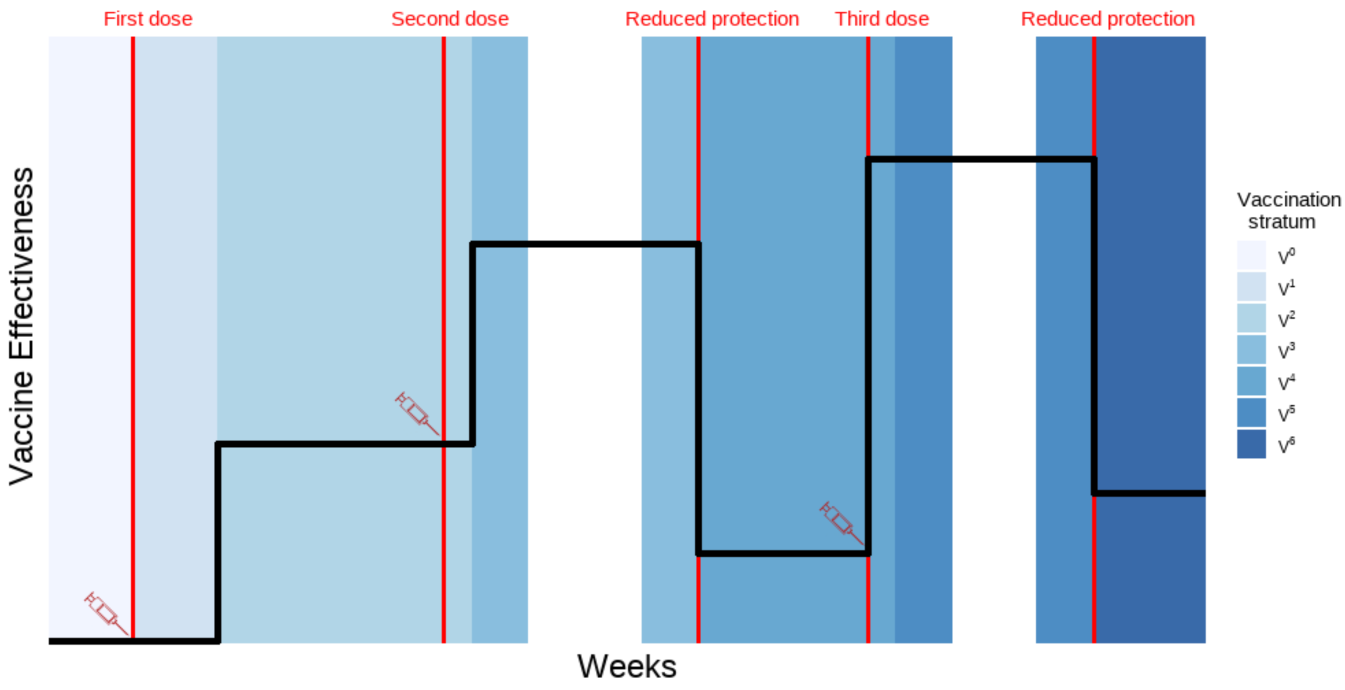

**Figure S2:** Vaccination strata duration and associated illustrative vaccine effectiveness. Red lines depict points at which a vaccine dose is administered. We assume an average 24-weeks to waning. Y-axis illustrates changing vaccine effectiveness. Vaccination strata are defined in Table S3.

also provides a similar level of protection against infection and that, in those individuals who do become infected after vaccination, onward transmission is also reduced [30].

| | | Alpha | | Delta | | Omicron | | Informed by |
|---|---|---|---|---|---|---|---|---|
| End point | Dose | AZ | PF/Mod | AZ | PF/Mod | AZ | PF/Mod | |
| Death | 1 | 88% | 89% | 87% | 89% | 53% | 53% | [31, 32] |
| | 2 (Full protection) | 99% | 99% | 99% | 99% | 97% | 97% | [31, 32] |
| | 2 (Waned protection) | 83% | 90% | 82% | 90% | 56% | 56% | [33, 34, 35] |
| | 3 (Full protection) | 99% | 99% | 99% | 99% | 96% | 96% | Assume same as vs. severe disease. |
| | 3 (Waned protection) | | | | | 62% | 62% | Assume same as vs. severe disease. |
| Severe disease | 1 | 81% | 89% | 81% | 89% | 53% | 53% | [36, 37, 38] |
| | 2 (Full protection) | 99% | 99% | 99% | 99% | 97% | 97% | [39, 40] |
| | 2 (Waned protection) | 77% | 90% | 77% | 90% | 56% | 56% | [33, 34, 35] |
| | 3 (Full protection) | 99% | 99% | 99% | 99% | 96% | 96% | [41] |
| | 3 (Waned protection) | | | | | 62% | 62% | [41] |
| Mild disease or infection | 1 | 64% | 79% | 51% | 51% | 23% | 37% | [42, 37, 39, 40, 28, 43] |
| | 2 (Full protection) | 92% | 99% | 87% | 95% | 41% | 60% | [42, 40, 28, 43, 44, 45] |
| | 2 (Waned protection) | 29% | 77% | 19% | 49% | 0% | 0% | [33, 34, 35] |
| | 3 (Full protection) | 92% | 92% | 92% | 92% | 72% | 74% | [46] |
| | 3 (Waned protection) | | | | | 0% | 0% | [46] |
| Transmission | 1 | 45% | 45% | 33% | 33% | 20% | 20% | [31] |
| | 2 (Full protection) | 45% | 45% | 40% | 40% | 40% | 40% | [31] |
| | 2 (Waned protection) | 40% | 40% | 19% | 19% | 0% | 0% | [33, 34, 35] |
| | 3 (Full protection) | 40% | 40% | 40% | 40% | 40% | 40% | [41] |
| | 3 (Waned protection) | | | | | 0% | 0% | [41] |

**Table S4:** Vaccine effectiveness parameters for AstraZeneca (AZ), Pfizer (PF), and Moderna (Mod) by vaccine dose. "Infection" refers to vaccine effectiveness protecting an individual from being infected with SARS-CoV-2, whilst "transmission" refers to the vaccine effectiveness at preventing onward transmission by an infected individual. See Table S16 for VE assumptions for booster and booster waned sensitivity analysis for the Omicron variant.

We model cases that require hospitalisation and are hospitalised, as well as cases that require hospitalisation but are not hospitalised; for this reason we refer to vaccine effectiveness against *severe disease* and not *hospitalisation*. Vaccine effectiveness against severe disease, conditional on symptoms, acts on transition to both this compartment of individuals and those admitted to hospital.

We do not model individual vaccines separately, instead vaccine compartments are type-agnostic, and for vaccine effectiveness we compute an age-dependent weighted mean of each vaccine's effectiveness. Weights for a given age group are the proportion of each vaccine type administered to that age group as of the 24th of February 2022. Whilst we assume vaccine effectiveness does not vary by age, our weighted VE did vary across age groups, given the proportion of each vaccine (PF, AZ or Mod) administered to each age group (from data) varied substantially (Figure S3) and VE varies between vaccines (Table S4).

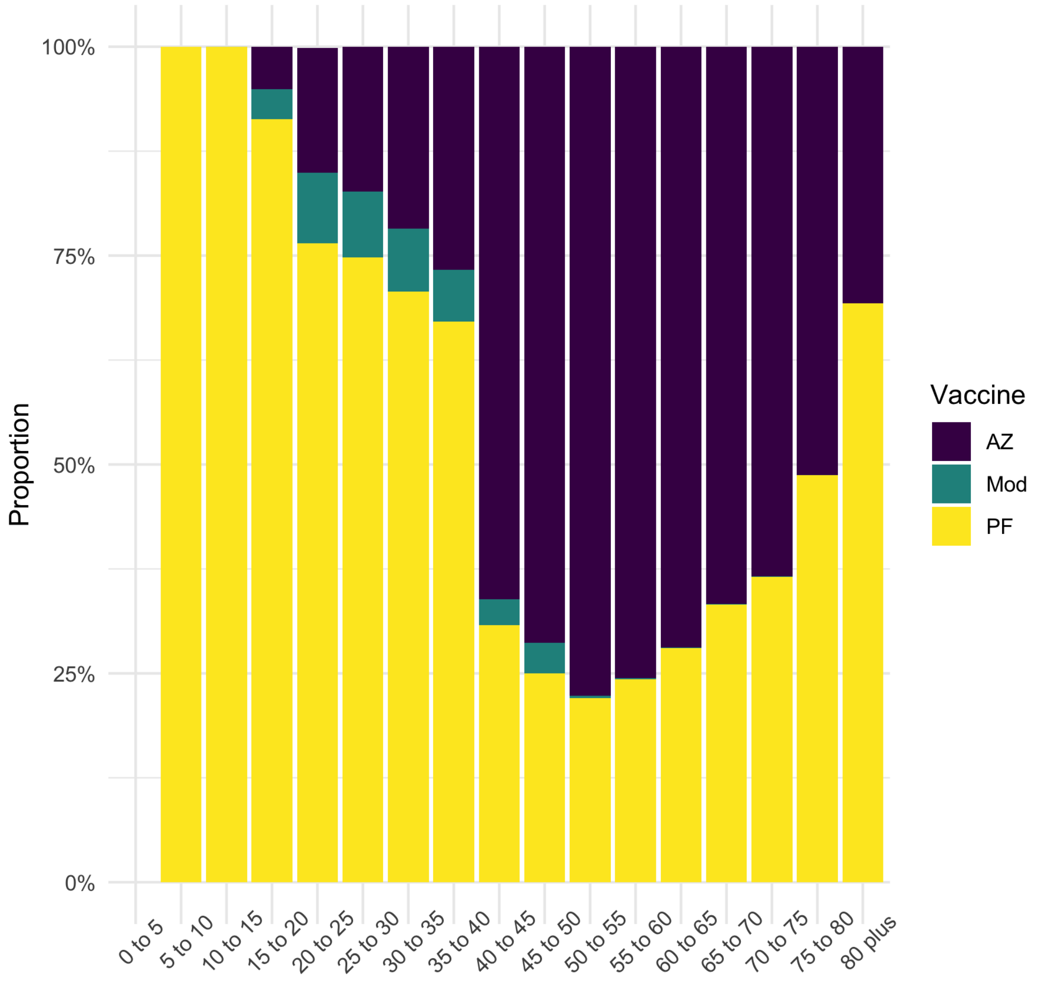

**Figure S3:** Proportion of each vaccine type: (Oxford-AstraZeneca (AZ), Pfizer-BioNTech (PF), Moderna (Mod)) dispensed to each five-year age band as of 24th February 2022. Data taken from UK Health Security Agency Immunisations database for vaccine delivery and ONS population estimates for each age group.

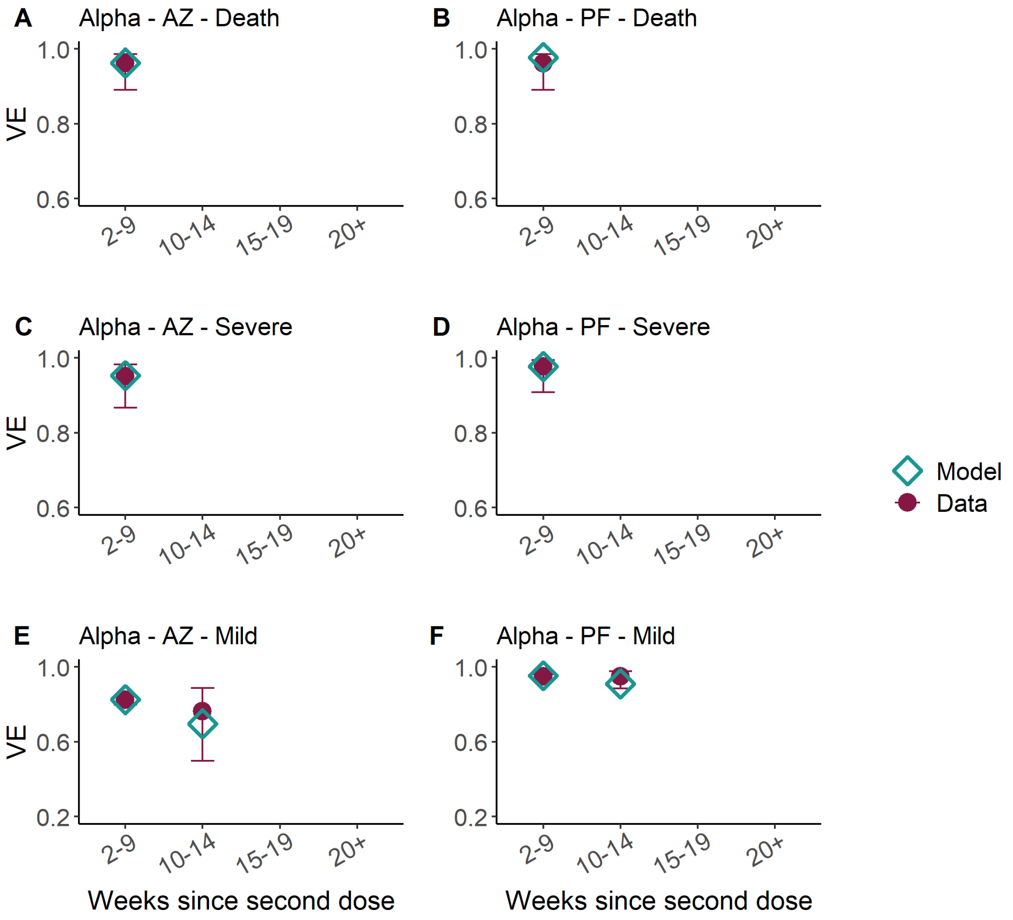

**Figure S4:** Vaccine effectiveness in weeks since second dose of AstraZeneca (AZ, left column) and Pfizer (PF, right column) vaccines against Alpha for death (top), severe disease, (middle) and mild disease/infection (bottom). We assume the same protection against infection and mild disease. Turquoise diamonds show model parameters, and the purple points estimates from data. We assumed that the Moderna vaccine has the same VE as PF.

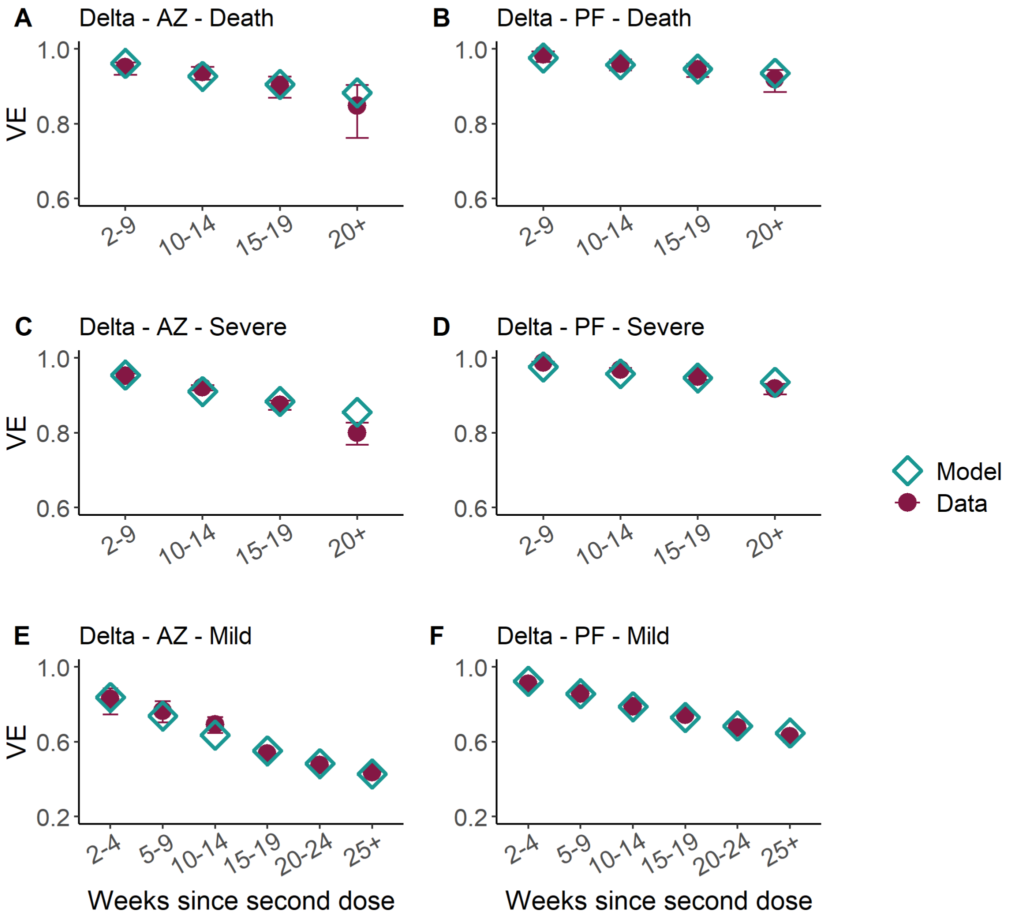

**Figure S5:** Vaccine effectiveness in weeks since second dose of AstraZeneca (AZ, left column) and Pfizer (PF, right column) vaccines against Delta for death (top), severe disease, (middle) and mild disease/infection (bottom). We assume the same protection against infection and mild disease. Turquoise diamonds show model parameters, and the purple points estimates from data. We assumed that the Moderna vaccine has the same VE as PF.

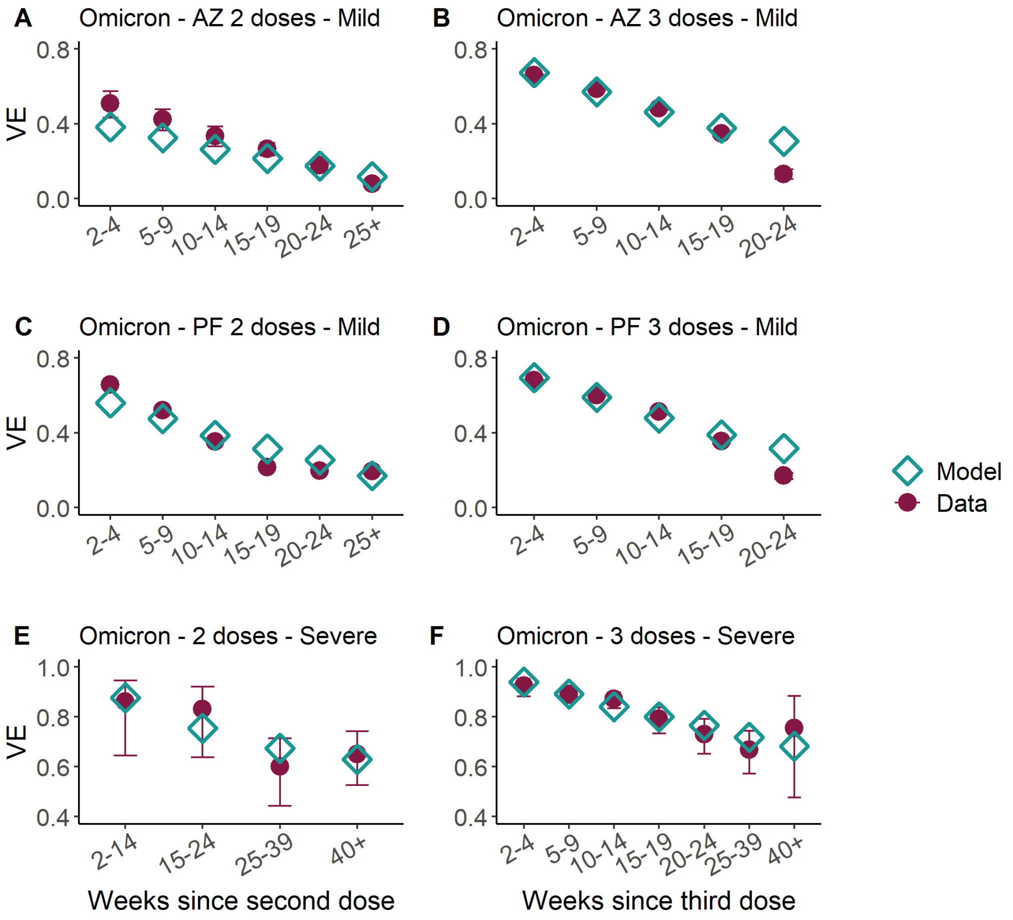

**Figure S6:** Vaccine effectiveness in weeks since second (left column) and third (booster, right column) dose of AstraZeneca (AZ, top row) and Pfizer (PF, middle row) vaccines against Omicron for mild disease (top and middle rows) and severe disease (bottom row). We assume the same protection against infection and mild disease. Turquoise diamonds show model parameters, and the purple points estimates from data. We assumed that the Moderna vaccine has the same VE as PF.

#### 3.2 Waning of vaccine-induced immunity

Vaccine-derived immunity is observed to gradually wane [35]. We assume that, upon receiving a second or booster dose, individuals firstly progress to the $V_3$ or $V_5$ vaccination strata, respectively, granting full vaccine protection. The duration spent in these strata is stochastic, with an individual's rate of progression drawn from an exponential distribution with a mean duration of 24 weeks. Note this is a simplifying assumption we make, as in real-life studies it has been shown that VE waning occurs gradually and continuously over time [33].

We fit VE protection values from second and booster doses ($V_3$ and $V_5$ compartments), as well as reduced protection VE values ($V_4$ and $V_6$ compartments) for all vaccines and health outcomes such that the mean VE for an individual at any time best replicates real world data. At the population-level, the exponential distribution waning model we employ yields a mean protection against the outcomes modelled in line with reported literature (e.g. waned second dose protection against death if infected with the Delta variant 20 weeks after vaccination of 0.89, compared to 84.8% (95% CI 76.2–90.3) from Andrews et al. [35, 46]).

Due to the limited data available for waning effectiveness against the Alpha variant, we choose to fix the log-odds difference between full second dose VE and reduced second dose VE to be the same for each variant, assuming the same proportion of protection drop-off from their initial full second dose VE for each variant. Mathematically, if $V(x)$ represents the VE of vaccine/health outcome $x$, then we define the log odds as $L(x) = \log\left(\frac{V(x)}{1-V(x)}\right)$. Thus, we fix that $L$(Reduced second dose VE vs. Delta) $-$ $L$(Full second dose VE vs. Delta) $=$ $L$(Reduced second dose VE vs. Alpha) - $L$(Full second dose VE vs. Alpha) for each health outcome. Andrews et al. provide VE estimates for timeframes of multiple weeks, against which we compared the mean population average VE from our continuous model output for the timeframe indicated. For the 20+ weeks data we compared against our model average between 20 and 29 weeks, and for the 25+ data we compared against the average between 25 and 30 weeks.These averages were fit to the data via weighted least squares, using 1/width of the associated 95% CIs reported with the data as the weights.

For first dose VE estimates we assumed the 16+ age group Delta VE values as presented in supplementary table S6 of Andrews et al. (2021) [35], however due to prioritised groups being vaccinated during the Alpha wave, the reported VEs are unlikely to be generalisable to the public at large, and as such we assumed first dose VE estimates for Alpha such that $L$(First dose VE vs. Alpha) $-$ $L$(Full second dose VE vs. Alpha) $=$ $L$(First dose VE vs. Delta) - $L$(Full second dose VE vs. Delta).

Where data was not available for certain outcomes (booster dose VE vs. the Alpha variant, booster dose VE against death for the Delta and Omicron variants), we fixed values following the logical requirements that VE against more severe outcomes must be greater than that against less severe outcomes, and assuming that VE vs. the Alpha variant must be greater than the associated protection against the Delta variant.

#### 3.3 Conditional dependencies of vaccine-immunity

We present unconditional VE in Table S4 however our model is framed as a compartmental cascade of symptom severity. Hence, unconditional VE values are converted to conditional in the model, as detailed in Table S5.

| VE vs. | Symbol / Calculation |
|---|---|
| Infection | $e_{inf}$ |
| Severe disease | $e_{SD}$ |
| Death | $e_{death}$ |
| Severe disease given infection | $e_{SD\|inf} = \dfrac{e_{SD} - e_{inf}}{(1-e_{inf})}$ |
| Death given severe disease | $e_{death\|SD} = \dfrac{e_{death} - e_{SD}}{(1-e_{inf})(1-e_{SD\|inf})}$ |

**Table S5:** Conditional vaccine effectiveness values that we model.

#### 3.4 Vaccine roll-out

The Joint Committee on Vaccination and Immunisation (JCVI) established an ordered list of individuals prioritised for vaccination in the UK, first prioritising care home residents and care home workers, and then other adults by decreasing age and clinical vulnerability [47, 48].

We assume vaccine doses are delivered in England as reported in age-stratified data from UK Health Security Agency (UKHSA) and the Department of Health and Social Care (DHSC) [13].

### 4 Model Parameterisation and Fitting

#### 4.1 Model compartments and parameters

In the following, $i$ denotes the age group of individuals ($i = [0,5),[5,10),\ldots,[75-80),[80+)$), and $j$ denotes their variant status (described in Section 4.2). Finally, $k$ denotes the index of the vaccination stratum of individuals (with $k$ corresponding to $V_k$ as defined in Table S3).

$\zeta^{i,k}(t)$ is the rate of movement from vaccination stratum $k$ to vaccination stratum $k+1$ at time $t$, for individuals in group $i$. For $k=0$, $k=2$ and $k=4$, this was set to match the observed number of daily first, second and booster (third) doses aimed to be given to each group at time step $t$. For $k=1$ this was set so that the average time to first dose effect is 3 weeks, while for $k=3$ and $k=5$, it was set so that the average time to waning of the second dose vaccine effectiveness was 24 weeks (see section 3.2). Note that due to the assumption that individuals can be boosted before the effects of their second dose have waned, vaccine-eligible individuals in stratum $k=3$ can additionally move to $k=5$ at rate $\zeta^{i,4}(t)$ (i.e. at the same rate as those whose second dose effects have waned).

We define all model compartments and parameters in Table S6 and Table S7 below, and illustrate the model structure and flows between compartments in Figure S1. The model assumes discrete time and four time steps are taken per day.

| Compartment | Definition | Erlang duration parameters $k_X$ | $\gamma_X$ | Mean | |
|---|---|---|---|---|---|
| $S^{i,k}(t)$ | Susceptible | Determined by infection dynamics | | | |
| $E^{i,j,k}(t)$ | Exposed (latent infection) | 2 | $\gamma_E^j$ | $2/\gamma_E^j$ | (see Table S2) |
| $I_A^{i,j,k}(t)$ | Asymptomatic infected | 1 | $\gamma_A^j$ | $1/\gamma_A^j$ | (see Table S2) |
| $I_P^{i,j,k}(t)$ | Presymptomatic infected (infectious) | 1 | $\gamma_P^j$ | $1/\gamma_P^j$ | (see Table S2) |
| $I_{C_1}^{i,j,k}(t)$ | Symptomatic infected (infectious) | 1 | $\gamma_{C_1}^j$ | $1/\gamma_{C_1}^j$ | (see Table S2) |
| $I_{C_2}^{i,j,k}(t)$ | Symptomatic infected (not infectious) | 1 | $\gamma_{C_2}^j$ | $1/\gamma_{C_2}^j$ | (see Table S2) |
| $G_D^{i,j,k}(t)$ | Severely diseased, leading to death (in the community) | 2 | 0.67 | 3.00 | |
| $D^{i,j,k}(t)$ | Deceased (as a result of COVID-19) | - | - | - | |
| $H_D^{i,j,k}(t)$ | Hospitalised on general ward leading to death | 2 | $0.19h_\gamma(t)$ | $10.33/h_\gamma(t)$ | (see Equation (38)) |
| $H_R^{i,j,k}(t)$ | Hospitalised on general ward leading to recovery | 1 | $0.09h_\gamma(t)$ | $10.67/h_\gamma(t)$ | (see Equation (38)) |
| $ICU_{pre}^{i,j,k}(t)$ | Awaiting admission to ICU | 1 | 0.40 | 2.50 | |
| $ICU_D^{i,j,k}(t)$ | Hospitalised in ICU, leading to death | 2 | 0.17 | 11.79 | |
| $ICU_{W_R}^{i,j,k}(t)$ | Hospitalised in ICU, leading to recovery | 1 | 0.06 | 15.61 | |
| $ICU_{W_D}^{i,j,k}(t)$ | Hospitalised in ICU, leading to death following step-down from ICU | 1 | 0.14 | 6.97 | |
| $W_R^{i,j,k}(t)$ | Step-down recovery period | 2 | $0.16h_\gamma(t)$ | $12.22/h_\gamma(t)$ | (see Equation (38)) |
| $W_D^{i,j,k}(t)$ | Step-down post-ICU period, leading to death | 1 | $0.12h_\gamma(t)$ | $8.06/h_\gamma(t)$ | (see Equation (38)) |
| $R^{i,j,k}(t)$ | Recovered | 1 | $1/1095$ | 1095 | |

**Table S6:** Definitions of model compartments shown in Figure S1, with indices: $i$ for age group ($i \in \{[0,5),[5,10),\ldots,[75-80),[80+)\}$), $j$ for variant ($j \in \{$Wildtype,Alpha,Delta,Omicron$\}$ and $k$ for vaccination strata ($k \in \{V_0,V_1,\ldots V_6\}$. Durations have Erlang$(k_X,\gamma_X)$ distributions, with mean $k_X/\gamma_X$. See Section 2 and Section 3) for further details.

| Parameter | Definition |
|---|---|
| $\lambda^{i,j,k}(t)$ | Force of infection. |
| $\gamma_x$ | Rate of progression from compartment $x$. |
| $\gamma_U(t)$ | Rate at which unconfirmed hospital patients are confirmed as infected. |
| $p_C^i$ | Probability of being symptomatic if infected. |
| $p_H^{i,j,k}(t)$ | Probability of admission to hospital, conditional on symptomatic infection. |
| $p_{G_D}^{i,j,k}(t)$ | Probability of death for severe symptomatic cases outside of hospital. |
| $p^*(t)$ | Probability of COVID-19 diagnosis confirmed prior to admission to hospital. |
| $p_{ICU}^{i,j}(t)$ | Probability of admission to ICU, conditional on hospitalisation. |
| $p_{H_D}^{i,j,k}(t)$ | Probability of death for hospitalised cases not in ICU. |
| $p_{ICU_D}^{i,j,k}(t)$ | Probability of death for cases in ICU. |
| $p_{W_D}^{i,j,k}(t)$ | Probability of death for cases after discharge from ICU. |
| $\zeta^{i,k}(t)$ | Rate of movement from vaccine strata $k$ to $k+1$. |
| $\eta_j$ | Probability of being protected against infection with variant $j$ for those recovered from earlier variants relative to those in the susceptible class. |

**Table S7:** Definitions of model parameters shown in Figure S1. These parameters define the routes of transmission through model compartments defined in Table S6.

#### 4.2 Modelling of variants

We use the $j$ dimension of the model to model variants. At any one time, there are two active variants, $j_1$ and $j_2$. If an individual in the $S$ compartment gets infected with variant $j_1$ they will move into the $j_1$ layer in the $j$ dimension for subsequent compartments, and similarly the $j_2$ layer if infected by variant $j_2$.

We also model reinfections: if any individual in the $R$ compartment gets infected with variant $j_1$ then they will move into the $j_1^{reinf}$ layer in the $j$ dimension of the $E$ compartment, and similarly the $j_2^{reinf}$ layer if infected with variant $j_2$. These additional layers allow us to account for a prior infection offering protection against severe outcomes. Note that within our model an individual can move from the $R$ compartment to the $S$ compartment - it is assumed that in doing so they lose all immunity and any potential subsequent infection is not modelled as a reinfection.

We thus have four layers in most compartments with a $j$ dimension: $j = j_1, j_2, j_1^{reinf}, j_2^{reinf}$. The sole exception is the $R$ compartment where we have an additional layer, $j_H$ accounting for individuals recovered from historic variants.

We have three two-variant phases within our model:

1. $j_1 = Wildtype,\ j_2 = Alpha,\ j_H = \{\}$
2. $j_1 = Alpha,\ j_2 = Delta,\ j_H = \{Wildtype\}$
3. $j_1 = Delta,\ j_2 = Omicron,\ j_H = \{Wildtype, Alpha\}$.

Thus, within each phase variant $j_2$ is the newer variant. We assume that individuals in the $R$ compartment can only get infected by a variant newer than the one they are recovered from. Hence, individuals recovered from $j_H$ can get infected with variant $j_1$ or $j_2$, those recovered from $j_1$ can only get infected by variant $j_2$, and those recovered from variant $j_2$ cannot get infected. Note that we assume that vaccine protections are independent of whether an infection is a reinfection or not.

At the point of switching between phases, individuals in the $R$ compartment in layers $j_1$ and $j_1^{reinf}$ move to layer $j_H$, while individuals in layers $j_2$ and $j_2^{reinf}$ move to layers $j_1$ and $j_1^{reinf}$, respectively. Meanwhile, in all other compartments with a $j$ dimension, individuals in layers $j_1$ and $j_1^{reinf}$ will remain in those layers, and individuals in layers $j_2$ and $j_2^{reinf}$ will move into the $j_1$ and $j_1^{reinf}$ layers, respectively. The $j_2$ and $j_2^{reinf}$ layers across all compartments are then empty for the introduction of the new variant.

#### 4.3 Parallel flows

In addition to compartments involved in the transmission dynamics and clinical progression, there are three parallel flows which we use for fitting to testing data from surveys: (i) one for PCR testing and (ii) two for serology testing (Figure S7), with separate flows used for testing with the EuroImmun and Roche N assays.

The PCR flow is used for fitting to data from the REACT-1 study and ONS infection survey. Upon infection, an individual enters the PCR flow in a pre-positivity compartment ($T_{PCR_{pre}}$) before moving into the PCR positivity compartment ($T_{PCR_{pos}}$) and then ultimately into the PCR negativity compartment ($T_{PCR_{neg}}$). We model the the duration of PCR positivity as an Erlang distribution. As previously detailed for our model [2, 3], we assume a mean of 12 days for this distribution for the Wildtype variant. For the VOCs Alpha, Delta and Omicron, we assume the mean of the Erlang decreased proportionally to their assumed serial interval duration (see Section 2.2 and Table S2).

With regards to the serology parallel flows, we used EuroImmun for testing NHS Blood and Transplant (NHSBT) samples from the first wave onwards, while Roche N only started being used in November 2020. Roche N tests only for seropositivity resulting from infection, whereas EuroImmun does not distinguish between seropositivity resulting from infection or from vaccination. Since our serology flows are only designed to capture seroconversion resulting from infection, we do not fit to samples using the EuroImmun assay from 15th January 2021 onwards as we can expect the vaccination to impact beyond this. After a seroconversion period ($T_{sero^1_{pre}}$ for EuroImmun, $T_{sero^2_{pre}}$ for Roche N), individuals can seroconvert ($T_{sero^1_{pos}}$ for EuroImmun, $T_{sero^2_{pos}}$ for Roche N) or not ($T_{sero^1_{neg}}$ for EuroImmun, $T_{sero^2_{neg}}$ for Roche N) ; if they do seroconvert, they eventually serorevert to $T_{sero^1_{neg}}$ or $T_{sero^2_{neg}}$ accordingly.

| | | Erlang duration parameters | | |
|---|---|---|---|---|
| Compartment | Definition | $k_X$ | $\gamma_X$ | Mean |
| $T^i_{PCR_{pre}}(t)$ | Pre-PCR positive | 1 | 0.5 | 2 |
| $T^i_{PCR_{pos}}(t)$ | True PCR positive | 1 | 0.083 | 12.05 |
| $T^i_{PCR_{neg}}(t)$ | True PCR negative (after infection) | - | - | - |
| $T^i_{sero^1_{pre}}(t)$ | Pre-seropositive for EuroImmun assay | 1 | 0.077 | 13 |
| $T^i_{sero^1_{pos}}(t)$ | True seropositive for EuroImmun assay | 1 | 0.0025 | 400 |
| $T^i_{sero^1_{neg}}(t)$ | True seropositive (after infection) for EuroImmun assay | - | - | - |
| $T^i_{sero^2_{pre}}(t)$ | Pre-seropositive for Roche N assay | 1 | 0.077 | 13 |
| $T^i_{sero^2_{pos}}(t)$ | True seropositive for Roche N assay | 1 | 0.001 | 1000 |
| $T^i_{sero^2_{neg}}(t)$ | True seropositive (after infection) for Roche N assay | - | - | - |

**Table S8:** Definitions of model parallel flow compartments shown in Figure S7, with indices: $i$ for age group ($i \in \{[0,5),[5,10),\ldots,[75-80),[80+)\}$). Durations (in days) have Erlang$(k_X,\gamma_X)$ distributions, with mean $k_X/\gamma_X$.

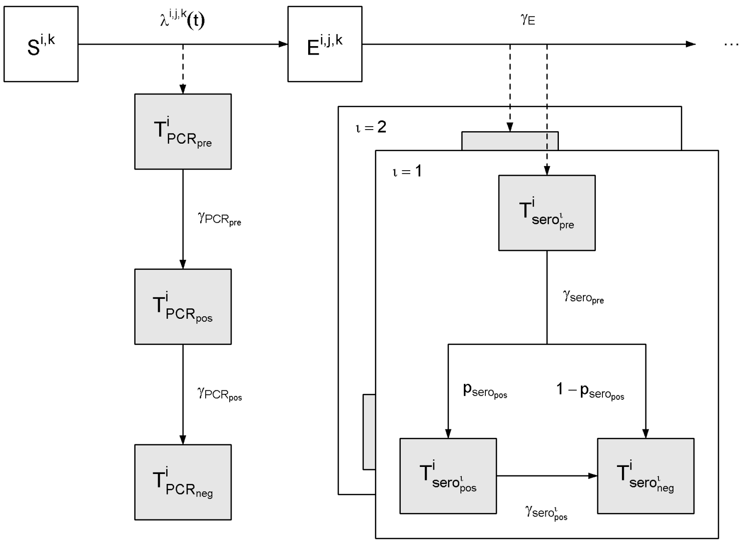

**Figure S7:** PCR positivity and seropositivity model structure flow diagram. Upon infection, an individual enters the pre-positive PCR compartment ($T_{PCR_{pre}}$) before moving into the PCR positivity compartment ($T_{PCR_{pos}}$) and then into the PCR negativity compartment ($T_{PCR_{neg}}$). After a seroreversion period ($T_{sero_{pre}}$), individuals can seroconvert ($T_{sero_{pos}}$) or not ($T_{sero_{neg}}$); if they do seroconvert, they eventually serorevert to $T_{sero_{pos}}$.

### 4.4 Equations

#### 4.4.1 Force of infection

We let $\chi^{i,j,k}$ be the susceptibility to variant $j$ of a susceptible individual in group $i$ and vaccine stratum $k$, relative to a non vaccinated individual (so that $\chi^{i,j,0} = 1$ for all $i$ and $j$), given by

$$\chi^{i,j,k} = (1 - e^{i,j,k}_{inf}), \tag{2}$$

where $e^{i,j,k}_{inf}$ is the vaccine effectiveness against infection of variant $j$ in vaccine strata $k$ (Table S5), scaled across vaccine types according to the distribution presented in Figure S3.

We let $\xi^{i,j,k}$ be the infectivity of an individual in group $i$ and vaccine stratum $k$ infected with variant $j$ relative to a non vaccinated individual infected with the Wildtype variant (so that $\xi^{i,Wildtype,0} = 1$). This infectivity captures both the vaccine effectiveness against infectiousness as presented in Table S4 and also the increased transmissibility of an emerging variant compared to the one being replaced. As such $\xi^{i,j,k}$ is equal to

$$\xi^{i,j,k} = (1 - e^{i,j,k}_{ins})\sigma_j, \tag{3}$$

where $e^{i,j,k}_{ins}$ is the vaccine effectiveness against infectiousness of variant $j$ in vaccine strata $k$ as defined in Table S4, scaled across vaccine types according to the distribution presented in Figure S3, and $\sigma_j$ is the region-specific transmission advantage of variant $j$ over the Wildtype which we further parameterise as

$$\sigma_j = \begin{cases} 1 & \text{if } j = Wildtype \\ \sigma_{Wildtype}\sigma_{Alpha/Wildtype} & \text{if } j = Alpha \\ \sigma_{Alpha}\sigma_{Delta/Alpha} & \text{if } j = Delta \\ \sigma_{Delta}\sigma_{Omicron/Delta} & \text{if } j = Omicron \end{cases} \tag{4}$$

and we fit $\sigma_{Alpha/Wildtype}$, $\sigma_{Delta/Alpha}$ and $\sigma_{Omicron/Delta}$, which are the region-specific transmission advantages of the Alpha over Wildtype, Delta over Alpha and Omicron over Delta, respectively. We use uniform prior distributions between 0 and 3 for each of these (Table S12).

We let $\Theta_{i,j,k}(t)$ be the number of infectious individuals with variant $j$ in group $i$ and vaccination stratum $k$, weighted by infectivity, given by:

$$\Theta_{i,j,k}(t) = \xi^{i,j,k}\left(\theta_{I_A} I_A^{i,j,k}(t) + I_P^{i,j,k}(t) + I_{C_1}^{i,j,k}(t)\right). \tag{5}$$

where $\theta_{I_A}$ is the infectivity of an asymptomatic infected individual, relative to a symptomatic individual infected with the same variant, and in the same vaccination strata.

The force of infection, $\lambda^{i,j,k}(t)$, of variant $j \in \{j_1, j_2\}$ on a susceptible individual in group $i \in \{[0,5), \ldots, [75,80), [80+)\}$ and vaccination stratum $k = 0,1,\ldots,6$ is then given by

$$\lambda^{i,j,k}(t) = \chi^{i,j,k} \sum_{i'} m_{i,i'}(t) \sum_{k'} \left(\Theta_{i',j,k'}(t) + \Theta_{i',j^{reinf},k'}(t)\right) \tag{6}$$

where $m_{i,i'}(t)$ is the (symmetric) time-varying person-to-person transmission rate from group $i'$ to group $i$.

We let $\Lambda^{i,k}(t)$ be the total force of infection on a susceptible individual in group $i$ and vaccination stratum $k$, i.e.

$$\Lambda^{i,k}(t) = \lambda^{i,j_1,k}(t) + \lambda^{i,j_2,k}(t). \tag{7}$$

We let $\eta_{inf}^{j}$ be the parameter for cross immunity against infection from earlier variants against variant $j$ Table S7. The force of infection of variant $j_1$ on an individual recovered from historic variants $j_H$ in group $i$ and vaccine stratum $k$ is $(1-\eta_{j_1})\lambda^{i,j_1,k}(t)$. There is no force of infection of variant $j_1$ on individuals recovered from variants $j_1$ or $j_2$. The force of infection of variant $j_2$ on an individual recovered from variant $j_1$ or historic variants $j_H$ in group $i$ and vaccine stratum $k$ is $(1-\eta_{j_2})\lambda^{i,j_2,k}(t)$. There is no force of infection of variant $j_2$ on individuals recovered from variant $j_2$.

Transmission between different age groups $(i,i') \in \{[0,5),\ldots,[75,80),[80+)\}^2$ was parameterised as follows:

$$m_{i,i'}(t) = \beta(t) c_{i,i'}, \tag{8}$$

where $c_{i,i'}$ is the (symmetric) person-to-person contact rate between age group $i$ and $i'$, derived from pre-pandemic data from the POLYMOD survey [4] for the United Kingdom. For each region, the `socialmixr` package [49] was used to derive the contact matrix between different age groups $(i,i') \in \{[0,5),\ldots,[75,80),[80+)\}^2$, which was then scaled by the regional population demography to yield the required person-to-person daily contact rate matrix, $c_{i,i'}$.

$\beta(t)$ is the time-varying transmission rate which encompasses both changes over time in transmission efficiency (e.g. due to temperature) and temporal changes in the overall level of contacts in the population (due to changes in policy and behaviours).

We assumed $\beta(t)$ to be piecewise linear:

$$\beta(t) = \begin{cases} \beta_i, & \text{if } t \le t_i,\ i = 1 \\ \dfrac{t_i - t}{t_i - t_{i-1}}\beta_{i-1} + \dfrac{t - t_{i-1}}{t_i - t_{i-1}}\beta_i, & \text{if } t_{i-1} < t \le t_i,\ i \in \{2,\ldots,36\} \\ \beta_i & \text{if } t > t_i,\ i = 36 \end{cases} \tag{9}$$

with 36 change points $t_i$ corresponding to major policy implementations or lifting and other relevant changes in contact rates (e.g. school holidays) (see Table S12).

#### 4.4.2 Seeding of variants

We seed each variant $j$ at a daily rate $\phi_j$, over a period of $\nu_j$ days from time $t_j$. All seeding infections are from the S to E compartment in the 15-19 year old group and unvaccinated class.

For the Wildtype we seed at a rate of 10 per million of the total regional population per day, over a 1-day period, so $\phi_{Wildtype} = \frac{1}{100{,}000}\sum_i N^i$ and $\nu_{Wildtype} = 1$.

For the Alpha, Delta and Omicron variants we seed each at a rate of 2 per million of the total regional population per day, over a 7-day period, so for $j \in \{Alpha, Delta, Omicron\}$, $\phi_j = \frac{1}{500{,}000}\sum_i N^i$ and $\nu_j = 7$.

The seeding dates $t_{Wildtype}$ (which corresponds to the start date of the regional epidemic), $t_{Alpha}$, $t_{Delta}$ and $t_{Omicron}$ are fitted parameters (see Table S12).

The seeding rate in age group $i$ and vaccine stratum $k$ of variant $j$ is then given by

$$\delta^{i,j,k}(t) = \begin{cases} \phi_j & \text{if } i = [15,20),\ j \in \{Wildtype,\ Alpha,\ Delta,\ Omicron\},\ k = 0 \text{ and } t_j \le t < t_j + \nu_j, \\ 0 & \text{otherwise,} \end{cases} \quad (10)$$

where $\delta^{i,j,k}(t)$ is the daily seeding rate of variant $j$ (stratified by age and vaccination strata).

#### 4.4.3 Pathway probabilities and rates

The movement between model compartments is primarily dictated by the parameters $p_x^{i,j,k}$, defining the probability of progressing to compartment $x$ (Table S7), as well as rate parameters $\gamma$. These parameters vary between age groups ($i$), variant of infection ($j$), and vaccine strata ($k$). Additionally, for some of these parameters, we allowed them to vary over time by fitting them as a piecewise linear function. Further details on the definition of changepoints for piecewise parameters are presented in the following section (Section 4.4.4). This section outlines how the pathway probabilities defining movement between model compartments are formally defined and calculated. Some of the probabilities are affected by whether an infection is a reinfection or not. Equations are defined for $i \in \{[0,5),\ \ldots, [75,80), [80+)\}$ and vaccination stratum $k = 0, 1, \ldots, 6$.

The probability that an infected individual will have a symptomatic infection depends only on their age group $i$ and is given by (as in Knock et al [1, 2, 3]):

$$p_C^i = \begin{cases} 0.25 & \text{for i = [0,5)} \\ 0.26875 & \text{for i = [5,10)} \\ 0.325 & \text{for i = [10,15)} \\ 0.41875 & \text{for i = [15,20)} \\ 0.55 & \text{for i = [20,25, ...[75,80), [80+).} \end{cases} \quad (11)$$

The probability that a symptomatic individual has severe disease requiring hospitalisation is given by, for $j \in \{j_1, j_2\}$,

$$p_H^{i,j,k}(t) = \min\left\{h_H(t)\psi_H^i\left(1 - e_{SD|sympt}^{i,j,k}\right)\pi_H^j,\ 1\right\}, \quad (12)$$

$$p_H^{i,j^{reinf},k}(t) = \min\left\{h_H(t)\psi_H^i\left(1 - e_{SD|sympt}^{i,j,k}\right)\pi_H^j\left(1 - \eta_H^j\right),\ 1\right\}, \quad (13)$$

where $\psi_H^i$ represents the age scaling (such that $\psi_H^i = 1$ for the group with the maximum probability; this has been newly tuned to the data and is given in Table S9), and $h_H(t)$ has a piecewise linear form with the following changepoints (see Table S10):

$$h_H = (t)\begin{cases} p_{H,1}^{max} & \text{on (and before) 04/11/21,} \\ p_{H,2}^{max} & \text{on (and after) 31/12/21} \end{cases} \quad (14)$$

and $\pi_H^j$ is a multiplier accounting for the changing severity of the variants of concern with respect to the Wildtype variant (see Section 2.4):

$$\pi_H^j = \begin{cases} 1 & \text{if } j = Wildtype, \\ \pi_H^{Wildtype}\pi_H^{Alpha/Wildtype} & \text{if } j = Alpha, \\ \pi_H^{Alpha}\pi_H^{Delta/Alpha} & \text{if } j = Delta, \\ \pi_H^{Delta}\pi_H^{Omicron/Delta} & \text{if } j = Omicron, \end{cases} \quad (15)$$

where $\pi_H^{Alpha/Wildtype}$, $\pi_H^{Delta/Alpha}$ and $\pi_H^{Omicron/Delta}$ are the relative probabilities of severe disease requiring hospitalisation given infection for Alpha over Wildtype, Delta over Alpha and Omicron over Delta, respectively. We fit these three parameters, see S12.

The probability that an individual dies in the community given they have severe disease is given by, for $j \in \{j_1, j_2\}$:

$$p_{G_D}^{i,j,k}(t) = \min\left\{h_{G_D}(t)\psi_{G_D}^i\left(1 - e_{death|SD}^{i,j,k}\right)\pi_D^j,\ 1\right\}, \quad (16)$$

$$p_{G_D}^{i,j^{reinf},k}(t) = \min\left\{h_{G_D}(t)\psi_{G_D}^i\left(1 - e_{death|SD}^{i,j,k}\right)\pi_D^j\left(1 - \eta_D^j\right),\ 1\right\}, \quad (17)$$

where $h_{G_D}$ has a piecewise linear form with the following changepoints (see Table S10):

$$h_{G_D}(t) = \begin{cases} \pi_{G_D,1}^{max} & \text{on (and before) 01/05/20,} \\ \pi_{G_D,2}^{max} & \text{on (and after) 01/07/20,} \end{cases} \tag{18}$$

$psi_{G_D}$ is an age multiplier (newly tuned to the data and presented in Table S9), and $\pi_D^j$ is a multiplier accounting for the changing severity of the variants of concern with respect to the Wildtype variant (see Section 2.4):

$$\pi_D^j = \begin{cases} 1 & \text{if } j = Wildtype, \\ \pi_D^{Wildtype} \pi_D^{Alpha/Wildtype} & \text{if } j = Alpha, \\ \pi_D^{Alpha} \pi_D^{Delta/Alpha} & \text{if } j = Delta, \\ \pi_D^{Delta} \pi_D^{Omicron/Delta} & \text{if } j = Omicron, \end{cases} \tag{19}$$

where $\pi_D^{Alpha/Wildtype}$, $\pi_D^{Delta/Alpha}$ and $\pi_D^{Omicron/Delta}$ are the relative probabilities of death given severe disease for Alpha over Wildtype, Delta over Alpha and Omicron over Delta, respectively. We fit these three parameters, see S12.

The probability that an individual will be admitted to ICU given that they have been hospitalised is given by, for $j \in \{j_1, j_2\}$,

$$p_{ICU}^{i,j}(t) = \min\left\{ h_{ICU}(t) \psi_{ICU}^i \pi_{ICU}^j,\ 1 \right\}, \tag{20}$$

$$p_{ICU}^{i,j^{reinf}}(t) = p_{ICU}^{i,j}(t) \tag{21}$$

where $\psi_{ICU}^i$ is the age scaling (as in [1, 2, 3] and presented again in Table S9), $h_{ICU}(t)$ has a piecewise linear form with the following changepoints (see Table S10):

$$h_{ICU} = (t) \begin{cases} p_{ICU,1}^{max} & \text{on (and before) 01/04/20,} \\ p_{ICU,2}^{max} & \text{on 01/06/20,} \end{cases} \tag{22}$$

and $\pi_{ICU}^j$ is a multiplier accounting for the changing severity of the variants of concern with respect to the Wildtype variant (see Section 2.4):

$$\pi_{ICU}^j = \begin{cases} 1 & \text{if } j = Wildtype, \\ \pi_{ICU}^{Wildtype} \pi_{ICU}^{Alpha/Wildtype} & \text{if } j = Alpha, \\ \pi_{ICU}^{Alpha} \pi_{ICU}^{Delta/Alpha} & \text{if } j = Delta, \\ \pi_{ICU}^{Delta} \pi_{ICU}^{Omicron/Delta} & \text{if } j = Omicron, \end{cases} \tag{23}$$

where $\pi_{ICU}^{Alpha/Wildtype}$, $\pi_{ICU}^{Delta/Alpha}$ and $\pi_{ICU}^{Omicron/Delta}$ are the relative probabilities of admission to ICU given hospitalisation for Alpha over Wildtype, Delta over Alpha and Omicron over Delta, respectively. We fit these three parameters, see S12.

The probability that an individual will die in general beds given that they are not admitted to ICU is, for $j \in \{j_1, j_2\}$,

$$p_{H_D}^{i,j,k}(t) = \min\left\{ p_{H_D}^{max} h_D(t) \psi_{H_D}^i \left(1 - e_{death|SD}^{i,j,k}\right) \pi_D^j, 1 \right\}, \tag{24}$$

$$p_{H_D}^{i,j^{reinf},k}(t) = \min\left\{ p_{H_D}^{max} h_D(t) \psi_{H_D}^i \left(1 - e_{death|SD}^{i,j,k}\right) \pi_D^j \left(1 - \eta_D^j\right), 1 \right\}, \tag{25}$$

where $\psi_{H_D}^i$ represents the age scaling (as in [1, 2, 3] and presented again in Table S9) as in [1, 2, 3]), and $h_D(t)$ has a piecewise linear form with the following changepoints (see Table S10):

$$h_D = (t) \begin{cases} 1 & \text{on (and before) 01/04/20,} \\ \mu_{D,1}^{max} & \text{on 16/06/20,} \\ \mu_{D,1}^{max} & \text{on 15/09/20,} \\ \mu_{D,2}^{max} & \text{on 01/11/20,} \\ \mu_{D,2}^{max} & \text{on 15/12/20,} \\ \mu_{D,3}^{max} & \text{on 04/02/21,} \\ \mu_{D,4}^{max} & \text{on 01/04/21,} \\ \mu_{D,4}^{max} & \text{on 04/11/21,} \\ \mu_{D,5}^{max} & \text{on (and after) 31/12/21.} \end{cases} \tag{26}$$

The probability that an individual dies in ICU given that they are not admitted to ICU is given by, for $j \in \{j_1, j_2\}$:

$$p^{i,j,k}_{ICU_D}(t) = \min\left\{p^{max}_{ICU_D} h_D(t) \psi^i_{ICU_D}\left(1 - e^{i,j,k}_{death|SD}\right)\pi^j_D,\ 1\right\} \tag{27}$$

$$p^{i,j^{reinf},k}_{ICU_D}(t) = \min\left\{p^{max}_{ICU_D} h_D(t) \psi^i_{ICU_D}\left(1 - e^{i,j,k}_{death|SD}\right)\pi^j_D\left(1 - \eta^j_D\right),\ 1\right\} \tag{28}$$

where $\psi^i_{ICU_D}$ represents the age scaling (as in [1, 2, 3] and presented again in Table S9).

The probability that an individual who has been in ICU dies in stepdown beds given that they have not died in ICU is given by, for $j \in \{j_1, j_2\}$:

$$p^{i,j,k}_{W_D}(t) = \min\left\{p^{max}_{W_D} h_D(t) \psi^i_{W_D}\left(1 - e^{i,j,k}_{death|SD}\right)\pi^j_D,\ 1\right\}, \tag{29}$$

$$p^{i,j^{reinf},k}_{W_D}(t) = \min\left\{p^{max}_{W_D} h_D(t) \psi^i_{W_D}\left(1 - e^{i,j,k}_{death|SD}\right)\pi^j_D\left(1 - \eta^j_D\right),\ 1\right\}, \tag{30}$$

where $\psi^i_{W_D}$ represents the age scaling (as in [1, 2, 3] and presented again in Table S9).

Finally, the probability of individuals having had a COVID-19 diagnosis confirmed prior to admission to hospital, $p^*(t)$ has a piecewise linear form with the following changepoints:

$$p^*(t) = \begin{cases} 0.1 & \text{on (and before) 15/03/20,} \\ 0.42 & \text{on 01/07/20,} \\ 0.2 & \text{on 20/09/20,} \\ 0.45 & \text{on 27/06/21,} \\ 0.45 & \text{on 01/12/21,} \\ 0.33 & \text{on (and after) 01/01/22.} \end{cases} \tag{31}$$

These were informed by data on COVID-19 admissions and inpatient diagnoses from NHS England [50]. Meanwhile, the rate at which unconfirmed covid patients in hospital become confirmed, $\gamma_U(t)$, has a piecewise linear form with the following changepoints:

$$\gamma_U(t) = \begin{cases} 1/2.1 & \text{on (and before) 15/03/20,} \\ 1/1.2 & \text{on 05/04/20,} \\ 1/1.2 & \text{on 15/09/20,} \\ 1/0.7 & \text{on 15/11/20,} \\ 1/1.3 & \text{on 10/12/20,} \\ 1/0.7 & \text{on 20/01/21,} \\ 1/0.4 & \text{on 01/08/21,} \\ 1/0.9 & \text{on 01/03/22.} \end{cases} \tag{32}$$

These were informed by data on times to diagnosis from CHESS [51].

In addition, the duration rates for some hospital compartments are time-varying to account for changes in length of stay over time. We let

$$\gamma_{H_R}(t) = h_\gamma(t)\gamma_{H_R} \tag{33}$$

$$\gamma_{H_D}(t) = h_\gamma(t)\gamma_{H_D} \tag{34}$$

$$\gamma_{W_R}(t) = h_\gamma(t)\gamma_{W_R} \tag{35}$$

$$\gamma_{W_D}(t) = h_\gamma(t)\gamma_{W_D} \tag{36}$$

(37)

where $h_\gamma(t)$ has a piecewise linear form with changepoints given by

$$h_\gamma(t) = \begin{cases} 1 & \text{on (and before) 01/12/20,} \\ 1/\mu_{\gamma_H,1} & \text{on 01/01/21,} \\ 1/\mu_{\gamma_H,2} & \text{on 01/03/21,} \\ 1/\mu_{\gamma_H,3} & \text{on 01/06/21,} \\ 1/\mu_{\gamma_H,4} & \text{on (and after) 01/09/21.} \end{cases} \tag{38}$$

| Age group $i$ | $\psi^i_H$ | $\psi^i_{G_D}$ | $\psi^i_{ICU}$ | $\psi^i_{H_D}$ | $\psi^i_{ICU_D}$ | $\psi^i_{W_D}$ |
|---|---|---|---|---|---|---|
| $[0,5)$ | 0.0149 | 0.0000 | 0.2428 | 0.0386 | 0.2823 | 0.0911 |
| $[5,10)$ | 0.0027 | 0.0078 | 0.2891 | 0.0365 | 0.2861 | 0.0830 |
| $[10,15)$ | 0.0033 | 0.0033 | 0.3377 | 0.0353 | 0.2913 | 0.0775 |
| $[15,20)$ | 0.0065 | 0.0123 | 0.3894 | 0.0351 | 0.2991 | 0.0744 |
| $[20,25)$ | 0.0129 | 0.0139 | 0.4426 | 0.0362 | 0.3103 | 0.0739 |
| $[25,30)$ | 0.0191 | 0.0129 | 0.5027 | 0.0391 | 0.3276 | 0.0760 |
| $[30,35)$ | 0.0269 | 0.0242 | 0.5697 | 0.0447 | 0.3526 | 0.0802 |
| $[35,40)$ | 0.0255 | 0.0420 | 0.6530 | 0.0552 | 0.3909 | 0.0860 |
| $[40,45)$ | 0.0287 | 0.0604 | 0.7559 | 0.0743 | 0.4465 | 0.0927 |
| $[45,50)$ | 0.0336 | 0.0801 | 0.8659 | 0.1067 | 0.5196 | 0.1016 |
| $[50,55)$ | 0.0568 | 0.1054 | 0.9541 | 0.1568 | 0.6044 | 0.1167 |
| $[55,60)$ | 0.0769 | 0.1317 | 1.0000 | 0.2385 | 0.7047 | 0.1482 |
| $[60,65)$ | 0.1075 | 0.1679 | 0.9720 | 0.3528 | 0.8057 | 0.2113 |
| $[65,70)$ | 0.1364 | 0.2165 | 0.8544 | 0.5020 | 0.8988 | 0.3315 |
| $[70,75)$ | 0.2452 | 0.2992 | 0.6454 | 0.6750 | 0.9692 | 0.5263 |
| $[75,80)$ | 0.4218 | 0.4327 | 0.4024 | 0.8319 | 1.0000 | 0.7531 |
| $[80+)$ | 1.0000 | 1.0000 | 0.1074 | 1.0000 | 0.9178 | 1.0000 |

**Table S9:** Age multipliers for pathway probabilities. These were estimated using a progression model fitted with MCMC to age-stratified linelist data from CHESS [51], as previously published by our group [1].

#### 4.4.4 Time-varying severity parameters

Severity pathway probabilities $p_H^{i,j,k}(t)$, $p_{ICU}^{i,j,k}(t)$, $p_{G_D}^{i,j,k}(t)$ and $p_{H_D}^{i,j,k}(t)$ are fitted regionally as time-varying parameters using a piecewise form, as defined in the section above. We fit as few changepoints as possible to: a) allow the model flexibility to capture variations in severity over time given underlying changes in healthcare practices and/or population healthcare seeking behaviours (Table S10); and b) avoid over-fitting and identifiability issues across different parameters governing severity dynamics.

| Parameter | Dates | Rationale | Reference |
|---|---|---|---|
| $h_H(t)$ | 04-11-2021<br>31-12-2021 | Approval and roll-out of novel outpatient treatments for COVID-19. | [52, 53, 54, 55] |
| $h_{ICU}(t)$ | 01-04-2020<br>01-06-2020 | First hospital treatment protocols and use of dexamethasone established. | [56, 57] |
| $h_{G_D}(t)$ | 01-05-2020<br>01-07-2020 | Potential change in healthcare seeking behaviour or case management after the first wave in the community/carehomes. | NA |
| $h_D(t)$ | 01-04-2020 | First wave | NA |
| | 01-07-2020 to 15-09-2020 | Trough after first wave | NA |
| | 15-10-2020 to 01-12-2020 | Winter 2020/21 wave | NA |
| | 04-02-2021 | Trough after winter 2020/21 wave | NA |
| | 01-04-2021 to 04-11-2021<br>31-12-2021 | Approval and roll-out of novel oral treatments for COVID-19 | [52, 53, 54, 55] |

**Table S10:** Fitted changepoints for time-varying severity parameters with piecewise form. We explored alternative change points for $\mu_D(t)$ in sensitivity analysis (see section 5.3).

#### 4.4.5 Compartmental model equations

To clearly illustrate the model dynamics, we describe a deterministic version of the model in differential equations (40)-(80), followed by the stochastic implementation used in the analysis. Full definitions of compartments and model parameters are set out in Tables S6 and S7.

Unless otherwise specified, $\sum_i$ refers to the sum across age groups (i.e. $i \in \{[0,5), \ldots, [75,80), [80+)\}$), $\sum_j$ refers to the sum across all combinations of co-circulating variants ($j_1$, $j_2$, $j_1^{reinf}$, $j_2^{reinf}$), and $\sum_k$ refers to the sum across all vaccination strata ($k \in \{0,1,...,6\}$).

In the following model equations we use $\mathbb{1}_A$ as an indicator function, such that

$$\mathbb{1}_A(j) := \begin{cases} 1 & \text{if } j \in A, \\ 0 & \text{if } j \notin A. \end{cases} \tag{39}$$

Further note that we split some compartments in two distinct compartments. For example, the exposed class, $E^{i,j,k}$, is modelled via two separate compartments, $E^{i,j,k,1}$ and $E^{i,j,k,2}$ (equations (41) and (42)). This is to be able to capture a non exponentially distributed duration of stay in certain compartments; the split allows us to model the duration of stay as an Erlang distribution instead (sum of independent exponential distributions) [58].

$$\begin{aligned} \frac{dS^{i,k}(t)}{dt} &= \zeta^{i,k-1}(t)S^{i,k-1}(t) + \mathbb{1}_{\{5\}}(k)\zeta^{i,4}(t)S^{i,3}(t) - \left(\zeta^{i,k}(t) + \mathbb{1}_{\{3\}}(k)\zeta^{i,4}(t) + \Lambda^{i,k}(t)\right)S^{i,k}(t) \\ &\quad - \sum_j \left(\delta^{i,j,k}(t) + \gamma_R R^{i,j,k}(t)\right) + \gamma_R R^{i,j_H,k} \end{aligned} \tag{40}$$

$$\begin{aligned} \frac{dE^{i,j,k,1}(t)}{dt} &= \mathbb{1}_{\{j_1,j_2\}}(j)\lambda^{i,j,k}(t)S^{i,k}(t) + \mathbb{1}_{\left\{j_1^{reinf},j_2^{reinf}\right\}}(j)\left(1-\eta_j\right)\lambda^{i,j,k}(t)R^{i,j_H,k}(t) \\ &\quad + \mathbb{1}_{\left\{j_2^{reinf}\right\}}(j)\left(1-\eta_j\right)\lambda^{i,j,k}(t)\left(R^{i,j_1,k}(t) + R^{i,j_1^{reinf},k}(t)\right) + \zeta^{i,k-1}(t)E^{i,j,k-1,1}(t) \\ &\quad + \mathbb{1}_{\{5\}}(k)\zeta^{i,4}(t)E^{i,j,3,1}(t) + \delta^{i,j,k}(t) - \left(\gamma_E^j + \zeta^{i,k}(t) + \mathbb{1}_{\{3\}}(k)\zeta^{i,4}(t)\right)E^{i,j,k,1}(t) \end{aligned} \tag{41}$$

$$\begin{aligned} \frac{dE^{i,j,k,2}(t)}{dt} &= \gamma_E^j E^{i,j,k,1}(t) + \zeta^{i,k-1}(t)E^{i,j,k-1,2}(t) + \mathbb{1}_{\{5\}}(k)\zeta^{i,4}(t)E^{i,j,3,2}(t) \\ &\quad - \left(\gamma_E^j + \zeta^{i,k}(t) + \mathbb{1}_{\{3\}}(k)\zeta^{i,4}(t)\right)E^{i,j,k,2}(t) \end{aligned} \tag{42}$$

$$\begin{aligned} \frac{dI_A^{i,j,k}(t)}{dt} &= \left(1-p_C^i\right)\gamma_E^j E^{i,j,k,2}(t) + \zeta^{i,k-1}(t)I_A^{i,j,k-1}(t) + \mathbb{1}_{\{5\}}(k)\zeta^{i,4}(t)I_A^{i,j,3}(t) \\ &\quad - \left(\gamma_A^j + \zeta^{i,k}(t) + \mathbb{1}_{\{3\}}(k)\zeta^{i,4}(t)\right)I_A^{i,j,k}(t) \end{aligned} \tag{43}$$

$$\begin{aligned} \frac{dI_P^{i,j,k}(t)}{dt} &= p_C^i\gamma_E^j E^{i,j,k,2}(t) + \zeta^{i,k-1}(t)I_P^{i,j,k-1}(t) + \mathbb{1}_{\{5\}}(k)\zeta^{i,4}(t)I_P^{i,j,3}(t) \\ &\quad - \left(\gamma_P^j + \zeta^{i,k}(t) + \mathbb{1}_{\{3\}}(k)\zeta^{i,4}(t)\right)I_P^{i,j,k}(t) \end{aligned} \tag{44}$$

$$\frac{dI_{C_1}^{i,j,k}(t)}{dt} = \gamma_P^j I_P^{i,j,k}(t) - \gamma_{C_1}^j I_{C_1}^{i,j,k}(t) \tag{45}$$

$$\frac{dI_{C_2}^{i,j,k}(t)}{dt} = \gamma_{C_1}^j I_{C_1}^{i,j,k}(t) - \gamma_{C_2}^j I_{C_2}^{i,j,k}(t) \tag{46}$$

$$\frac{dG_D^{i,j,k,1}(t)}{dt} = p_H^{i,j,k}(t)p_{G_D}^{i,j,k}(t)\gamma_{C_2}^j I_{C_2}^{i,j,k}(t) - \gamma_{G_D}G_D^{i,j,k,1}(t) \tag{47}$$

$$\frac{dG_D^{i,j,k,2}(t)}{dt} = \gamma_{G_D}G_D^{i,j,k,1}(t) - \gamma_{G_D}G_D^{i,j,k,2}(t) \tag{48}$$

$$\begin{aligned} \frac{dICU_{pre}^{i,j,k}(t)}{dt} &= p_H^{i,j,k}(t)\left(1-p_{G_D}^{i,j,k}(t)\right)\left(1-p^*(t)\right)p_{ICU}^{i,j}(t)\gamma_{C_2}^j I_{C_2}^{i,j,k}(t) \\ &\quad - \left(\gamma_{ICU_{pre}} + \gamma_U(t)\right)ICU_{pre}^{i,j,k}(t) \end{aligned} \tag{49}$$

$$\begin{aligned} \frac{dICU_{pre^*}^{i,j,k}(t)}{dt} &= p_H^{i,j,k}(t)\left(1-p_{G_D}^{i,j,k}(t)\right)p^*(t)p_{ICU}^{i,j}(t)\gamma_{C_2}^j I_{C_2}^{i,j,k}(t) - \gamma_{ICU_{pre}}ICU_{pre^*}^{i,j,k}(t) \\ &\quad + \gamma_U(t)ICU_{pre}^{i,j,k}(t) \end{aligned} \tag{50}$$

$$\begin{aligned} \frac{dICU_{W_R}^{i,j,k}(t)}{dt} &= \left(1-p_{ICU_D}^{i,j,k}(t)\right)\left(1-p_{W_D}^{i,j,k}(t)\right)\gamma_{ICU_{pre}}ICU_{pre}^{i,j,k}(t) \\ &\quad - \left(\gamma_{ICU_{W_R}} + \gamma_U(t)\right)ICU_{W_R}^{i,j,k}(t) \end{aligned} \tag{51}$$

$$\begin{aligned}\frac{dICU_{W_{R^*}}^{i,j,k}(t)}{dt} &= \left(1-p_{ICU_D}^{i,j,k}(t)\right)\left(1-p_{W_D}^{i,j,k}(t)\right)\gamma_{ICU_{pre}}ICU_{pre^*}^{i,j,k}(t)-\gamma_{ICU_{W_R}}ICU_{W_{R^*}}^{i,j,k}(t) \\ &\quad +\gamma_U(t)ICU_{W_R}^{i,j,k}(t)\end{aligned} \tag{52}$$

$$\frac{dICU_{W_D}^{i,j,k}(t)}{dt} = \left(1-p_{ICU_D}^{i,j,k}(t)\right)p_{W_D}^{i,j,k}(t)\gamma_{ICU_{pre}}ICU_{pre}^{i,j,k}(t)-\left(\gamma_{ICU_{W_D}}+\gamma_U(t)\right)ICU_{W_D}^{i,j,k}(t) \tag{53}$$

$$\begin{aligned}\frac{dICU_{W_{D^*}}^{i,j,k}(t)}{dt} &= \left(1-p_{ICU_D}^{i,j,k}(t)\right)p_{W_D}^{i,j,k}(t)\gamma_{ICU_{pre}}ICU_{pre^*}^{i,j,k}(t)-\gamma_{ICU_{W_D}}ICU_{W_{D^*}}^{i,j,k}(t) \\ &\quad +\gamma_U(t)ICU_{W_D}^{i,j,k}(t)\end{aligned} \tag{54}$$

$$\frac{dICU_D^{i,j,k,1}(t)}{dt} = p_{ICU_D}^{i,j,k}(t)\gamma_{ICU_{pre}}ICU_{pre}^{i,j,k}(t)-\left(\gamma_{ICU_D}+\gamma_U(t)\right)ICU_D^{i,j,k,1}(t) \tag{55}$$

$$\frac{dICU_D^{i,j,k,2}(t)}{dt} = \gamma_{ICU_D}ICU_D^{i,j,k,1}(t)-\left(\gamma_{ICU_D}+\gamma_U(t)\right)ICU_D^{i,j,k,2}(t) \tag{56}$$

$$\frac{dICU_{D^*}^{i,j,k,1}(t)}{dt} = p_{ICU_D}^{i,j,k}(t)\gamma_{ICU_{pre}}ICU_{pre^*}^{i,j,k}(t)-\gamma_{ICU_D}ICU_{D^*}^{i,j,k,1}(t)+\gamma_U(t)ICU_D^{i,j,k,1}(t) \tag{57}$$

$$\frac{dICU_{D^*}^{i,j,k,2}(t)}{dt} = \gamma_{ICU_D}ICU_{D^*}^{i,j,k,1}(t)-\gamma_{ICU_D}ICU_{D^*}^{i,j,k,2}(t)+\gamma_U(t)ICU_D^{i,j,k,2}(t) \tag{58}$$

$$\frac{dW_R^{i,j,k,1}(t)}{dt} = \gamma_{ICU_{W_R}}ICU_{W_R}^{i,j,k}(t)-\left(\gamma_{W_R}(t)+\gamma_U(t)\right)W_R^{i,j,k,1}(t) \tag{59}$$

$$\frac{dW_R^{i,j,k,2}(t)}{dt} = \gamma_{W_R}(t)W_R^{i,j,k,1}(t)-\left(\gamma_{W_R}(t)+\gamma_U(t)\right)W_R^{i,j,k,2}(t) \tag{60}$$

$$\frac{dW_{R^*}^{i,j,k,1}(t)}{dt} = \gamma_{ICU_{W_R}}ICU_{W_{R^*}}^{i,j,k}(t)-\gamma_{W_R}(t)W_{R^*}^{i,j,k,1}(t)+\gamma_U(t)W_R^{i,j,k,1}(t) \tag{61}$$

$$\frac{dW_{R^*}^{i,j,k,2}(t)}{dt} = \gamma_{W_R}(t)W_{R^*}^{i,j,k,1}(t)-\gamma_{W_R}(t)W_{R^*}^{i,j,k,2}(t)+\gamma_U(t)W_R^{i,j,k,2}(t) \tag{62}$$

$$\frac{dW_D^{i,j,k}(t)}{dt} = \gamma_{ICU_{W_D}}ICU_{W_D}^{i,j,k}(t)-\left(\gamma_{W_D}(t)+\gamma_U(t)\right)W_D^{i,j,k}(t) \tag{63}$$

$$\frac{dW_{D^*}^{i,j,k}(t)}{dt} = \gamma_{ICU_{W_D}}ICU_{W_{D^*}}^{i,j,k}(t)-\gamma_{W_D}(t)W_{D^*}^{i,j,k}(t)+\gamma_U(t)W_D^{i,j,k}(t) \tag{64}$$

$$\begin{aligned}\frac{dH_R^{i,j,k}(t)}{dt} &= p_H^{i,j,k}(t)\left(1-p_{G_D}^{i,j,k}(t)\right)\left(1-p^*(t)\right)\left(1-p_{ICU}^{i,j}(t)\right)\left(1-p_{H_D}^{i,j,k}(t)\right)\gamma_{C_2}^jI_{C_2}^{i,j,k}(t) \\ &\quad -\left(\gamma_{H_R}(t)+\gamma_U(t)\right)H_R^{i,j,k}(t)\end{aligned} \tag{65}$$

$$\begin{aligned}\frac{dH_{R^*}^{i,j,k}(t)}{dt} &= p_H^{i,j,k}(t)\left(1-p_{G_D}^{i,j,k}(t)\right)p^*(t)\left(1-p_{ICU}^{i,j}(t)\right)\left(1-p_{H_D}^{i,j,k}(t)\right)\gamma_{C_2}^jI_{C_2}^{i,j,k}(t) \\ &\quad +\gamma_U(t)H_R^{i,j,k}(t)-\gamma_{H_R}(t)H_{R^*}^{i,j,k}(t)\end{aligned} \tag{66}$$

$$\begin{aligned}\frac{dH_D^{i,j,k,1}(t)}{dt} &= p_H^{i,j,k}(t)\left(1-p_{G_D}^{i,j,k}(t)\right)\left(1-p^*(t)\right)\left(1-p_{ICU}^{i,j}(t)\right)p_{H_D}^{i,j,k}(t)\gamma_{C_2}^jI_{C_2}^{i,j,k}(t) \\ &\quad -\left(\gamma_{H_D}(t)+\gamma_U(t)\right)H_D^{i,j,k,1}(t)\end{aligned} \tag{67}$$

$$\frac{dH_D^{i,j,k,2}(t)}{dt} = \gamma_{H_D}(t)H_D^{i,j,k,1}(t)-\left(\gamma_{H_D}(t)+\gamma_U(t)\right)H_D^{i,j,k,2}(t) \tag{68}$$

$$\begin{aligned}\frac{dH_{D^*}^{i,j,k,1}(t)}{dt} &= p_H^{i,j,k}(t)\left(1-p_{G_D}^{i,j,k}(t)\right)p^*(t)\left(1-p_{ICU}^{i,j}(t)\right)\ p_{H_D}^{i,j,k}(t)\gamma_{C_2}^jI_{C_2}^{i,j,k}(t)+\gamma_U(t)H_D^{i,j,k,1}(t) \\ &\quad -\gamma_{H_D}(t)H_{D^*}^{i,j,k,1}(t)\end{aligned} \tag{69}$$

$$\frac{dH_{D^*}^{i,j,k,2}(t)}{dt} = \gamma_{H_D}(t)H_{D^*}^{i,j,k,1}(t)-\gamma_{H_D}(t)H_{D^*}^{i,j,k,2}(t)+\gamma_U(t)H_D^{i,j,k,2}(t) \tag{70}$$

$$
\begin{aligned}
\frac{dR^{i,j,k}(t)}{dt} = & \mathbb{1}_{\left\{j_1,j_2,j_1^{reinf},j_2^{reinf}\right\}}(j)\left(\gamma_A^j I_A^{i,j,k}(t) + \left(1-p_H^{i,j,k}(t)\right)\gamma_{C_2}^j I_{C_2}^{i,j,k}(t) + \gamma_{H_R}(t)\left(H_R^{i,j,k}(t)+H_{R^*}^{i,j,k}(t)\right)\right. \\
& \left. +\gamma_{W_R}(t)\left(W_R^{i,j,k,2}(t)+W_{R^*}^{i,j,k,2}(t)\right)\right) \\
& - \mathbb{1}_{\{j_H\}}(j)\left(1-\eta_{j_1}\right)\lambda^{i,j_1,k}(t)R^{i,j,k}(t) - \mathbb{1}_{\left\{j_1,j_1^{reinf},j_H\right\}}(j)\left(1-\eta_{j_2}\right)\lambda^{i,j_2,k}(t)R^{i,j,k}(t) \\
& + \zeta^{i,k-1}(t)R^{i,j,k-1}(t) + \mathbb{1}_{\{5\}}(k)\zeta^{i,4}(t)R^{i,j,3}(t) - \left(\gamma_R + \zeta^{i,k}(t) + \mathbb{1}_{\{3\}}(k)\zeta^{i,4}(t)\right)R^{i,j,k}(t)
\end{aligned} \tag{71}
$$

$$\frac{dT^i_{sero^1_{pre}}(t)}{dt} = -\gamma_{sero_{pre}}T^i_{sero^1_{pre}}(t) + \sum_j\sum_k \gamma_E^j E^{i,j,k,2}(t) \tag{72}$$

$$\frac{dT^i_{sero^1_{pos}}(t)}{dt} = p_{sero_{pos}}\gamma_{sero_{pre}}T^i_{sero^1_{pre}}(t) - \gamma_{sero^1_{pos}}T^i_{sero^1_{pos}}(t) \tag{73}$$

$$\frac{dT^i_{sero^1_{neg}}(t)}{dt} = \left(1-p_{sero_{pos}}\right)\gamma_{sero_{pre}}T^i_{sero^1_{pre}}(t) + \gamma_{sero^1_{pos}}T^i_{sero^1_{pos}}(t) \tag{74}$$

$$\frac{dT^i_{sero^2_{pre}}(t)}{dt} = -\gamma_{sero_{pre}}T^i_{sero^2_{pre}}(t) + \sum_j\sum_k \gamma_E^j E^{i,j,k,2}(t) \tag{75}$$

$$\frac{dT^i_{sero^2_{pos}}(t)}{dt} = p_{sero_{pos}}\gamma_{sero_{pre}}T^i_{sero^2_{pre}}(t) - \gamma_{sero^2_{pos}}T^i_{sero^2_{pos}}(t) \tag{76}$$

$$\frac{dT^i_{sero^2_{neg}}(t)}{dt} = \left(1-p_{sero_{pos}}\right)\gamma_{sero_{pre}}T^i_{sero^2_{pre}}(t) + \gamma_{sero^2_{pos}}T^i_{sero^2_{pos}}(t) \tag{77}$$

$$\frac{dT^i_{PCR_{pre}}(t)}{dt} = -\gamma_{PCR_{pre}}T^i_{PCR_{pre}}(t) + \sum_k\left(\lambda^{i,Alpha,k}(t) + \lambda^{i,Delta,k}(t)\right)S^{i,k}(t) \tag{78}$$

$$\frac{dT^i_{PCR_{pos}}(t)}{dt} = \gamma_{PCR_{pre}}T^i_{PCR_{pre}}(t) - \gamma_{PCR_{pos}}T^i_{PCR_{pos}}(t) \tag{79}$$

$$\frac{dT^i_{PCR_{neg}}(t)}{dt} = \gamma_{PCR_{pos}}T^i_{PCR_{pos}}(t). \tag{80}$$

We used the tau-leap method [59] to create a stochastic, time-discretised version of the model described in equations (83) - (223), taking four update steps per day ($dt = 0.25$ days).

For each time step, the model iterated through the procedure described below. In the following, we introduce a small abuse of notation: for transitions involving multiple onward compartments (e.g. transition from compartment $E$ to compartments $I_A$ or $I_P$ or to the next vaccination strata within $E$), for conciseness, we write

$$\left(d^{i,j,k}_{E,I_A},\ d^{i,j,k}_{E,I_P},\ d^{i,j,k}_{E,v}\right) \sim \text{Multinom}\left(E^{i,j,k,2}(t),\ q^{i,j,k}_{E,I_A},\ q^{i,j,k}_{E,I_P},\ q^{i,j,k}_{E,v}\right) \tag{81}$$

instead of

$$\left(d^{i,j,k}_{E,I_A},\ d^{i,j,k}_{E,I_P},\ d^{i,j,k}_{E,v},\ d^{i,j,k}_{nomove}\right) \sim \text{Multinom}\left(E^{i,j,k,2}(t),\ q^{i,j,k}_{E,I_A},\ q^{i,j,k}_{E,I_P},\ q^{i,j,k}_{E,v},\ 1-\sum_{x\in I_A,I_P,v} q^{i,j,k}_{E,x}\right) \tag{82}$$

where $d^{i,j,k}_{nomove}$ is a dummy variable counting the number of individuals remaining in compartment $E^{i,j,k,2}$. We also omit the time dependency i.e. we use $d^{i,j,k}_{E,I_A}$ or $q^{i,j,k}_{E,I_A}$ instead of $d^{i,j,k}_{E,I_A}(t)$ or $q^{i,j,k}_{E,I_A}(t)$.

Using this convention, transition variables are drawn from the following distributions, with probabilities defined below:

$$q^{i,j_1,k}_{S,E} = \left(1-e^{-\Lambda^{i,k}(t)dt}\right)\frac{\lambda^{i,j_1,k}(t)}{\Lambda^{i,k}(t)} \tag{83}$$

$$q^{i,j_2,k}_{S,E} = \left(1-e^{-\Lambda^{i,k}(t)dt}\right)\frac{\lambda^{i,j_2,k}(t)}{\Lambda^{i,k}(t)} \tag{84}$$

$$\left(q^{i,k}_{S,v}, q^{i,k}_{S,v,2}\right) = \left(1-e^{-\zeta^{i,k}(t)dt}, \mathbb{1}_{\{3\}}(k)\left(1-e^{-\zeta^{i,4}(t)dt}\right)e^{-\zeta^{i,k}(t)dt}\right) \tag{85}$$

$$\left(d^{i,j_1,k}_{S,E},\ d^{i,j_2,k}_{S,E}\right) \sim \text{Multinom}\left(S^{i,k}(t),\ q^{i,j_1,k}_{S,E},\ q^{i,j_2,k}_{S,E}\right) \tag{86}$$

$$d_{seed}^{i,j,k} \sim \min\left(\text{Poisson}\left(\mathbb{1}_{\{j_1,j_2\}}(j)\hat{\delta}^{i,j,k}(t)dt\right),\ S^{i,k}(t) - d_{S,E}^{i,j_1,k} - d_{S,E}^{i,j_2,k}\right) \tag{87}$$

$$\left(d_{S,v}^{i,k}, d_{S,v,2}^{i,k}\right) \sim \text{Multinom}\left(S^{i,k}(t) - d_{seed}^{i,j,k} - d_{S,E}^{i,j_1,k} - d_{S,E}^{i,j_2,k},\ q_{S,v}^{i,k},\ q_{S,v,2}^{i,k}\right) \tag{88}$$

$$\left(q_{E,E}^{i,j,k},\ q_{E,v}^{i,j,k,1}\right) = \left(\ 1 - e^{-\gamma_E^j dt},\ e^{-\gamma_E^j dt}\left(1 - e^{-\zeta^{i,k}(t)dt}\right)\right) \tag{89}$$

$$q_{E,v,2}^{i,j,k,1} = \left(1 - q_{E,v}^{i,j,k,1}\right)\mathbb{1}_{\{3\}}(k)\left(1 - e^{-\zeta^{i,4}(t)dt}\right) \tag{90}$$

$$\left(d_{E,E}^{i,j,k},\ d_{E,v}^{i,j,k,1},\ d_{E,v,2}^{i,j,k,1}\right) \sim \text{Multinom}\left(E^{i,j,k,1}(t),\ q_{E,E}^{i,j,k},\ q_{E,v}^{i,j,k,1},\ q_{E,v,2}^{i,j,k,1}\right) \tag{91}$$

$$q_{E,I_A}^{i,j,k} = \left(1 - p_C^i\right)\left(1 - e^{-\gamma_E^j dt}\right) \tag{92}$$

$$q_{E,I_P}^{i,j,k} = p_C^i\left(1 - e^{-\gamma_E^j dt}\right) \tag{93}$$

$$q_{E,v}^{i,j,k,2} = e^{-\gamma_E^j dt}\left(1 - e^{-\zeta^{i,k}(t)dt}\right) \tag{94}$$

$$q_{E,v,2}^{i,j,k,2} = \left(1 - q_{E,v}^{i,j,k,2}\right)\mathbb{1}_{\{3\}}(k)\left(1 - e^{-\zeta^{i,4}(t)dt}\right) \tag{95}$$

$$\left(d_{E,I_A}^{i},\ d_{E,I_P}^{i},\ d_{E,v}^{i,j,k,2},\ d_{E,v,2}^{i,j,k,2}\right) \sim \text{Multinom}\left(E^{i,j,k,2}(t),\ q_{E,I_A}^{i,j,k},\ q_{E,I_P}^{i,j,k},\ q_{E,v}^{i,j,k,2},\ q_{E,v,2}^{i,j,k,2}\right) \tag{96}$$

$$\left(q_{I_A,R}^{i,j,k},\ q_{I_A,v}^{i,j,k}\right) = \left(1 - e^{-\gamma_A^j dt},\ e^{-\gamma_A^j dt}\left(1 - e^{-\zeta^{i,k}(t)dt}\right)\right) \tag{97}$$

$$q_{I_A,v,2}^{i,j,k} = \left(1 - q_{I_A,v}^{i,j,k}\right)\mathbb{1}_{\{3\}}(k)\left(1 - e^{-\zeta^{i,4}(t)dt}\right) \tag{98}$$

$$\left(d_{I_A,R}^{i,j,k},\ d_{I_A,v}^{i,j,k},\ d_{I_A,v,2}^{i,j,k}\right) \sim \text{Multinom}\left(I_A^i(t),\ q_{I_A,R}^{i,j,k},\ q_{I_A,v}^{i,j,k},\ q_{I_A,v,2}^{i,j,k}\right) \tag{99}$$

$$\left(q_{I_P,I_{C_1}}^{i,j,k},\ q_{I_P,v}^{i,j,k}\right) = \left(1 - e^{-\gamma_P^j dt},\ e^{-\gamma_P^j dt}\left(1 - e^{-\zeta^{i,k}(t)dt}\right)\right) \tag{100}$$

$$q_{I_P,v,2}^{i,j,k} = \left(1 - q_{I_P,v}^{i,j,k}\right)\mathbb{1}_{\{3\}}(k)\left(1 - e^{-\zeta^{i,4}(t)dt}\right) \tag{101}$$

$$\left(d_{I_P,I_{C_1}}^{i,j,k},\ d_{I_P,v}^{i,j,k},\ d_{I_P,v,2}^{i,j,k}\right) \sim \text{Multinom}\left(I_P^i(t),\ q_{I_P,I_{C_1}}^{i,j,k},\ q_{I_P,v}^{i,j,k},\ q_{I_P,v,2}^{i,j,k}\right) \tag{102}$$

$$d_{I_{C_1},I_{C_2}}^{i,j,k} \sim \text{Binom}\left(I_{C_1}^{i,j,k}(t),\ 1 - e^{-\gamma_{C_1}^j dt}\right) \tag{103}$$

$$q_{I_{C_2},G_D}^{i,j,k} = p_H^{i,j,k}(t)p_{G_D}^{i,j,k}(t)\left(1 - e^{-\gamma_{C_2}^j dt}\right) \tag{104}$$

$$q_{I_{C_2},R}^{i,j,k} = \left(1 - p_H^{i,j,k}(t)\right)\left(1 - e^{-\gamma_{C_2}^j dt}\right) \tag{105}$$

$$q_{I_{C_2},ICU_{pre}}^{i,j,k} = p_H^{i,j,k}(t)\left(1 - p_{G_D}^{i,j,k}(t)\right)\left(1 - p^*(t)\right)p_{ICU}^{i,j}(t)\left(1 - e^{-\gamma_{C_2}^j dt}\right) \tag{106}$$

$$q_{I_{C_2},ICU_{pre^*}}^{i,j,k} = p_H^{i,j,k}(t)\left(1 - p_{G_D}^{i,j,k}(t)\right)p^*(t)p_{ICU}^{i,j}(t)\left(1 - e^{-\gamma_{C_2}^j dt}\right) \tag{107}$$

$$q_{I_{C_2},H_R}^{i,j,k} = p_H^{i,j,k}(t)\left(1 - p_{G_D}^{i,j,k}(t)\right)\left(1 - p^*(t)\right)\left(1 - p_{ICU}^{i,j}(t)\right)\left(1 - p_{H_D}^{i,j,k}(t)\right)\left(1 - e^{-\gamma_{C_2}^j dt}\right) \tag{108}$$

$$q_{I_{C_2},H_{R^*}}^{i,j,k} = p_H^{i,j,k}(t)\left(1 - p_{G_D}^{i,j,k}(t)\right)\ p^*(t)\left(1 - p_{ICU}^{i,j}(t)\right)\left(1 - p_{H_D}^{i,j,k}(t)\right)\left(1 - e^{-\gamma_{C_2}^j dt}\right) \tag{109}$$

$$q_{I_{C_2},H_D}^{i,j,k} = p_H^{i,j,k}(t)\left(1 - p_{G_D}^{i,j,k}(t)\right)\left(1 - p^*(t)\right)\left(1 - p_{ICU}^{i,j}(t)\right)p_{H_D}^{i,j,k}(t)\left(1 - e^{-\gamma_{C_2}^j dt}\right) \tag{110}$$

$$q_{I_{C_2},H_{D^*}}^{i,j,k} = p_H^{i,j,k}(t)\left(1 - p_{G_D}^{i,j,k}(t)\right)p^*(t)\left(1 - p_{ICU}^{i,j}(t)\right)p_{H_D}^{i,j,k}(t)\left(1 - e^{-\gamma_{C_2}^j dt}\right) \tag{111}$$

$$\left(d_{I_{C_2},G_D}^{i,j,k}, \ldots, d_{I_{C_2},H_{D^*}}^{i,j,k}\right) \sim \text{Multinom}\left(I_{C_2}^{i,j,k}(t),\ q_{I_{C_2},G_D}^{i,j,k}, \ldots, q_{I_{C_2},H_{D^*}}^{i,j,k}\right) \tag{112}$$

$$d_{G_D,G_D}^{i,j,k} \sim \text{Binom}\left(G_D^{i,j,k,1}(t),\ 1 - e^{-\gamma_{G_D} dt}\right) \tag{113}$$

$$d_{G_D,D}^{i,j,k} \sim \text{Binom}\left(G_D^{i,j,k,2}(t),\ 1 - e^{-\gamma_{G_D} dt}\right) \tag{114}$$

$$q_{ICU_{pre},ICU_{W_R}}^{i,j,k} = \left(1 - p_{ICU_D}^{i,j,k}(t)\right)\left(1 - p_{W_D}^{i,j,k}(t)\right)\left(1 - e^{-\gamma_{ICU_{pre}} dt}\right)e^{-\gamma_U(t)dt} \tag{115}$$

$$q_{ICU_{pre},ICU_{W_{R^*}}}^{i,j,k} = \left(1 - p_{ICU_D}^{i,j,k}(t)\right)\left(1 - p_{W_D}^{i,j,k}(t)\right)\left(1 - e^{-\gamma_{ICU_{pre}} dt}\right)\left(1 - e^{-\gamma_U(t)dt}\right) \tag{116}$$

$$q^{i,j,k}_{ICU_{pre},ICU_{W_D}} = \left(1-p^{i,j,k}_{ICU_D}(t)\right) p^{i,j,k}_{W_D}(t) \left(1-e^{-\gamma_{ICU_{pre}} dt}\right) e^{-\gamma_U(t)dt} \tag{117}$$

$$q^{i,j,k}_{ICU_{pre},ICU_{W_{D^*}}} = \left(1-p^{i,j,k}_{ICU_D}(t)\right) p^{i,j,k}_{W_D}(t) \left(1-e^{-\gamma_{ICU_{pre}} dt}\right) \left(1-e^{-\gamma_U(t)dt}\right) \tag{118}$$

$$q^{i,j,k}_{ICU_{pre},ICU_D} = p^{i,j,k}_{ICU_D}(t) \left(1-e^{-\gamma_{ICU_{pre}} dt}\right) e^{-\gamma_U(t)dt} \tag{119}$$

$$q^{i,j,k}_{ICU_{pre},ICU_{D^*}} = p^{i,j,k}_{ICU_D}(t) \left(1-e^{-\gamma_{ICU_{pre}} dt}\right) \left(1-e^{-\gamma_U(t)dt}\right) \tag{120}$$

$$q^{i,j,k}_{ICU_{pre},ICU_{pre^*}} = e^{-\gamma_{ICU_{pre}} dt} \left(1-e^{-\gamma_U(t)dt}\right) \tag{121}$$

$$\begin{aligned}\left(d^{i,j,k}_{ICU_{pre},ICU_{W_R}}, \ldots, d^{i,j,k}_{ICU_{pre},ICU_{pre^*}}\right) \\ \sim \text{Multinom}\left(ICU^{i,j,k}_{pre}(t),\ q^{i,j,k}_{ICU_{pre},ICU_{W_R}}, \ldots, q^{i,j,k}_{ICU_{pre},ICU_{pre^*}}\right)\end{aligned} \tag{122}$$

$$q^{i,j,k}_{ICU_{pre^*},ICU_{W_{R^*}}} = \left(1-p^{i,j,k}_{ICU_D}(t)\right) \left(1-p^{i,j,k}_{W_D}(t)\right) \left(1-e^{-\gamma_{ICU_{pre}} dt}\right) \tag{123}$$

$$q^{i,j,k}_{ICU_{pre^*},ICU_{W_{D^*}}} = \left(1-p^{i,j,k}_{ICU_D}(t)\right) p^{i,j,k}_{W_D}(t) \left(1-e^{-\gamma_{ICU_{pre}} dt}\right) \tag{124}$$

$$q^{i,j,k}_{ICU_{pre^*},ICU_{D^*}} = p^{i,j,k}_{ICU_D}(t) \left(1-e^{-\gamma_{ICU_{pre}} dt}\right) \tag{125}$$

$$\begin{aligned}\left(d^{i,j,k}_{ICU_{pre^*},ICU_{W_{R^*}}}, \ldots, d^{i,j,k}_{ICU_{pre^*},ICU_{D^*}}\right) \\ \sim \text{Multinom}\left(ICU^{i,j,k}_{pre^*}(t),\ q^{i,j,k}_{ICU_{pre^*},ICU_{W_{R^*}}}, \ldots, q^{i,j,k}_{ICU_{pre^*},ICU_{D^*}}\right)\end{aligned} \tag{126}$$

$$q^{i,j,k}_{H_D,H_D} = \left(1-e^{-\gamma_{H_D}(t)dt}\right) e^{-\gamma_U(t)dt} \tag{127}$$

$$q^{i,j,k,1,1}_{H_D,H_{D^*}} = e^{-\gamma_{H_D}(t)dt} \left(1-e^{-\gamma_U(t)dt}\right) \tag{128}$$

$$q^{i,j,k,1,2}_{H_D,H_{D^*}} = \left(1-e^{-\gamma_{H_D}(t)dt}\right) \left(1-e^{-\gamma_U(t)dt}\right) \tag{129}$$

$$\begin{aligned}\left(d^{i,j,k}_{H_D,H_D},\ d^{i,j,k,1,1}_{H_D,H_{D^*}},\ d^{i,j,k,1,2}_{H_D,H_{D^*}}\right) \\ \sim \text{Multinom}\left(H^{i,j,k,1}_D(t),\ q^{i,j,k}_{H_D,H_D},\ q^{i,j,k,1,1}_{H_D,H_{D^*}},\ q^{i,j,k,1,2}_{H_D,H_{D^*}}\right)\end{aligned} \tag{130}$$

$$d^{i,j,k}_{H_{D^*},H_{D^*}} \sim \text{Binom}\left(H^{i,j,k,1}_{D^*}(t),\ 1-e^{-\gamma_{H_D}(t)dt}\right) \tag{131}$$

$$\left(d^{i,j,k}_{H_D,D},\ d^{i,j,k,2,2}_{H_D,H_{D^*}}\right) \sim \text{Multinom}\left(H^{i,j,k,2}_D(t),\ 1-e^{-\gamma_{H_D}(t)dt},\ e^{-\gamma_{H_D}(t)dt}\left(1-e^{-\gamma_U(t)dt}\right)\right) \tag{132}$$

$$d^{i,j,k}_{H_{D^*},D} \sim \text{Binom}\left(H^{i,j,k,2}_{D^*}(t),\ 1-e^{-\gamma_{H_D}(t)dt}\right) \tag{133}$$

$$\left(d^{i,j,k}_{H_R,R},\ d^{i,j,k}_{H_R,H_{R^*}}\right) \sim \text{Multinom}\left(H^{i,j,k}_R(t),\ 1-e^{-\gamma_{H_R}(t)dt},\ e^{-\gamma_{H_R}(t)dt}\left(1-e^{-\gamma_U(t)dt}\right)\right) \tag{134}$$

$$d^{i,j,k}_{H_{R^*},R} \sim \text{Binom}\left(H^{i,j,k}_{R^*}(t),\ 1-e^{-\gamma_{H_R}(t)dt}\right) \tag{135}$$

$$q^{i,j,k}_{ICU_{W_R},W_R} = \left(1-e^{-\gamma_{ICU_{W_R}} dt}\right) e^{-\gamma_U(t)dt} \tag{136}$$

$$q^{i,j,k}_{ICU_{W_R},ICU_{W_{R^*}}} = e^{-\gamma_{ICU_{W_R}} dt} \left(1-e^{-\gamma_U(t)dt}\right) \tag{137}$$

$$q^{i,j,k}_{ICU_{W_R},W_{R^*}} = \left(1-e^{-\gamma_{ICU_{W_R}} dt}\right) \left(1-e^{-\gamma_U(t)dt}\right) \tag{138}$$

$$\begin{aligned}\left(d^{i,j,k}_{ICU_{W_R},W_R}, \ldots, d^{i,j,k}_{ICU_{W_R},W_{R^*}}\right) \\ \sim \text{Multinom}\left(ICU^{i,j,k}_{W_R}(t),\ q^{i,j,k}_{ICU_{W_R},W_R}, \ldots, q^{i,j,k}_{ICU_{W_R},W_{R^*}}\right)\end{aligned} \tag{139}$$

$$d^{i,j,k}_{ICU_{W_{R^*}},W_{R^*}} \sim \text{Binom}\left(ICU^{i,j,k}_{W_{R^*}}(t),\ 1-e^{-\gamma_{ICU_{W_R}} dt}\right) \tag{140}$$

$$q^{i,j,k}_{ICU_{W_D},W_D} = \left(1-e^{-\gamma_{ICU_{W_D}} dt}\right) e^{-\gamma_U(t)dt} \tag{141}$$

$$q^{i,j,k}_{ICU_{W_D},ICU_{W_{D^*}}} = e^{-\gamma_{ICU_{W_D}} dt} \left(1-e^{-\gamma_U(t)dt}\right) \tag{142}$$

$$q^{i,j,k}_{ICU_{W_D},W_{D^*}} = \left(1-e^{-\gamma_{ICU_{W_D}} dt}\right) \left(1-e^{-\gamma_U(t)dt}\right) \tag{143}$$

$$\begin{aligned}\left(d^{i,j,k}_{ICU_{W_D},W_D}, \ldots, d^{i,j,k}_{ICU_{W_D},W_{D^*}}\right) \\ \sim \text{Multinom}\left(ICU^{i,j,k}_{W_D}(t),\ q^{i,j,k}_{ICU_{W_D},W_D}, \ldots, q^{i,j,k}_{ICU_{W_D},W_{D^*}}\right)\end{aligned} \tag{144}$$

$$d^{i,j,k}_{ICU_{W_{D^*}},W_{D^*}} \sim \text{Binom}\left(ICU^{i,j,k}_{W_{D^*}}(t),\ 1-e^{-\gamma_{ICU_{W_D}}dt}\right) \tag{145}$$

$$q^{i,j,k}_{ICU_D,ICU_D} = \left(1-e^{-\gamma_{ICU_D}dt}\right)e^{-\gamma_U(t)dt} \tag{146}$$

$$q^{i,j,k,1,1}_{ICU_D,ICU_{D^*}} = e^{-\gamma_{ICU_D}dt}\left(1-e^{-\gamma_U(t)dt}\right) \tag{147}$$

$$q^{i,j,k,1,2}_{ICU_D,ICU_{D^*}} = \left(1-e^{-\gamma_{ICU_D}dt}\right)\left(1-e^{-\gamma_U(t)dt}\right) \tag{148}$$

$$\begin{aligned}&\left(d^{i,j,k}_{ICU_D,ICU_D},\ d^{i,j,k,1,1}_{ICU_D,ICU_{D^*}},\ d^{i,j,k,1,2}_{ICU_D,ICU_{D^*}}\right)\\ &\quad\sim \text{Multinom}\left(ICU^{i,j,k,1}_D(t),\ q^{i,j,k}_{ICU_D,ICU_D},\ q^{i,j,k,1,1}_{ICU_D,ICU_{D^*}},\ q^{i,j,k,1,2}_{ICU_D,ICU_{D^*}}\right)\end{aligned} \tag{149}$$

$$d^{i,j,k}_{ICU_{D^*},ICU_{D^*}} \sim \text{Binom}\left(ICU^{i,j,k,1}_{D^*}(t),\ 1-e^{-\gamma_{ICU_D}dt}\right) \tag{150}$$

$$\left(q^{i,j,k}_{ICU_D,D},\ q^{i,j,k,2,2}_{ICU_D,ICU_{D^*}}\right) = \left(1-e^{-\gamma_{ICU_D}dt},\ e^{-\gamma_{ICU_D}dt}\left(1-e^{-\gamma_U(t)dt}\right)\right) \tag{151}$$

$$\left(d^{i,j,k}_{ICU_D,D},\ d^{i,j,k,2,2}_{ICU_D,ICU_{D^*}}\right) \sim \text{Multinom}\left(ICU^{i,j,k,2}_D(t),\ q^{i,j,k}_{ICU_D,D},\ q^{i,j,k,2,2}_{ICU_D,ICU_{D^*}}\right) \tag{152}$$

$$d^{i,j,k}_{ICU_{D^*},D} \sim \text{Binom}\left(ICU^{i,j,k,2}_{D^*}(t),\ 1-e^{-\gamma_{ICU_D}dt}\right) \tag{153}$$

$$q^{i,j,k}_{W_R,W_R} = \left(1-e^{-\gamma_{W_R}(t)dt}\right)e^{-\gamma_U(t)dt} \tag{154}$$

$$q^{i,j,k,1,1}_{W_R,W_{R^*}} = e^{-\gamma_{W_R}(t)dt}\left(1-e^{-\gamma_U(t)dt}\right) \tag{155}$$

$$q^{i,j,k,1,2}_{W_R,W_{R^*}} = \left(1-e^{-\gamma_{W_R}(t)dt}\right)\left(1-e^{-\gamma_U(t)dt}\right) \tag{156}$$

$$\begin{aligned}&\left(d^{i,j,k}_{W_R,W_R},\ d^{i,j,k,1,1}_{W_R,W_{R^*}},\ d^{i,j,k,1,2}_{W_R,W_{R^*}}\right)\\ &\quad\sim\ \text{Multinom}\left(W^{i,j,k,1}_R(t),\ q^{i,j,k}_{W_R,W_R},\ q^{i,j,k,1,1}_{W_R,W_{R^*}},\ q^{i,j,k,1,2}_{W_R,W_{R^*}}\right)\end{aligned} \tag{157}$$

$$d^{i,j,k}_{W_{R^*},W_{R^*}} \sim \text{Binom}\left(W^{i,j,k,1}_{R^*}(t),\ 1-e^{-\gamma_{W_R}(t)dt}\right) \tag{158}$$

$$\left(q^{i,j,k}_{W_R,R},\ q^{i,j,k,2,2}_{W_R,W_{R^*}}\right) = \left(1-e^{-\gamma_{W_R}(t)dt},\ e^{-\gamma_{W_R}(t)dt}\left(1-e^{-\gamma_U(t)\ dt}\right)\right) \tag{159}$$

$$\left(d^{i,j,k}_{W_R,R},\ d^{i,j,k,2,2}_{W_R,W_{R^*}}\right) \sim \text{Multinom}\left(W^{i,j,k,2}_R(t),\ q^{i,j,k}_{W_R,R},\ q^{i,j,k,2,2}_{W_R,W_{R^*}}\right) \tag{160}$$

$$d^{i,j,k}_{W_{R^*},R} \sim \text{Binom}\left(W^{i,j,k,2}_{R^*}(t),\ 1-e^{-\gamma_{W_R}(t)dt}\right) \tag{161}$$

$$\left(q^{i,j,k}_{W_D,D},\ q^{i,j,k}_{W_D,W_{D^*}}\right) = \left(1-e^{-\gamma_{W_D}(t)dt},\ e^{-\gamma_{W_D}(t)dt}\left(1-e^{-\gamma_U(t)dt}\right)\right) \tag{162}$$

$$\left(d^{i,j,k}_{W_D,D},\ d^{i,j,k}_{W_D,W_{D^*}}\right) \sim \text{Multinom}\left(W^{i,j,k}_D(t),\ q^{i,j,k}_{W_D,D},\ q^{i,j,k}_{W_D,W_{D^*}}\right) \tag{163}$$

$$d^{i,j,k}_{W_{D^*},D} \sim \text{Binom}\left(W^{i,j,k}_{D^*}(t),\ 1-e^{-\gamma_{W_D}(t)dt}\right) \tag{164}$$

$$\gamma^{i,j,k,j_1}_{R,E} = \mathbb{1}_{\{j_H\}}(j)(1-\eta_{j_1})\lambda^{i,j_1,k}(t) \tag{165}$$

$$\gamma^{i,j,k,j_2}_{R,E} = \mathbb{1}_{\left\{j_1,j_1^{reinf},j_H\right\}}(j)(1-\eta_{j_2})\lambda^{i,j_2,k}(t) \tag{166}$$

$$q^{i,j,k}_{R,S} = \left(1-e^{-\left(\gamma_R+\gamma^{i,j,k,j_1}_{R,E}+\gamma^{i,j,k,j_2}_{R,E}\right)dt}\right)\frac{\gamma_R}{\gamma_R+\gamma^{i,j,k,j_1}_{R,E}+\gamma^{i,j,k,j_2}_{R,E}} \tag{167}$$

$$q^{i,j,k,j_1}_{R,E} = \left(1-e^{-\left(\gamma_R+\gamma^{i,j,k,j_1}_{R,E}+\gamma^{i,j,k,j_2}_{R,E}\right)dt}\right)\frac{\gamma^{i,j,k,j_1}_{R,E}}{\gamma_R+\gamma^{i,j,k,j_1}_{R,E}+\gamma^{i,j,k,j_2}_{R,E}} \tag{168}$$

$$q^{i,j,k,j_2}_{R,E} = \left(1-e^{-\left(\gamma_R+\gamma^{i,j,k,j_1}_{R,E}+\gamma^{i,j,k,j_2}_{R,E}\right)dt}\right)\frac{\gamma^{i,j,k,j_2}_{R,E}}{\gamma_R+\gamma^{i,j,k,j_1}_{R,E}+\gamma^{i,j,k,j_2}_{R,E}} \tag{169}$$

$$q^{i,j,k}_{R,v} = e^{-\left(\gamma_R+\gamma^{i,j,k,j_1}_{R,E}+\gamma^{i,j,k,j_2}_{R,E}\right)dt}\left(1-e^{-\zeta^{i,k}(t)dt}\right) \tag{170}$$

$$q^{i,j,k}_{R,v,2} = \left(1-q^{i,j,k}_{R,v}\right)\mathbb{1}_{\{3\}}(k)\left(1-e^{-\zeta^{i,4}(t)dt}\right) \tag{171}$$

$$\begin{aligned}&\left(d^{i,j,k}_{R,S},\ d^{i,j,k,j_1}_{R,E},\ d^{i,j,k,j_2}_{R,E},\ d^{i,j,k}_{R,v},\ d^{i,j,k}_{R,v,2}\right)\\ &\quad\sim \text{Multinom}\left(R^{i,j,k}(t),\ q^{i,j,k}_{R,S},\ q^{i,j,k,j_1}_{R,E},\ q^{i,j,k,j_1}_{R,E},\ q^{i,j,k}_{R,v},\ q^{i,j,k}_{R,v,2}\right)\end{aligned} \tag{172}$$

$$q^i_{T_{sero^1_{pre}},T_{sero^1_{pos}}} = p_{sero_{pos}} \left(1 - e^{-\gamma_{sero_{pre}} dt}\right) \tag{173}$$

$$q^i_{T_{sero^1_{pre}},T_{sero^1_{neg}}} = \left(1 - p_{sero_{pos}}\right) \left(1 - e^{-\gamma_{sero_{pre}} dt}\right) \tag{174}$$

$$\begin{aligned}\left(d^i_{T_{sero^1_{pre}},T_{sero^1_{pos}}},\ d^i_{T_{sero^1_{pre}},T_{sero^1_{neg}}}\right)& \\ &\sim \text{Multinom}\left(T^i_{sero^1_{pre}}(t),\ q^i_{T_{sero^1_{pre}},T_{sero^1_{pos}}},\ q^i_{T_{sero^1_{pre}},T_{sero^1_{neg}}}\right)\end{aligned} \tag{175}$$

$$d^i_{T_{sero^1_{pos}},T_{sero^1_{neg}}} \sim \text{Binom}\left(T^i_{sero^1_{pos}}(t),\ 1 - e^{-\gamma_{sero^1_{pos}} dt}\right) \tag{176}$$

$$q^i_{T_{sero^2_{pre}},T_{sero^2_{pos}}} = p_{sero_{pos}} \left(1 - e^{-\gamma_{sero_{pre}} dt}\right) \tag{177}$$

$$q^i_{T_{sero^2_{pre}},T_{sero^2_{neg}}} = \left(1 - p_{sero_{pos}}\right) \left(1 - e^{-\gamma_{sero_{pre}} dt}\right) \tag{178}$$

$$\begin{aligned}\left(d^i_{T_{sero^2_{pre}},T_{sero^2_{pos}}},\ d^i_{T_{sero^2_{pre}},T_{sero^2_{neg}}}\right)& \\ &\sim \text{Multinom}\left(T^i_{sero^2_{pre}}(t),\ q^i_{T_{sero^2_{pre}},T_{sero^2_{pos}}},\ q^i_{T_{sero^2_{pre}},T_{sero^2_{neg}}}\right)\end{aligned} \tag{179}$$

$$d^i_{T_{sero^2_{pos}},T_{sero^2_{neg}}} \sim \text{Binom}\left(T^i_{sero^2_{pos}}(t),\ 1 - e^{-\gamma_{sero^2_{pos}} dt}\right) \tag{180}$$

$$d^i_{T_{PCR_{pre}},T_{PCR_{pos}}} \sim \text{Binom}\left(T^i_{PCR_{pre}}(t),\ 1 - e^{-\gamma_{PCR_{pre}} dt}\right) \tag{181}$$

$$d^i_{T_{PCR_{pos}},T_{PCR_{neg}}} \sim \text{Binom}\left(T^i_{PCR_{pos}}(t),\ 1 - e^{-\gamma_{PCR_{pos}} dt}\right) \tag{182}$$

Model compartments were then updated as follows (Note that $d^{i,j_1^{reinf},k}_{S,E} = d^{i,j_2^{reinf},k}_{S,E} = 0$):

$$\begin{aligned}S^{i,k}(t+dt) :=\ & S^{i,k}(t) - d^{i,j_1,k}_{S,E} - d^{i,j_2,k}_{S,E} + d^{i,k-1}_{S,v} - d^{i,k}_{S,v} + \mathbb{1}_{\{5\}}(k) d^{i,3}_{S,v,2} - \mathbb{1}_{\{3\}} d^{i,3}_{S,v,2} - d^{i,j,k}_{seed} - d^{i,j_2,k}_{seed} \\ & + \sum_j d^{i,j,k}_{R,S} + d^{i,j_H,k}_{R,S}\end{aligned} \tag{183}$$

$$\begin{aligned}E^{i,j,k,1}(t+dt) :=\ & E^{i,j,k,1}(t) + d^{i,j,k}_{S,E} + \mathbb{1}_{\left\{j_1^{reinf}\right\}}(j) d^{i,j_H,k,j_1}_{R,E} + \mathbb{1}_{\left\{j_2^{reinf}\right\}}(j)\left(d^{i,j_H,k,j_1}_{R,E} + d^{i,j_1,k,j_1}_{R,E} + d^{i,j_1^{reinf},k,j_1}_{R,E}\right) \\ & - d^{i,j,k}_{E,E} + d^{i,j,k-1,1}_{E,v} - d^{i,j,k,1}_{E,v} + \mathbb{1}_{\{5\}}(k) d^{i,j,3,1}_{E,v,2} - \mathbb{1}_{\{3\}} d^{i,j,3,1}_{E,v,2} + d^{i,j,k}_{seed}\end{aligned} \tag{184}$$

$$E^{i,j,k,2}(t+dt) := E^{i,j,k,2}(t) + d^{i,j,k}_{E,E} - d^{i,j,k}_{E,I_A} - d^{i,j,k}_{E,I_P} + d^{i,j,k-1,2}_{E,v} - d^{i,j,k,2}_{E,v} + \mathbb{1}_{\{5\}}(k) d^{i,j,3,2}_{E,v,2} - \mathbb{1}_{\{3\}} d^{i,j,3,2}_{E,v,2} \tag{185}$$

$$I^{i,j,k}_A(t+dt) := I^{i,j,k}_A(t) + d^{i,j,k}_{E,I_A} - d^{i,j,k}_{I_A,R} + d^{i,j,k-1}_{I_A,v} - d^{i,j,k}_{I_A,v} + \mathbb{1}_{\{5\}}(k) d^{i,j,3}_{I_A,v,2} - \mathbb{1}_{\{3\}} d^{i,j,3}_{I_A,v,2} \tag{186}$$

$$I^{i,j,k}_P(t+dt) := I^{i,j,k}_P(t) + d^{i,j,k}_{E,I_P} - d^{i,j,k}_{I_P,I_{C_1}} + d^{i,j,k-1}_{I_P,v} - d^{i,j,k}_{I_P,v} + \mathbb{1}_{\{5\}}(k) d^{i,j,3}_{I_P,v,2} - \mathbb{1}_{\{3\}} d^{i,j,3}_{I_P,v,2} \tag{187}$$

$$I^{i,j,k}_{C_1}(t+dt) := I^{i,j,k}_{C_1}(t) + d^{i,j,k}_{I_P,I_{C_1}} - d^{i,j,k}_{I_{C_1},I_{C_2}} \tag{188}$$

$$\begin{aligned}I^{i,j,k}_{C_2}(t+dt) :=\ & I^{i,j,k}_{C_2}(t) + d^{i,j,k}_{I_{C_1},I_{C_2}} - d^{i,j,k}_{I_{C_2},G_D} - d^{i,j,k}_{I_{C_2},R} - d^{i,j,k}_{I_{C_2},ICU_{pre}} \\ & - d^{i,j,k}_{I_{C_2},ICU_{pre*}} - d^{i,j,k}_{I_{C_2},H_R} - d^{i,j,k}_{I_{C_2},H_{R*}} - d^{i,j,k}_{I_{C_2},H_D} - d^{i,j,k}_{I_{C_2},H_{D*}}\end{aligned} \tag{189}$$

$$G^{i,j,k,1}_D(t+dt) := G^{i,j,k,1}_D(t) + d^{i,j,k}_{I_{C_2},G_D} - d^{i,j,k}_{G_D,G_D} \tag{190}$$

$$G^{i,j,k,2}_D(t+dt) := G^{i,j,k,2}_D(t) + d^{i,j,k}_{G_D,G_D} - d^{i,j,k}_{G_D,D} \tag{191}$$

$$\begin{aligned}ICU^{i,j,k}_{pre}(t+dt) :=\ & ICU^{i,j,k}_{pre}(t) + d^{i,j,k}_{I_{C_2},ICU_{pre}} - d^{i,j,k}_{ICU_{pre},ICU_{W_R}} - d^{i,j,k}_{ICU_{pre},ICU_{W_D}} \\ & - d^{i,j,k}_{ICU_{pre},ICU_D} - d^{i,j,k}_{ICU_{pre},ICU_{pre*}} - d^{i,j,k}_{ICU_{pre},ICU_{W_{R*}}} - d^{i,j,k}_{ICU_{pre},ICU_{W_{D*}}} \\ & - d^{i,j,k}_{ICU_{pre},ICU_{D*}}\end{aligned} \tag{192}$$

$$\begin{aligned}ICU^{i,j,k}_{pre*}(t+dt) :=\ & ICU^{i,j,k}_{pre*}(t) + d^{i,j,k}_{I_{C_2},ICU_{pre*}} - d^{i,j,k}_{ICU_{pre},ICU_{W_{D*}}} - d^{i,j,k}_{ICU_{pre*},ICU_{W_{R*}}} \\ & - d^{i,j,k}_{ICU_{pre*},ICU_{D*}}\end{aligned} \tag{193}$$

$$\begin{aligned} ICU^{i,j,k}_{W_R}(t+dt) &:= ICU^{i,j,k}_{W_R}(t) + d^{i,j,k}_{ICU_{pre},ICU_{W_R}} - d^{i,j,k}_{ICU_{W_R},W_R} - d^{i,j,k}_{ICU_{W_R},ICU_{W_{R^*}}} \\ &\quad - d^{i,j,k}_{ICU_{W_R},W_{R^*}} \end{aligned} \tag{194}$$

$$\begin{aligned} ICU^{i,j,k}_{W_{R^*}}(t+dt) &:= ICU^{i,j,k}_{W_{R^*}}(t) + d^{i,j,k}_{ICU_{pre^*},ICU_{W_{R^*}}} + d^{i,j,k}_{ICU_{W_R},ICU_{W_{R^*}}} + d^{i,j,k}_{ICU_{pre},ICU_{W_{R^*}}} \\ &\quad - d^{i,j,k}_{ICU_{W_{R^*}},W_{R^*}} \end{aligned} \tag{195}$$

$$\begin{aligned} ICU^{i,j,k}_{W_D}(t+dt) &:= ICU^{i,j,k}_{W_D}(t) + d^{i,j,k}_{ICU_{pre},ICU_{W_D}} - d^{i,j,k}_{ICU_{W_D},W_D} - d^{i,j,k}_{ICU_{W_D},ICU_{W_{D^*}}} \\ &\quad - d^{i,j,k}_{ICU_{W_D},W_{D^*}} \end{aligned} \tag{196}$$

$$\begin{aligned} ICU^{i,j,k}_{W_{D^*}}(t+dt) &:= ICU^{i,j,k}_{W_{D^*}}(t) + d^{i,j,k}_{ICU_{pre^*},ICU_{W_{D^*}}} + d^{i,j,k}_{ICU_{W_D},ICU_{W_{D^*}}} + d^{i,j,k}_{ICU_{pre},ICU_{W_{D^*}}} \\ &\quad - d^{i,j,k}_{ICU_{W_{D^*}},W_{D^*}} \end{aligned} \tag{197}$$

$$\begin{aligned} ICU^{i,j,k,1}_{D}(t+dt) &:= ICU^{i,j,k,1}_{D}(t) + d^{i,j,k}_{ICU_{pre},ICU_D} - d^{i,j,k}_{ICU_D,ICU_D} - d^{i,j,k,1,1}_{ICU_D,ICU_{D^*}} \\ &\quad - d^{i,j,k,1,2}_{ICU_D,ICU_{D^*}} \end{aligned} \tag{198}$$

$$ICU^{i,j,k,2}_{D}(t+dt) := ICU^{i,j,k,2}_{D}(t) + d^{i,j,k}_{ICU_D,ICU_D} - d^{i,j,k}_{ICU_D,D} - d^{i,j,k,2,2}_{ICU_D,ICU_{D^*}} \tag{199}$$

$$\begin{aligned} ICU^{i,j,k,1}_{D^*}(t+dt) &:= ICU^{i,j,k,1}_{D^*}(t) + d^{i,j,k}_{ICU_{pre^*},ICU_{D^*}} + d^{i,j,k,1,1}_{ICU_D,ICU_{D^*}} + d^{i,j,k}_{ICU_{pre},ICU_{D^*}} \\ &\quad - d^{i,j,k}_{ICU_{D^*},ICU_{D^*}} \end{aligned} \tag{200}$$

$$\begin{aligned} ICU^{i,j,k,2}_{D^*}(t+dt) &:= ICU^{i,j,k,2}_{D^*}(t) + d^{i,j,k}_{ICU_{D^*},ICU_{D^*}} + d^{i,j,k,1,2}_{ICU_D,ICU_{D^*}} + d^{i,j,k,2,2}_{ICU_D,ICU_{D^*}} \\ &\quad - d^{i,j,k}_{ICU_{D^*},D} \end{aligned} \tag{201}$$

$$W^{i,j,k,1}_R(t+dt) := W^{i,j,k,1}_R(t) + d^{i,j,k}_{ICU_{W_R},W_R} - d^{i,j,k}_{W_R,W_R} - d^{i,j,k,1,1}_{W_R,W_{R^*}} - d^{i,j,k,1,2}_{W_R,W_{R^*}} \tag{202}$$

$$W^{i,j,k,2}_R(t+dt) := W^{i,j,k,2}_R(t) + d^{i,j,k}_{W_R,W_R} - d^{i,j,k}_{W_R,R} - d^{i,j,k,2,2}_{W_R,W_{R^*}} \tag{203}$$

$$W^{i,j,k,1}_{R^*}(t+dt) := W^{i,j,k,1}_{R^*}(t) + d^{i,j,k}_{ICU_{W_{R^*}},W_{R^*}} + d^{i,j,k,1,1}_{W_R,W_{R^*}} + d^{i,j,k}_{ICU_{W_R},W_{R^*}} - d^{i,j,k}_{W_{R^*},W_{R^*}} \tag{204}$$

$$W^{i,j,k,2}_{R^*}(t+dt) := W^{i,j,k,2}_{R^*}(t) + d^{i,j,k}_{W_{R^*},W_{R^*}} + d^{i,j,k,2,2}_{W_R,W_{R^*}} + d^{i,j,k,1,2}_{W_R,W_{R^*}} - d^{i,j,k}_{W_{R^*},R} \tag{205}$$

$$W^{i,j,k}_D(t+dt) := W^{i,j,k}_D(t) + d^{i,j,k}_{ICU_{W_D},W_D} - d^{i,j,k}_{W_D,D} - d^{i,j,k}_{W_D,W_{D^*}} \tag{206}$$

$$W^{i,j,k}_{D^*}(t+dt) := W^{i,j,k}_{D^*}(t) + d^{i,j,k}_{ICU_{W_{D^*}},W_{D^*}} + d^{i,j,k}_{W_D,W_{D^*}} + d^{i,j,k}_{ICU_{W_D},W_{D^*}} - d^{i,j,k}_{W_{D^*},D} \tag{207}$$

$$H^{i,j,k,1}_D(t+dt) := H^{i,j,k,1}_D(t) + d^{i,j,k}_{I_{C_2},H_D} - d^{i,j,k}_{H_D,H_D} - d^{i,j,k,1,1}_{H_D,H_{D^*}} - d^{i,j,k,1,2}_{H_D,H_{D^*}} \tag{208}$$

$$H^{i,j,k,2}_D(t+dt) := H^{i,j,k,2}_D(t) + d^{i,j,k}_{H_D,H_D} - d^{i,j,k}_{H_D,D} - d^{i,j,k,2,2}_{H_D,H_{D^*}} \tag{209}$$

$$H^{i,j,k,1}_{D^*}(t+dt) := H^{i,j,k,1}_{D^*}(t) + d^{i,j,k}_{I_{C_2},H_{D^*}} + d^{i,j,k,1,1}_{H_D,H_{D^*}} - d^{i,j,k}_{H_{D^*},H_{D^*}} \tag{210}$$

$$H^{i,j,k,2}_{D^*}(t+dt) := H^{i,j,k,2}_{D^*}(t) + d^{i,j,k}_{H_{D^*},H_{D^*}} + d^{i,j,k,2,2}_{H_D,H_{D^*}} + d^{i,j,k,1,2}_{H_D,H_{D^*}} - d^{i,j,k}_{H_{D^*},D} \tag{211}$$

$$H^{i,j,k}_R(t+dt) := H^{i,j,k}_R(t) + d^{i,j,k}_{I_{C_2},H_R} - d^{i,j,k}_{H_R,R} - d^{i,j,k}_{H_R,H_{R^*}} \tag{212}$$

$$H^{i}_{R^*}(t+dt) := H^{i,j,k}_{R^*}(t) + d^{i,j,k}_{I_{C_2},H_{R^*}} + d^{i,j,k}_{H_R,H_{R^*}} - d^{i,j,k}_{H_{R^*},R} \tag{213}$$

$$\begin{aligned} R^{i,j,k}(t+dt) &:= R^{i,j,k}(t) + \mathbb{1}_{\left\{j_1,j_2,j_1^{reinf},j_2^{reinf}\right\}}(j)\left(d^{i,j,k}_{I_A,R} + d^{i,j,k}_{I_{C_2},R} + d^{i,j,k}_{H_R,R} + d^{i,j,k}_{H_{R^*},R} + d^{i,j,k}_{W_R,R} + d^{i,j,k}_{W_{R^*},R}\right) \\ &\quad - d^{i,j,k}_{R,S} - \mathbb{1}_{\{j_H\}} d^{i,j,k,j_1}_{R,E} + \mathbb{1}_{\left\{j_H,j_1,j_1^{reinf}\right\}} d^{i,j,k,j_2}_{R,E} + d^{i,j,k-1}_{R,v} - d^{i,j,k}_{R,v} + \mathbb{1}_{\{5\}}(k) d^{i,j,3}_{R,v,2} - \mathbb{1}_{\{3\}} d^{i,j,3}_{R,v,2} \end{aligned} \tag{214}$$

$$T^i_{sero^1_{pre}}(t+dt) := T^i_{sero^1_{pre}}(t) - d^i_{T_{sero^1_{pre}},T_{sero^1_{pos}}} - d^i_{T_{sero^1_{pre}},T_{sero^1_{neg}}} + \sum_j \sum_k \left(d^{i,j,k}_{E,I_A} + d^{i,j,k}_{E,I_P}\right) \tag{215}$$

$$T^i_{sero^1_{pos}}(t+dt) := T^i_{sero^1_{pos}}(t) + d^i_{T_{sero^1_{pre}},T_{sero^1_{pos}}} - d^i_{T_{sero^1_{pos}},T_{sero^1_{neg}}} \tag{216}$$

$$T^i_{sero^1_{neg}}(t+dt) := T^i_{sero^1_{neg}}(t) + d^i_{T_{sero^1_{pre}},T_{sero^1_{neg}}} + d^i_{T_{sero^1_{pos}},T_{sero^1_{neg}}} \tag{217}$$

$$T^i_{sero^2_{pre}}(t+dt) := T^i_{sero^2_{pre}}(t) - d^i_{T_{sero^2_{pre}},T_{sero^2_{pos}}} - d^i_{T_{sero^2_{pre}},T_{sero^2_{neg}}} + \sum_j \sum_k \left(d^{i,j,k}_{E,I_A} + d^{i,j,k}_{E,I_P}\right) \tag{218}$$

$$T^i_{sero^2_{pos}}(t+dt) := T^i_{sero^2_{pos}}(t) + d^i_{T_{sero^2_{pre}},T_{sero^2_{pos}}} - d^i_{T_{sero^2_{pos}},T_{sero^2_{neg}}} \tag{219}$$

$$T^i_{sero^2_{neg}}(t+dt) := T^i_{sero^2_{neg}}(t) + d^i_{T_{sero^2_{pre}}, T_{sero^2_{neg}}} + d^i_{T_{sero^2_{pos}}, T_{sero^2_{neg}}} \tag{220}$$

$$T^i_{PCR_{pre}}(t+dt) := T^i_{PCR_{pre}}(t) - d^i_{T_{PCR_{pre}}, T_{PCR_{pos}}} + \sum_j \sum_k \left( d^{i,j,k}_{S,E} + d^{i,j,k,j_1}_{R,E} + d^{i,j,k,j_2}_{R,E} \right) + \sum_k \left( d^{i,j_H,k,j_1}_{R,E} + d^{i,j_H,k,j_2}_{R,E} \right) \tag{221}$$

$$T^i_{PCR_{pos}}(t+dt) := T^i_{PCR_{pos}}(t) + d^i_{T_{PCR_{pre}}, T_{PCR_{pos}}} - d^i_{T_{PCR_{pos}}, T_{PCR_{neg}}} \tag{222}$$

$$T^i_{PCR_{neg}}(t+dt) := T^i_{PCR_{neg}}(t) + d^i_{T_{PCR_{pos}}, T_{PCR_{neg}}}. \tag{223}$$

$$\tag{224}$$

Note that the fitted seeding dates of the epidemic ($t_{Wildtype}$), Alpha variant ($t_{Alpha}$), Delta variant ($t_{Delta}$) and Omicron BA.1 variant ($t_{Omicron}$) have continuous support. The seeding process (see Section 2.1) is handled within the discretisation to four update steps per day such that:

$$\hat{\delta}^{i,j,k}(t) = \begin{cases} \phi_j f_j(t) & \text{if } i = [15, 20),\ j \in \{Wildtype,\ Alpha,\ Delta,\ Omicron\},\ k = 0 \\ 0 & \text{otherwise,} \end{cases} \tag{225}$$

where

$$f_j(t) = \begin{cases} \left( \left\lceil \frac{t_j}{dt} \right\rceil - \frac{t_j}{dt} \right) & \text{if } t = dt \left\lfloor \frac{t_j}{dt} \right\rfloor \\ 1 & \text{if } dt \left\lfloor \frac{t_j}{dt} \right\rfloor < t < dt \left\lfloor \frac{t_j}{dt} \right\rfloor + \nu_j \\ \left( \frac{t_j}{dt} - \left\lfloor \frac{t_j}{dt} \right\rfloor \right) & \text{if } t = dt \left\lfloor \frac{t_j}{dt} \right\rfloor + \nu_j \\ 0 & \text{otherwise.} \end{cases} \tag{226}$$

where $\lfloor . \rfloor$ and $\lceil . \rceil$ denote the floor and ceiling functions respectively.

#### 4.5 Observation process

To describe the epidemic in each NHS region, we fitted our model to time series data on hospital admissions, hospital ward occupancy (both in general beds and in ICU beds), deaths in hospitals, deaths in the community, population serological surveys, PCR testing data and Variant and Mutation (VAM) data (see Table S1).

##### 4.5.1 Notation for distributions used in this section

If $X \sim \text{Binom}(n,p)$, then $X$ follows a binomial distribution with mean $np$ and variance $np(1-p)$, such that

$$P(X = x) = P_{\text{Binom}}(x|n,\ p) = \binom{n}{x} p^x (1-p)^{(n-x)}. \tag{227}$$

If $Y \sim \text{NegBinom}(m,\kappa)$, then $Y$ follows a negative binomial distribution with mean $m$ and shape $\kappa$, such that

$$P(Y = y) = P_{\text{NegBinom}}(y|m,\kappa) = \frac{\Gamma(\kappa+y)}{y!\Gamma(\kappa)} \left( \frac{\kappa}{\kappa+m} \right)^{\kappa} \left( \frac{m}{\kappa+m} \right)^{y}, \tag{228}$$

where $\Gamma(x)$ is the gamma function. The variance of $Y$ is $m + m^2/\kappa$.

If $Z \sim \text{BetaBinom}(n,\omega,\rho)$, then $Z$ follows a beta-binomial distribution with size $n$, mean probability $\omega$ and overdispersion parameter $\rho$, such that

$$P(Z = z) = P_{\text{BetaBinom}}(z|n,\omega,\rho) = \binom{n}{z} \frac{B(z+a, n-z+b)}{B(a,b)}, \tag{229}$$

where $a = \omega \left( \frac{1-\rho}{\rho} \right)$, $b = (1-\omega) \left( \frac{1-\rho}{\rho} \right)$ and $B(a,b)$ is the beta function. The mean of Z is $n\omega$ and the variance is $n\omega(1-\omega)[1+(n-1)\rho]$.

In the following, we use $t$ to represent a day with observations. Note that different data streams had different sets of days with observations.

#### 4.5.2 Hospital admissions and new diagnoses in hospital

We represented $Y^z_{adm}(t)$, the daily number of confirmed COVID-19 hospital admissions and new diagnoses for existing hospitalised cases in age band $z \in Z_{adm}$ ($Z_{adm} = \{[0,10),[10,20),\ldots,[70,80),[80+)\}$), as the observed realisations of an underlying hidden Markov process, $X^z_{adm}(t)$, defined as:

$$\begin{aligned} X^z_{adm}(t) := \sum_{i \in z}\sum_j\sum_k \Big( & d^{i,j,k}_{I_C,H_{R^*}} + d^{i,j,k}_{I_C,H_{D^*}} + d^{i,j,k}_{I_C,ICU_{pre^*}} + d^{i,j,k}_{H_R,H_{R^*}} + d^{i,j,k}_{ICU_{pre},ICU_{pre^*}} + d^{i,j,k}_{ICU_{W_R},ICU_{W_{R^*}}} \\ & + d^{i,j,k}_{ICU_{W_D},ICU_{W^*_D}} + d^{i,j,k}_{W_D,W_{D^*}} + d^{i,j,k,1,1}_{H_D H_{D^*}} + d^{i,j,k,1,2}_{H_D,H_{D^*}} + d^{i,j,k,2,2}_{H_D,H_{D^*}} + d^{i,j,k,1,1}_{ICU_D,ICU_{D^*}} \\ & + d^{i,j,k,1,2}_{ICU_D,ICU_{D^*}} + d^{i,j,k,2,2}_{ICU_D,ICU_{D^*}} + d^{i,j,k,1,1}_{W_R,W_{R^*}} + d^{i,j,k,1,2}_{W_R,W_{R^*}} + d^{i,j,k,2,2}_{W_R,W_{R^*}} + d^{i,j,k}_{ICU_{pre},ICU_{W_{R^*}}} \\ & + d^{i,j,k}_{ICU_{pre},ICU_{W_{D^*}}} + d^{i,j,k}_{ICU_{pre},ICU_{D^*}} + d^{i,j,k}_{ICU_{W_{D^*}},W_{D^*}} + d^{i,j,k}_{ICU_{W_{R^*}},W_{R^*}} \Big) \end{aligned} \tag{230}$$

which was related to the data via a reporting distribution:

$$Y^z_{adm}(t) \sim \text{NegBinom}\left(X^z_{adm}(t), \kappa_A\right). \tag{231}$$

We allow for overdispersion in the observation process to account for noise in the underlying data streams, for example due to day-of-week effects on data collection. We fit the overdispersion parameter $\alpha_A = \frac{1}{\kappa_A}$.

The contribution to the likelihood of the data on hospital admissions and new diagnoses in hospital in age band $z$ was therefore:

$$\mathscr{L}_{adm} = \prod_t \prod_{z \in Z_{adm}} P_{\text{NegBinom}}\left(Y^z_{adm}(t) \,\middle|\, X^z_{adm}(t),\ \kappa_A\right) \tag{232}$$

#### 4.5.3 Hospital bed occupancy by confirmed COVID-19 cases

The model predicted general hospital bed occupancy by confirmed COVID-19 cases, $X_{hosp}(t)$ as:

$$X_{hosp}(t) := \sum_i\sum_j\sum_k \left( H^{i,j,k}_{R^*}(t) + H^{i,j,k,1}_{D^*}(t) + H^{i,j,k,2}_{D^*}(t) + ICU^{i,j,k}_{pre^*}(t) + W^{i,j,k}_{D^*}(t) + W^{i,j,k,1}_{R^*}(t) + W^{i,j,k,2}_{R^*}(t) \right), \tag{233}$$

which was related to the observed daily general bed-occupancy via a reporting distribution:

$$Y_{hosp}(t) \sim \text{NegBinom}\left(X_{hosp}(t), \kappa_H\right). \tag{234}$$

Similarly, the model predicted ICU bed occupancy by confirmed COVID-19 cases, $X_{ICU}(t)$ as:

$$X_{ICU}(t) := \sum_i\sum_j\sum_k \left( ICU^{i,j,k}_{W_{R^*}}(t) + ICU^{i,j,k}_{W_{D^*}}(t) + ICU^{i,j,k,1}_{D^*}(t) + ICU^{i,j,k,2}_{D^*}(t) \right), \tag{235}$$

which was related to the observed daily ICU bed-occupancy via a reporting distribution:

$$Y_{ICU}(t) \sim \text{NegBinom}\left(X_{ICU}(t), \kappa_H\right). \tag{236}$$

We fit the overdispersion parameter $\alpha_H = \frac{1}{\kappa_H}$, which we use for both general hospital bed and ICU bed occupancy.

The overall contribution to the likelihood of the data on general bed and ICU bed occupancy was:

$$\begin{aligned} \mathscr{L}_{beds} = & \prod_t P_{\text{NegBinom}}\left(Y_{hosp}(t) \,\middle|\, X_{hosp}(t), \kappa_H\right) \\ & \times \prod_t P_{\text{NegBinom}}\left(Y_{ICU}(t) \,\middle|\, X_{ICU}(t), \kappa_H\right). \end{aligned} \tag{237}$$

#### 4.5.4 Hospital and community COVID-19 deaths

We considered $Y^z_{D_{hosp}}(t)$, the reported number of daily COVID-19 deaths in hospitals in age band $z \in Z_D$ ($Z_D = \{[0,50), [50,55), [55,60) \ldots, [75,80), [80+)\}$), as the observed realisation of an underlying hidden Markov process, $X^z_{D_{hosp}}(t)$, defined as:

$$X^z_{D_{hosp}}(t) := \sum_{i \in z} \sum_j \sum_k \left( d^{i,j,k}_{H_D,D} + d^{i,j,k}_{H_{D^*},D} + d^{i,j,k}_{ICU_D,D} + d^{i,j,k}_{ICU_{D^*},D} + d^{i,j,k}_{W_D,D} + d^{i,j,k}_{W_{D^*},D} \right), \tag{238}$$

which was related to the data via a reporting distribution:

$$Y^z_{D_{hosp}}(t) \sim \text{NegBinom}\left( X^z_{D_{hosp}}(t), \kappa_{D_{hosp}} \right). \tag{239}$$

Similarly, we represented the reported number of daily COVID-19 deaths in the community in age band $z \in Z_D$, $Y^z_{D_{comm}}(t)$, as the observed realisations of an underlying hidden Markov process, $X^z_{D_{comm}}(t)$, defined as:

$$X^z_{D_{comm}}(t) := \sum_{i \in z} \sum_j \sum_k d^{i,j,k}_{G_D,D}, \tag{240}$$

which was related to the data via a reporting distribution:

$$Y^z_{D_{comm}}(t) \sim \text{NegBinom}\left( X^z_{D_{comm}}(t), \kappa_{D_{comm}} \right). \tag{241}$$

We fit the overdispersion parameters $\alpha_{D_{hosp}} = \frac{1}{\kappa_{D_{hosp}}}$ for hospital deaths and $\alpha_{D_{comm}} = \frac{1}{\kappa_{D_{comm}}}$ for community deaths.

The overall contribution to the likelihood of the data on COVID-19 deaths in hospitals and the community was:

$$\begin{aligned} \mathscr{L}_{deaths} = & \prod_t \prod_{z \in Z_D} P_{\text{NegBinom}}\left( Y_{D_{hosp}}(t) \middle| X_{D_{hosp}}(t), \kappa_{D_{hosp}} \right) \\ & \times \prod_t \prod_{z \in Z_D} P_{\text{NegBinom}}\left( Y_{D_{comm}}(t) \middle| X_{D_{comm}}(t), \kappa_{D_{comm}} \right) \end{aligned} \tag{242}$$

#### 4.5.5 Serosurveys

We model serological testing of all individuals aged 15-64 inclusive, and define the resulting number of seropositive and seronegative individuals (were all individuals aged 15-65 to be tested) from serology flow $j$ (where $j = 1$ corresponds to EuroImmun and $j = 2$ to Roche N), as:

$$X_{sero^j_{pos}}(t) := \sum_{i=[15,20)}^{[60,65)} T^i_{sero^j_{pos}}(t) \tag{243}$$

$$X_{sero^j_{neg}}(t) := \left( \sum_{i=[15,20)}^{[60,65)} N^i \right) - X_{sero^j_{pos}}(t). \tag{244}$$

We compared the observed number of seropositive results, $Y_{sero^j_{pos}}(t)$, with that predicted by our model, allowing for i) the sample size of each serological survey, $Y_{sero^j_{test}}(t)$ and ii) imperfect sensitivity ($p_{sero_{sens}}$) and specificity ($p_{sero_{spec}}$) of the serological assay:

$$Y_{sero^j_{pos}}(t) \sim \text{Binom}\left( Y_{sero^j_{test}}(t), \omega_{sero_{pos}}(t) \right) \tag{245}$$

where:

$$\omega_{sero^j_{pos}}(t) := \frac{p_{sero_{sens}} X_{sero^j_{pos}}(t) + \left(1 - p_{sero_{spec}}\right) X_{sero^j_{neg}}(t)}{X_{sero^j_{pos}}(t) + X_{sero^j_{neg}}(t)}. \tag{246}$$

The contribution to the likelihood of the serosurvey data was:

$$\mathscr{L}_{sero} = \prod_t \prod_{j=1,2} P_{\text{Binom}}\left( Y_{sero^j_{pos}}(t) \middle| X_{sero^j_{test}}(t), \omega_{sero^j_{pos}}(t) \right) \tag{247}$$

##### 4.5.6 PCR testing

As described in the data section (section 4), we fitted the model to PCR testing data from three separate sources:

- Pillar 2 testing by age: government community testing programme, which recommended that symptomatic individuals in the community with COVID-19 symptoms were tested. [13].
- REACT-1 by age: study which aimed to quantify the prevalence of SARS-CoV-2 in a random sample of the England population between 07-05-2020 and 31-03-2022 (note data was capped for analysis on 24-02-20222). [14].
- ONS survey: government weekly infection survey which aims to quantify the prevalence of SARS-CoV-2 in a random sample of the UK population (completely independent from REACT-1) on an ongoing basis. [15]

We fit to Pillar 2 PCR test results for each age band $z \in Z_{P2}$ ($Z_{P2} = \{[15,25),[25,50),[50,65),[65,80),[80+)\}$) and assume that individuals tested through this government programme were either newly symptomatic SARS-CoV-2 cases (who will test positive):

$$X^z_{P2_{pos}}(t) := \sum_{i\in z}\sum_j\sum_k d^{i,j,k}_{I_P,I_{C_1}} \tag{248}$$

or non-SARS-CoV-2 cases who have symptoms consistent with COVID-19 (who will test negative):

$$X^z_{P2_{neg}}(t) := g^z(t)\left(\left(\sum_{i\in z} N^i\right) - X^z_{P2_{pos}}(t)\right), \tag{249}$$

where

$$g^z(t) = \begin{cases} p^z_{NC} & \text{if } t \text{ is a weekday} \\ p^z_{NCW} & \text{if } t \text{ is a weekend day} \end{cases} \tag{250}$$

is the probability of non SARS-CoV-2 cases in age band $z$ having symptoms consistent with COVID-19 that might lead them to get a PCR test.

We compared the observed number of positive PCR tests, $Y^z_{P2_{pos}}(t)$ with that predicted by our model, accounting for the number of PCR tests conducted each day under pillar 2, $Y^z_{P2_{test}}(t)$, by calculating the probability of a positive PCR result (assuming perfect sensitivity and specificity of the PCR test):

$$\omega^z_{P2_{pos}}(t) := \frac{X^z_{P2_{pos}}(t)}{X^z_{P2_{pos}}(t) + X^z_{P2_{neg}}(t)} \tag{251}$$

People may seek PCR tests for many reasons and thus the pillar 2 data are subject to competing biases. We therefore allowed for an over-dispersion parameter $\rho_{P2_{test}}$, which we fitted separately for each region in the modelling framework:

$$Y^z_{P2_{pos}}(t) \sim \text{BetaBinom}\left(Y^z_{P2_{test}}(t), \omega^z_{P2_{pos}}(t), \rho_{P2_{test}}\right). \tag{252}$$

We incorporated the REACT-1 PCR testing data for each age band $z \in Z_{R1}$ ($Z_{R1} = \{[5,24),[25,34),[35,44),[45,54),[55,64),[65+)\}$) into the likelihood analogously to the serology data, by considering the model-predicted number of PCR-positives, $X^z_{R1_{pos}}(t)$, and PCR-negatives, $X^z_{R1_{neg}}(t)$:

$$X^z_{R1_{pos}}(t) := \sum_{i\in z} T^i_{PCR_{pos}}(t), \tag{253}$$

$$X^z_{R1_{neg}}(t) := \left(\sum_{i\in z} N^i\right) - X^z_{R1_{pos}}(t). \tag{254}$$

We compared the daily number of positive results observed in REACT-1, $Y^z_{R1_{pos}}(t)$, given the number of people tested on that day, $Y^z_{R1_{test}}(t)$, to our model predictions, by calculating the probability of a positive result, assuming perfect sensitivity and specificity of the REACT-1 assay:

$$\omega^z_{R1_{pos}}(t) := \frac{X^z_{R1_{pos}}(t)}{X^z_{R1_{pos}}(t) + X^z_{R1_{neg}}(t)} \tag{255}$$

so

$$Y^z_{R1_{pos}}(t) \sim \text{Binom}\left(Y^z_{R1_{test}}(t), \omega^z_{R1_{pos}}(t)\right). \tag{256}$$

Finally, we fit to ONS PCR testing data, for which the pool of testing is individuals aged 2 years and over. We consider the model-predicted number of PCR-positives, $X_{ONS_{pos}}(t)$, and PCR-negatives, $X_{ONS_{neg}}(t)$, in the population eligible for ONS PCR testing:

$$X_{ONS_{pos}}(t) := \frac{3}{5}T^{[0,5)}_{PCR_{pos}}(t) + \sum_{i=[5,10)}^{[80+)} T^{i}_{PCR_{pos}}(t), \tag{257}$$

$$X_{ONS_{neg}}(t) := \frac{3}{5}N^{[0,5)} + \left(\sum_{i=[5,10)}^{[80+)} N^{i}\right) - X_{ONS_{pos}}(t), \tag{258}$$

and we compared the daily number of positive results observed in the ONS infection survey, $Y_{ONS_{pos}}(t)$, given the number of people tested on that day, $Y_{ONS_{test}}(t)$, to our model predictions, by calculating the probability of a positive result, assuming perfect sensitivity and specificity of the assay:

$$\omega_{ONS_{pos}}(t) := \frac{X_{ONS_{pos}}(t)}{X_{ONS_{pos}}(t) + X_{ONS_{neg}}(t)} \tag{259}$$

so

$$Y_{ONS_{pos}}(t) \sim \text{Binom}\left(Y_{ONS_{test}}(t), \omega_{ONS_{pos}}(t)\right). \tag{260}$$

The contribution to the likelihood of the PCR testing data was:

$$\begin{aligned}
\mathscr{L}_{PCR} = &\prod_{t}\prod_{z \in Z_{P2}} P_{\text{BetaBinom}}\left(Y\text{-}P2_{pos}(t) \,\middle|\, Y^{z}_{P2_{test}}(t), \omega^{z}_{P2_{pos}}(t), \rho_{P2_{test}}\right) \\
&\times \prod_{t}\prod_{z \in Z_{R1}} P_{\text{Binom}}\left(Y^{z}_{R1_{pos}}(t) \,\middle|\, Y^{z}_{R1_{test}}(t), \omega^{z}_{R1_{pos}}(t)\right) \\
&\times \prod_{t} P_{\text{Binom}}\left(Y_{ONS_{pos}}(t) \,\middle|\, Y_{ONS_{test}}(t), \omega_{ONS_{pos}}(t)\right)
\end{aligned} \tag{261}$$

#### 4.5.7 Variant and Mutation data

To inform the replacement of the Wildtype variant by the Alpha variant, the Alpha variant by the Delta variant and the Delta variant by the Omicron variant, we fitted to Variant and Mutation (VAM) data. During each stage we have a variant pair $(j_1, j_2)$, where $j_2$ is the emerging variant. We assume that samples tested for VAM are newly symptomatic cases (across all age groups), with the number for variants $j_1$ and $j_2$ given by

$$X_{VAM_1}(t) := \sum_{i}\sum_{k} d^{i,j_1,k}_{I_P,I_{C_1}} + d^{i,j_1^{reinf},k}_{I_P,I_{C_1}} \tag{262}$$

$$X_{VAM_2}(t) := \sum_{i}\sum_{k} d^{i,j_2,k}_{I_P,I_{C_1}} + d^{i,j_2^{reinf},k}_{I_P,I_{C_1}} \tag{263}$$

We compared the observed number of variant $j_2$ VAM test results, $Y_{VAM_2}(t)$ with that predicted by our model, accounting for the combined number of variant $j_1$ and $j_2$ VAM test results each day, $Y_{VAM_{test}}(t)$, by calculating the probability of a variant $j_2$ VAM test result:

$$\omega_{VAM_2}(t) := \frac{X_{VAM_2}(t)}{X_{VAM_1}(t) + X_{VAM_2}(t)} \tag{264}$$

so

$$Y_{VAM_2}(t) \sim \text{Binom}\left(Y_{VAM_{test}}(t), \omega_{VAM_2}(t)\right). \tag{265}$$

The contribution to the likelihood of the VAM data was:

$$\mathscr{L}_{VAM} = \prod_{t} P_{\text{Binom}}\left(Y_{VAM_2}(t) \,\middle|\, Y_{VAM_{test}}(t), \omega_{VAM_2}(t)\right) \tag{266}$$

#### 4.5.8 Full likelihood

The overall likelihood was then calculated as the product of the likelihoods of the individual observations, i.e.:

$$\mathscr{L} = \mathscr{L}_{adm} \times \mathscr{L}_{beds} \times \mathscr{L}_{deaths} \times \mathscr{L}_{sero} \times \mathscr{L}_{PCR} \times \mathscr{L}_{VAM}. \tag{267}$$

#### 4.6 Reproduction number

Both $R_t^j$ and $R_t^{j,eff}$ are calculated using next generation matrix (NGM) methods [60]. Note that in this calculation only, we make a simplifying assumption that individuals cannot change vaccine strata between initial infection and the end of their infectious period (or death).

To compute the next generation matrix, we calculated the mean duration of infectiousness weighted by infectivity (asymptomatic individuals are less infectious than symptomatic individuals by factor $\theta_{I_A}$) for an individual in group $i$ and vaccine stage $k$, $\Delta_I^{i,k}$:

$$\Delta_I^{i,k} = \theta_{I_A}\left(1-p_C^i\right)\mathbb{E}\left[\tau_{I_A}\right] + p_C^i\left(\mathbb{E}\left[\tau_{I_P}\right] + \mathbb{E}\left[\tau_{I_{C_1}}\right]\right). \tag{268}$$

Note that $\Delta_I^{i,k}$ does not depend on $j$, as we assume the same duration spent in compartments and probability of being symptomatic between variants. The next generation matrices for variant $j$ ($j = Wildtype, Alpha, Delta, Omicron$) were calculated as,

$$NGM_{i,i'}^j(t) = m_{i,i'}(t)\xi^{i,j,0}\Delta_I^{i,0}N^{i'}, \tag{269}$$

where $\xi$ is the infectivity of an individual (fully defined in eq. (3)), $N^i$ is the total population of age group $i$, and with $R_t^j$ taken as the dominant eigenvalue of the 17 by 17 matrix $NGM^j(t)$. The element $NGM_{i,i'}^j(t)$ is therefore defined as the average number of secondary cases that an individual in age group $i'$ infected with variant $j$ at time $t$ would generate among a fully susceptible age group $i$.

The effective next generation matrices for co-circulating variants $j_1$ and $j_2$ were calculated as

$$NGM_{D(i,k),D(i',k')}^{j_1,eff}(t) = m_{i,i'}(t)\chi^{i,j_1,k}\xi^{i,j_1,k'}\Delta_I^{i,k}\left(S^{i',k'}(t) + (1-\eta_{j_1})R^{i',j_H,k'}(t)\right), \tag{270}$$

$$NGM_{D(i,k),D(i',k')}^{j_2,eff}(t) = m_{i,i'}(t)\chi^{i,j_2,k}\xi^{i,j_2,k'}\Delta_I^{i,k}\left(S^{i',k'}(t) + (1-\eta_{j_2})\left(R^{i',j_1,k'}(t) + R^{i',j_1^{reinf},k'}(t) + R^{i',j_H,k'}(t)\right)\right), \tag{271}$$

where $D: \{[0-4],[5-9],\ldots,[75-79],80+\}\times\{0,1,\ldots,6\}\to\{1,2,\ldots,119\}$ is a one-to-one mapping. Then $R_t^{j,eff}$ is taken as the dominant eigenvalue of the 119 by 119 matrix $NGM^{j,eff}(t)$ .

We calculate the reproduction numbers weighted by the two co-circulating variants $j_1$ and $j_2$ (see Section 4.5.7) as

$$R_t = \frac{w_{j_1}(t)R_t^{j_1} + w_{j_2}(t)R_t^{j_2}}{w_{j_1}(t) + w_{j_2}(t)} \tag{272}$$

$$R_t^{eff} = \frac{w_{j_1}(t)R_t^{j_1,eff} + w_{j_2}(t)R_t^{j_2,eff}}{w_{j_1}(t) + w_{j_2}(t)}, \tag{273}$$

where the weightings $w_j(t)$ are weightings based on the infectious prevalence of each variant (accounting for the baseline relative infectivity of each compartment), such that for $j = j_1, j_2$,

$$w_j(t) = \sum_i\sum_k\left(\theta_{I_A}\left(I_A^{i,j,k}(t) + I_A^{i,j^{reinf},k}(t)\right) + I_P^{i,j,k}(t) + I_P^{i,j^{reinf},k}(t) + I_{C_1}^{i,j,k}(t) + I_{C_1}^{i,j^{reinf},k}(t)\right). \tag{274}$$

#### 4.7 Basic and effective severity

We estimated the basic and effective severity by strain parametrically.

The infection hospitalisation ratio (IHR, probability that an individual infected at time $t$ will be hospitalised given pathway probabilites at time $t$) for an individual in age group $i$ and vaccine class $k$ infected with variant $j$ is derived from the model as

$$IHR^{i,j,k}(t) = p_C^i p_H^{i,j,k}(t)(1-p_{G_D}^{i,j,k}(t). \tag{275}$$

The infection hospitalisation ratio (HFR, probability that individual hospitalised at time $t$ will be die given pathway probabilites at time $t$) for an individual in age group $i$ and vaccine class $k$ infected with variant $j$ is derived from the model as

$$HFR^{i,j,k}(t) = \left(1-p_{ICU}^{i,j,k}(t)\right)p_{H_D}^{i,j,k}(t) + p_{ICU}^{i,j,k}(t)p_{ICU_D}^{i,j,k}(t) + p_{ICU}^{i,j,k}(t)\left(1-p_{ICU_D}^{i,j,k}(t)\right)p_{W_D}^{i,j,k}(t). \tag{276}$$

The infection hospitalisation ratio (IFR, probability that individual infected at time $t$ will be die given pathway probabilites at time $t$) for an individual in age group $i$ and vaccine class $k$ infected with variant $j$ is derived from the model as

$$IFR^{i,j,k}(t) = IHR^{i,j,k}(t)HFR^{i,j,k}(t) + p_C^i p_H^{i,j,k}(t) pG_D{}^{i,j,k}(t). \tag{277}$$

We then obtain overall estimates of effective severity over time by weighting by the new infections (for IFR and IHR) or new hospitalisations (for HFR) across age groups, variants and vaccine classses:

$$IHR(t) = \frac{\sum_i \sum_j \sum_k n_I^{i,j,k}(t) IHR^{i,j,k}(t)}{\sum_i \sum_j \sum_k n_I^{i,j,k}(t)} \tag{278}$$

$$HFR(t) = \frac{\sum_i \sum_j \sum_k n_H^{i,j,k}(t) HFR^{i,j,k}(t)}{\sum_i \sum_j \sum_k n_H^{i,j,k}(t)} \tag{279}$$

$$IFR(t) = \frac{\sum_i \sum_j \sum_k n_I^{i,j,k}(t) IFR^{i,j,k}(t)}{\sum_i \sum_j \sum_k n_I^{i,j,k}(t)}, \tag{280}$$

where $n_I^{i,j,k}(t)$ and $n_H^{i,j,k}(t)$ are the number of new infections and hospitalisations, respectively, at time $t$ for age group $i$, variant $j$ and vaccine class $k$.

Additionally we obtain estimates of effective severity over time for age group $i$, given by

$$IHR^i(t) = \frac{\sum_j \sum_k n_I^{i,j,k}(t) IHR^{i,j,k}(t)}{\sum_j \sum_k n_I^{i,j,k}(t)} \tag{281}$$

$$HFR^i(t) = \frac{\sum_j \sum_k n_H^{i,j,k}(t) HFR^{i,j,k}(t)}{\sum_j \sum_k n_H^{i,j,k}(t)} \tag{282}$$

$$IFR^i(t) = \frac{\sum_j \sum_k n_I^{i,j,k}(t) IFR^{i,j,k}(t)}{\sum_j \sum_k n_I^{i,j,k}(t)}, \tag{283}$$

and for variant $j \in \{j_1, j_2\}$, given by

$$IHR^j(t) = \frac{\sum_i \sum_k \left( n_I^{i,j,k}(t) IHR^{i,j,k}(t) + n_I^{i,j^{reinf},k}(t) IHR^{i,j^{reinf},k}(t) \right)}{\sum_i \sum_k \left( n_I^{i,j,k}(t) + n_I^{i,j^{reinf},k}(t) \right)} \tag{284}$$

$$HFR^j(t) = \frac{\sum_i \sum_k \left( n_H^{i,j,k}(t) HFR^{i,j,k}(t) + n_H^{i,j^{reinf},k}(t) HFR^{i,j^{reinf},k}(t) \right)}{\sum_i \sum_k \left( n_H^{i,j,k}(t) + n_H^{i,j^{reinf},k}(t) \right)} \tag{285}$$

$$IFR^j(t) = \frac{\sum_i \sum_k \left( n_I^{i,j,k}(t) IFR^{i,j,k}(t) + n_I^{i,j^{reinf},k}(t) IFR^{i,j^{reinf},k}(t) \right)}{\sum_i \sum_k \left( n_I^{i,j,k}(t) + n_I^{i,j^{reinf},k}(t) \right)}. \tag{286}$$

We defined basic severity of variant $j$ at time as the IHR, IFR, and HFR for the variant among a fully susceptible (with neither vaccine- nor infection-induced immunity) population at time $t$. For IHR and IFR, we weight across age groups for infections using the eigenvector corresponding to the leading eigenvalue of the next-generation matrix $NGM^j(t)$ (see Equation (269)), and for HFR we weight further by the probability of hospitalisation. This gives

$$IHR_0^j(t) = \frac{\sum_i w_I^i(t) IHR^{i,j,0}(t)}{\sum_i w_I^i(t)}, \tag{287}$$

$$HFR_0^j(t) = \frac{\sum_i w_I^i(t) IHR^{i,j,0}(t) HFR^{i,j,0}(t)}{\sum_i w_I^i(t) IHR^{i,j,0}(t)}, \tag{288}$$

$$IFR_0^j(t) = \frac{\sum_i w_I^i(t) IFR^{i,j,0}(t)}{\sum_i w_I^i(t)}, \tag{289}$$

where $w_I^i(t)$ are the weightings from the eigenvector.

#### 4.8 Effective population-level immunity against infection

We calculate the effective population-level immunity (EPI) in the susceptible and recovered population ($S$ and $R$ compartments) against infection with variant $j$ by weighting individuals by 1 minus their susceptibility to variant $j$ relative to an unvaccinated individual in the $S$ compartment. Thus unvaccinated individuals in the $S$ compartment are weighted 0, and we have three further types of individuals to account for

1. susceptible and vaccinated (vaccination-derived immunity only)
2. recovered and unvaccinated (infection-derived immunity only)
3. recovered and vaccinated (both vaccination- and infection-derived immunity, i.e. "hybrid immunity")

.

For the latter two, note that individuals in the $R$ compartment recovered from variants $j_1$ or $j_2$ are fully protected against infection with variant $j_1$, and those recovered from variant $j_2$ are additionally fully protected against infection with variant $j_2$.

The effective population-level immunity against infection with variant $j$ due to vaccination-derived immunity only is

$$EPI_V^j(t) = \sum_i \sum_{k>0} \left(1 - \chi^{i,j,k}(t)\right) S^{i,k}(t). \tag{290}$$

The effective population-level immunity against infection with variant $j$ due to infection-derived immunity only is

$$EPI_I^j(t) = \begin{cases} \sum_i \eta_{j_1} R^{i,j_H,0}(t) + R^{i,j_1,0}(t) + R^{i,j_1^{reinf},0}(t) + R^{i,j_2,0}(t) + R^{i,j_2^{reinf},0}(t) & \text{if } j = j_1, \\ \sum_i \eta_{j_2} \left(R^{i,j_H,0}(t) + R^{i,j_1,0}(t) + R^{i,j_1^{reinf},0}(t)\right) + R^{i,j_2,0}(t) + R^{i,j_2^{reinf},0}(t) & \text{if } j = j_2. \end{cases} \tag{291}$$

The effective population-level immunity against infection with variant $j$ due to hybrid immunity is

$$EPI_H^j(t) = \begin{cases} \sum_i \sum_{k>0} \left(1 - (1-\eta_{j_1})\chi^{i,j,k}\right) R^{i,j_H,k}(t) + R^{i,j_1,k}(t) + R^{i,j_1^{reinf},k}(t) + R^{i,j_2,k}(t) + R^{i,j_2^{reinf},k}(t) & \text{if } j = j_1, \\ \sum_i \sum_{k>0} \left(1 - (1-\eta_{j_2})\chi^{i,j_2,k}\right) \left(R^{i,j_H,k}(t) + R^{i,j_1,k}(t) + R^{i,j_1^{reinf},k}(t)\right) + R^{i,j_2,k}(t) + R^{i,j_2^{reinf},k}(t) & \text{if } j = j_2. \end{cases} \tag{292}$$

The effective population-level immunity against infection with variant $j$ is thus

$$EPI^j(t) = EPI_V^j(t) + EPI_I^j(t) + EPI_H^j(t). \tag{293}$$

For the overall effective population-level immunity against infection, we weight by the same weightings used for the variant-weighted reproduction number (see Equation (274)).

#### 4.9 Fixed parameters

We used parameter values calibrated to data from 24th February 2022. We assume that the performance of the tests (PCR and serological assays) are the same for all variants [2, 3].

| Parameter | Definition | Value | Source |
|---|---|---|---|
| $1/\gamma_R$ | Mean duration of natural immunity following infection. | 6 years | [12] |
| $p_{sero_{pos}}$ | Probability of seroconversion following infection. | 0.85 | [61] |
| $1/\gamma_{sero_{pre}}$ | Mean time to seroconversion from onset of infectiousness. | 13 days | [62] |
| $1/\gamma_{sero^1_{pos}}$ | Mean duration of seropositivity (Euroimmun assay). | 400 days | [61, 63, 64] |
| $1/\gamma_{sero^2_{pos}}$ | Mean duration of seropositivity (Roche N). | 1000 days | [61, 63, 64] |
| $p_{sero_{spec}}$ | Specificity of serology test. | 0.99 | [61] |
| $p_{sero_{sens}}$ | Sensitivity of serology test. | 1 | Assumed |
| $\eta_{Alpha}$ | Probability of cross-immunity to $Alpha$ following infection with $Wildtype$. | 0.95 | Assumed as in [3] |
| $\eta_{Delta}$ | Probability of cross-immunity to $Delta$ following infection with $Wildype$ or $Alpha$. | 0.85 | [65, 66] |
| $\eta_{Omicron}$ | Probability of cross-immunity to $Omicron$ following infection with $Wildtype$, $Delta$ or $Omicron$. | 0.25 | Assumed based on [67, 68] |
| $\theta_{I_A}$ | Infectivity of an asymptomatic individual, relative to a symptomatic individual. | 0.223 | [1] |

**Table S11:** Fixed model parameter notations, values, and evidence-base.

#### 4.10 Prior distributions

Prior distributions are described in table S12. Informative prior distributions for the single strain model are the same as prior distributions in the model given in [1, 2]. In the absence of evidence from the literature (or because existing evidence has been derived from the same datasets we use in our study), uninformative or weakly informative prior distributions have been chosen for the two-strain model; the prior for $t_{Delta}$ covers a wide period of time spanning over more than two months and the assumption of $\sigma$, and the prior for the Delta transmission advantage is assumed uniform between [0,3]."

**Table S12:** Inferred model parameter notations and prior distributions

| Parameter | Description | Prior distribution | 95% probability interval | Rationale |
|---|---|---|---|---|
| $t_{Wildtype}$ | Start date of regional outbreak (dd/mm/2020) | $U[01/01, 15/03]$ | (01/02 - 13/03) | Wide range of dates in early 2020 before our first data point |
| $t_{Alpha}$ | Alpha seeding date (dd/mm/2020) | $U[17/09/2020, 03/01/2021]$ | (17/09/2020 - 03/01/2021) | 16-week window around first Alpha case detection in England |
| $t_{Delta}$ | Delta seeding date (dd/mm/2021) | $U[08/03, 24/07]$ | (08/03 - 24/07) | 20-week window around first Delta case detection in England |
| $t_{Omicron}$ | Omicron seeding date (dd/mm/2021) | $U[14/09, 01/01]$ | (14/09/2021 - 01/01/2022) | 20-week window around first Omicron case detection in England |
| $\sigma_{Alpha/Wildtype}$ | Transmission advantage of Alpha over Wildtype | $U(0,3)$ | (0.075 - 2.925) | Wide range capturing epidemiologically plausible transmission advantages |
| $\sigma_{Delta/Alpha}$ | Tansmission advantage of Delta over Alpha | $U(0,3)$ | (0.075 - 2.925) | As above |
| $\sigma_{Omicron/Delta}$ | Transmission advantage of Omicron over Delta | $U(0,3)$ | (0.075 - 2.925) | As above |
| $\beta(t)$ | Transmission rate (pp) at $t = dd/mm/yy$ | | | |
| $\beta_1$ | 16/03/20: PM advises WFH and essential travel only | $\Gamma(136, 0.0008)$ | (0.0918,0.0128) | Range corresponding to a basic reproduction numbers between 2.5 and 3.5 consistent with [69, 70] |
| $\beta_2$ | 23/03/20: PM announces lockdown 1 | $\Gamma(3.73, 0.0154)$ | (0.0147,0.128) | Corresponding to R0 between 0.9 and 3.5, consistent with a 0% to 75% relative decrease from R0 |
| $\beta_3$ | 25/03/20: Lockdown 1 into full effect | $\Gamma(4.25, 0.0120)$ | (0.0147,0.110) | Corresponding to R0 between 0.4 and 3, allowing a further decrease in contact rates due to NPIs |
| $\beta_4$ | 11/05/20: Initial easing of lockdown 1 | $\Gamma(4.25, 0.0120)$ | (0.0147,0.110) | As above |
| $\beta_5$ | 15/06/20: Non-essential shops re-open | $\Gamma(4.25, 0.0120)$ | (0.0147,0.110) | As above |
| $\beta_6$ | 04/07/20: Hospitality re-opens | $\Gamma(4.25, 0.0120)$ | (0.0147,0.110) | As above |
| $\beta_7$ | 01/08/20: "Eat out to help out" scheme starts | $\Gamma(4.25, 0.0120)$ | (0.0147,0.110) | As above |
| $\beta_8$ | 01/09/20: Schools and universities re-open | $\Gamma(4.25, 0.0120)$ | (0.0147,0.110) | As above |
| $\beta_9$ | 14/09/20: "Rule of six" introduced | $\Gamma(4.25, 0.0120)$ | (0.0147,0.110) | As above |
| $\beta_{10}$ | 14/10/20: Tiered system introduced | $\Gamma(4.25, 0.0120)$ | (0.0147,0.110) | As above |
| $\beta_{11}$ | 31/10/20: Lockdown 2 announced | $\Gamma(4.25, 0.0120)$ | (0.0147,0.110) | As above |
| $\beta_{12}$ | 05/11/20: Lockdown 2 starts | $\Gamma(4.25, 0.0120)$ | (0.0147,0.110) | As above |
| $\beta_{13}$ | 02/12/20: Lockdown 2 ends | $\Gamma(4.25, 0.0120)$ | (0.0147,0.110) | As above |
| $\beta_{14}$ | 18/12/20: School holidays start | $\Gamma(4.25, 0.0120)$ | (0.0147,0.110) | As above |
| $\beta_{15}$ | 25/12/20: Last day of holiday season relaxation | $\Gamma(4.25, 0.0120)$ | (0.0147,0.110) | As above |
| $\beta_{16}$ | 05/01/21: Lockdown 3 starts | $\Gamma(4.25, 0.0120)$ | (0.0147,0.110) | As above |
| $\beta_{17}$ | 08/03/21: Roadmap step one - schools reopen | $\Gamma(4.25, 0.0120)$ | (0.0147,0.110) | As above |
| $\beta_{18}$ | 01/04/21: School holidays | $\Gamma(4.25, 0.0120)$ | (0.0147,0.110) | As above |
| $\beta_{19}$ | 19/04/21: Roadmap step two - outdoor rule of 6 (12/04) and schools re-open (19/04) | $\Gamma(4.25, 0.0120)$ | (0.0147,0.110) | As above |
| $\beta_{20}$ | 17/05/21: Roadmap step three - Indoor hospitality opens | $\Gamma(4.25, 0.0120)$ | (0.0147,0.110) | As above |
| $\beta_{21}$ | 21/06/21: Wedding and care home restrictions eased | $\Gamma(4.25, 0.0120)$ | (0.0147,0.110) | As above |
| $\beta_{22}$ | 03/07/21: Euro 2020 quarter finals (cited as significant influence [71]) | $\Gamma(4.25, 0.0120)$ | (0.0147,0.110) | As above |
| $\beta_{23}$ | 11/07/21: End of Euros football tournament | $\Gamma(4.25, 0.0120)$ | (0.0147,0.110) | As above |
| $\beta_{24}$ | 19/07/21: Full lift of NPIs | $\Gamma(4.25, 0.0120)$ | (0.0147,0.110) | As above |
| $\beta_{25}$ | 15/08/21: Summer festivals/holidays | $\Gamma(4.25, 0.0120)$ | (0.0147,0.110) | As above |
| $\beta_{26}$ | 01/09/21: Schools return | $\Gamma(4.25, 0.0120)$ | (0.0147,0.110) | As above |
| $\beta_{27}$ | 22/09/21: Mid-point between school start and half term | $\Gamma(4.25, 0.0120)$ | (0.0147,0.110) | As above |

Continued on next page

**Table S12 – continued from previous page**

| | Description | Prior distribution | 95% probability interval | Rationale |
|---|---|---|---|---|
| $\beta_{28}$ | 01/10/21: Point before sharp increase in epidemic wave | $\Gamma(4.25, 0.0120)$ | (0.0147,0.110) | As above |
| $\beta_{29}$ | 22/10/21: School holidays (half term) | $\Gamma(4.25, 0.0120)$ | (0.0147,0.110) | As above |
| $\beta_{30}$ | 01/11/21: School return | $\Gamma(4.25, 0.0120)$ | (0.0147,0.110) | As above |
| $\beta_{31}$ | 08/12/21: Plan B announced | $\Gamma(4.25, 0.0120)$ | (0.0147,0.110) | As above |
| $\beta_{32}$ | 23/12/21: School holidays start | $\Gamma(4.25, 0.0120)$ | (0.0147,0.110) | As above |
| $\beta_{33}$ | 04/01/22: Schools return | $\Gamma(4.25, 0.0120)$ | (0.0147,0.110) | As above |
| $\beta_{34}$ | 19/01/22: Announcement of end of Plan B | $\Gamma(4.25, 0.0120)$ | (0.0147,0.110) | As above |
| $\beta_{35}$ | 27/01/22: End of Plan B | $\Gamma(4.25, 0.0120)$ | (0.0147,0.110) | As above |
| $\beta_{36}$ | 24/02/22: End of self-isolation policy in England, end of fits | $\Gamma(4.25, 0.0120)$ | (0.0147,0.110) | As above |
| $p^{max}_{H,1}$, $p^{max}_{H,2}$ | The probability of symptomatic individuals developing serious disease requiring hospitalisation, for the group with the largest probability at different timepoints (see Section 4.4.3) | $U(0,1)$ | (0.025,0.975) | Uninformative |
| $p^{max}_{G_D,1}$, $p^{max}_{G_D,2}$ | Probability of death in the community given disease severe enough for hospitalisation for the group with the largest probability at different timepoints (see Section 4.4.3) | $U(0,1)$ | (0.025,0.975) | Uninformative |
| $p^{max}_{ICU,1}$ | Probability of triage to ICU for new hospital admissions, for the group with the largest probability at different timepoints (see Section 4.4.3) | $B(13.9, 43.9)$ | (0.140,0.357) | Informed by previous work [2, 1] based on [51] |
| $p^{max}_{ICU,2}$ | Probability of triage to ICU for new hospital admissions, for the group with the largest probability at different timepoints (see Section 4.4.3) | $U(0,1)$ | (0.025,0.975) | Uninformative |
| $p^{max}_{H_D}$ | Initial probability of death for general inpatients | $B(42.1, 50.1)$ | (0.356,0.558) | Informed by previous work [2, 1] based on [51] |
| $p^{max}_{ICU_D}$ | Initial probability of death for ICU inpatients | $B(60.2, 29.3)$ | (0.573,0.766) | Informed by previous work [2, 1] based on [51] |
| $p^{max}_{W_D}$ | Initial probability of death for stepdown inpatients | $B(28.7, 52.1)$ | (0.255,0.462) | Informed by previous work [2, 1] based on [51] |
| $\mu_{D,1}, \mu_{D,2}, \mu_{D,3}, \mu_{D,4}, \mu_{D,5}$ | Hospital mortality multipliers due to changes in clinical care at different timepoints (see Section 4.4.3) | $U(0,1)$ | (0.025,0.975) | Uninformative |
| $\pi_H^{Alpha/Wildtype}$ | Multiplier of the probability of hospitalisation with Alpha relative to the Wildtype variant (see Section 4.4.3) | $U(0,3)$ | (0.075,2.925) | Wide range capturing epidemiologically plausible intrinsic changes in the severity of SARS-CoV-2 |
| $\pi_H^{Delta/Alpha}$ | Multiplier of the probability of hospitalisation with Delta relative to the Alpha variant (see Section 4.4.3) | $U(0,3)$ | (0.075,2.925) | As Above |
| $\pi_H^{Omicron/Delta}$ | Multiplier of the probability of hospitalisation with Omicron relative to the Delta variant (see Section 4.4.3) | $U(0,3)$ | (0.075,2.925) | As Above |

Continued on next page

**Table S12 – continued from previous page**

| | Description | Prior distribution | 95% probability interval | Rationale |
|---|---|---|---|---|
| $\pi_{ICU}^{Alpha/Wildtype}$ | Multiplier of the probability of ICU admission with Alpha relative to the Wildtype variant (see Section 4.4.3) | $U(0,3)$ | (0.075,2.925) | Wide range capturing epidemiologically plausible intrinsic changes in the severity of SARS-CoV-2 |
| $\pi_{ICU}^{Delta/Alpha}$ | Multiplier of the probability of ICU admission with Delta relative to the Alpha variant (see Section 4.4.3) | $U(0,3)$ | (0.075,2.925) | As Above |
| $\pi_{ICU}^{Omicron/Delta}$ | Multiplier of the probability of ICU admission with Omicron relative to the Delta variant (see Section 4.4.3) | $U(0,3)$ | (0.075,2.925) | As Above |
| $\pi_{D}^{Alpha/Wildtype}$ | Multiplier of the probability of death with Alpha relative to the Wildtype variant (see Section 4.4.3) | $U(0,3)$ | (0.075,2.925) | Wide range capturing epidemiologically plausible intrinsic changes in the severity of SARS-CoV-2 |
| $\pi_{D}^{Delta/Alpha}$ | Multiplier of the probability of death with Delta relative to the Alpha variant (see Section 4.4.3) | $U(0,3)$ | (0.075,2.925) | As Above |
| $\pi_{D}^{Omicron/Delta}$ | Multiplier of the probability of death with Omicron relative to the Delta variant (see Section 4.4.3) | $U(0,3)$ | (0.075,2.925) | As Above |
| $\mu_{\gamma_H,1}$, $\mu_{\gamma_H,2}$, $\mu_{\gamma_H,3}$, $\mu_{\gamma_H,4}$ | Mean duration multipliers for non-ICU hospital compartments at different timepoints (see Section 4.4.3) | $U(0,2)$ | $(0.050, 1.950)$ | Wide range capturing epidemiologically plausible relative changes in durations |
| $p_{NC}^{[15,25)}$, $p_{NC}^{[25,50)}$, $p_{NC}^{[50,65)}$, $p_{NC}^{[65,80)}$, $p_{NC}^{[80+)}$ | Prevalence of non-COVID symptomatic illness that could lead to getting a PCR test for different age bands | $U(0,1)$ | (0.025,0.975) | Uninformative |
| $p_{NCW}^{[15,25)}$, $p_{NCW}^{[24,50)}$, $p_{NCW}^{[50,65)}$, $p_{NCW}^{[65,80)}$, $p_{NCW}^{[80+)}$ | Prevalence of non-COVID symptomatic illness that could lead to getting a PCR test on a weekend for different age bands | $U(0,1)$ | (0.025,0.975) | Uninformative |
| $\rho_{P2_{test}}$ | Overdispersion of PCR positivity | $U(0,1)$ | (0.025,0.975) | Uninformative |
| $\alpha_A$ | Overdispersion for hospital admission data streams | $U(0,1)$ | (0.025,0.975) | Uninformative |
| $\alpha_H$ | Overdispersion for hospital bed streams | $U(0,1)$ | (0.025,0.975) | Uninformative |
| $\alpha_{D_{hosp}}$ | Overdispersion for hospital death data streams | $U(0,1)$ | (0.025,0.975) | Uninformative |
| $\alpha_{D_{comm}}$ | Overdispersion for community death data streams | $U(0,1)$ | (0.025,0.975) | Uninformative |

#### 4.11 Running the model

The model is fitted to multiple data streams up to 24th February 2022, capturing the entirety of the SARS-CoV-2 epidemic up to the official end of the policy for self-isolation in England [72]. The model is run under baseline assumptions reflected in our fixed (Table S11) and VE parameters (Table S4).

Before running the pMCMC, we pre-tune the model by running a traditional MCMC on the equivalent "expectation model", defined as the same model but wherever a random draw arises, the mean of the corresponding distribution is used instead, thereby allowing compartments to take non-integer values. This was done to optimise computational efficiency in exploring the multi-dimensional parameter space across fitted parameters (figure S33, figure S34, figure S35 and figure S36). The "expectation" model is coupled with an adaptive MCMC to accelerate parameter space exploration.

We then use the pre-tuned parameter set from the "expectation" model with the highest posterior and variance-covariance (VCV) matrix of the posterior distribution parameters as the initial values and proposal kernel, respectively, for subsequent pMCMC runs. The latter uses sequential Monte Carlo estimates of the latent variables with a bootstrap particle filter. This avoided both the need for data augmentation techniques and to estimate latent variables (i.e. unobserved states) alongside fitted parameters.

At each iteration of the pMCMC, we randomly rerun the particle filter on the current parameter set with probability $\frac{1}{100}$ to get a new marginal likelihood estimate, which prevents chains from getting stuck at a particular parameter due to a high likelihood estimate [73, 74]. We run 4 chains with 192 particles in this process over 5,000 pMCMC iterations, of which 1000 are discarded as burn-in. We thin the combined sample uniformly to achieve a posterior sample size of 1000. To test the robustness of our choice of particle number, we also ran our model with double the number of particles (384) for our central assumptions (table S23), which comes at approximately double the run-time cost. Given results were generally similar, we are confident that using 192 particles consistently for our central assumption fits and all sensitivity analyses provides robust results in a more reasonable run-time.

Our final pMCMC model fit was produced through approximately 23,000 CPU hours, spread across 7 nodes of 32-core Xeons (dual 16-core 2.6 GHz). Implementation of the model described above is fully described in FitzJohn et al. [75]. The primary interface to the model is coded in R [76] with functions written in packages `sircovid` and `spimalot`. The model is written in `odin` and run with `dust`, the pMCMC functions are written in `mcstate`.

For this paper we used `sircovid` v0.14.12, `spimalot` v0.8.23, `dust` v0.14.1, and `mcstate` v0.9.15. The above packages are publicly available in the *mrc-ide* GitHub organisation (https://github.com/mrc-ide/). The code and scripts used to create the results in this paper are available in (https://github.com/mrc-ide/sarscov2-severity-england).

### 5 Sensitivity Analyses

The previous sections provided methodological details of our model structure and how we fit to existing epidemiological data. This section outlines the sensitivity analyses we ran to explore uncertainties around fixed model parameters during the period fitting to data from the Omicron variant given:

1. *Cross-immunity against infection.*
2. *Cross-immunity against hospitalisation.*
3. *Cross-immunity against death.*
4. *Efficacy of boosters against severe disease and death.*
5. *Mean serial interval duration.*

Table S13 summarises cross-immunity sensitivity analyses explored, and table S16 the booster VE against Omicron. In previous publications from our group [1, 2, 3], we have presented sensitivity analyses around VE, and the time to waning of natural and vaccine-induced immunity for the Alpha and Delta variants, so these are not repeated herein.

| | *Analysis* | *Description* | *Source* |
|---|---|---|---|
| *1* | *Cross-immunity against infection* | We assume a 25% protection against infection if infected with Omicron, given natural immunity (recovery from infection with Delta or other historic variant). We explored a higher and a lower value for this assumption, at 35% and 20%, respectively. | Assumed |
| *2* | *Cross-immunity against hospitalisation* | We assume a 55% protection against hospitalisation if infected with Omicron, given natural immunity (recovery from infection with Delta or other historic variant). We explored a higher and a lower value for this assumption, at 63% and 48%, respectively. | [77] |
| *3* | *Cross-immunity against death* | We assume an 18% protection against death if infected with Omicron, given natural immunity (recovery from infection with Delta or other historic variant). We explored a higher and a lower value for this assumption, at 57% and 6%, respectively. | [77] |
| *4* | *Vaccine efficacy* | We explored a plausible range of uncertainty in our central VE parameters. For the case of VE *vs* the Alpha and Delta variants, we assumed +/-10% for all VE parameters as we have included a wealth of data to derive central parameters (see Table S4). Given greater uncertainty around VE of second and booster doses *vs* the Omicron variant ($V_3$ to $V_6$ strata), we explored the upper and lower bound from available literature. See Table S14, Table S15 and Table S16 for specific parameters. | Assumption and [41] |
| *5* | *Alternative* $\mu_D$ | We explored two alternative parameterisations of the $\mu_D$ piecewise linear function (see section 4.4.4), varying change points around the point of emergence of the Alpha and Delta variants. | Assumed |
| *6* | *Serial interval duration* | In our central parameters, we assume a decreasing SI for each variant in succession, compared to the Wildtype variant (Section 2.2). We fitted our model assuming a fixed SI of 5.2 days (Wildtype variant-like) and of 3.9 days (Omicron variant-like) for all variants. | Assumed |

**Table S13:** Summary of sensitivity analyses explored

For vaccine efficacy, our central model parameters were as defined in section 3.1. Values for vaccine effectiveness (VE) are derived from both vaccine efficacy measured in clinical trials and vaccine effectiveness studies (Table S4). Where possible, data from the UK have been used and represent effectiveness of dosing schedules with an 12 week gap between doses. We assumed that there are no significant differences in vaccine effectiveness by age, sex, or underlying health conditions [28, 29]. As previously describe for our model [2, 3], we assume that vaccine protection against symptomatic disease also provides a similar level of protection against infection and that, in those individuals who do become infected after vaccination, onward transmission is also reduced [30].

### 5.1 Vaccine efficacy sensitivity analysis

Table S14, table S15 and table S16 summarise VE values used for each scenario. Please see comparison of inferred intrinsic transsibility and basic severity for each scenario, compared to central parameters, in figure S8.

| | | Central | | Lower VE | | Higher VE | | Informed by |
|---|---|---|---|---|---|---|---|---|
| End point | Dose | AZ | PF/Mod | AZ | PF/Mod | AZ | PF/Mod | |
| Death | 1 | 88% | 89% | 79% | 80% | 96% | 98% | Assumed -10% lower, +10% higher VE. |
| | 2 (Full protection) | 99% | 99% | 89% | 89% | 99% | 99% | |
| | 2 (Waned protection) | 83% | 90% | 75% | 81% | 91% | 99% | |
| Severe disease | 1 | 81% | 89% | 73% | 80% | 89% | 98% | Assumed -10% lower, +10% higher VE. |
| | 2 (Full protection) | 99% | 99% | 89% | 89% | 99% | 99% | |
| | 2 (Waned protection) | 77% | 90% | 70% | 81% | 85% | 99% | |
| Mild disease or infection | 1 | 64% | 79% | 58% | 71% | 70% | 87% | Assumed -10% lower, +10% higher VE. |
| | 2 (Full protection) | 92% | 99% | 83% | 89% | 99% | 99% | |
| | 2 (Waned protection) | 29% | 77% | 26% | 70% | 32% | 85% | |
| Transmission | 1 | 45% | 45% | 41% | 41% | 50% | 50% | Assumed -10% lower, +10% higher VE. |
| | 2 (Full protection) | 45% | 45% | 41% | 41% | 50% | 50% | |
| | 2 (Waned protection) | 40% | 40% | 36% | 36% | 44% | 44% | |

**Table S14:** Sensitivity analysis parameters for vaccine efficacy *vs* the Alpha variant. "Infection" refers to vaccine effectiveness protecting an individual from being infected with SARS-CoV-2, whilst "transmission" refers to the vaccine effectiveness at preventing onward transmission by an infected individual.

| | | Central | | Lower VE | | Higher VE | | Informed by |
|---|---|---|---|---|---|---|---|---|
| End point | Dose | AZ | PF/Mod | AZ | PF/Mod | AZ | PF/Mod | |
| Death | 1 | 87% | 89% | 78% | 80% | 96% | 98% | Assumed -10% lower, +10% higher VE. |
| | 2 (Full protection) | 99% | 99% | 89% | 89% | 99% | 99% | |
| | 2 (Waned protection) | 82% | 90% | 74% | 81% | 90% | 99% | |
| | 3 (Full protection) | 99% | 99% | 89% | 89% | 99% | 99% | |
| | 3 (Waned protection) | 89% | 89% | 81% | 81% | 99% | 99% | |
| Severe disease | 1 | 81% | 89% | 73% | 80% | 89% | 98% | Assumed -10% lower, +10% higher VE. |
| | 2 (Full protection) | 99% | 99% | 89% | 89% | 99% | 99% | |
| | 2 (Waned protection) | 77% | 90% | 70% | 81% | 85% | 99% | |
| | 3 (Full protection) | 99% | 99% | 89% | 89% | 99% | 99% | |
| | 3 (Waned protection) | 89% | 89% | 81% | 81% | 99% | 99% | |
| Mild disease or infection | 1 | 51% | 51% | 47% | 47% | 57% | 57% | Assumed -10% lower, +10% higher VE. |
| | 2 (Full protection) | 87% | 95% | 79% | 86% | 96% | 99% | |
| | 2 (Waned protection) | 19% | 49% | 18% | 44% | 21% | 54% | |
| | 3 (Full protection) | 92% | 92% | 83% | 83% | 99% | 99% | |
| | 3 (Waned protection) | 36% | 36% | 39% | 39% | 54% | 54% | |
| Transmission | 1 | 33% | 33% | 30% | 30% | 36% | 36% | Assumed -10% lower, +10% higher VE. |
| | 2 (Full protection) | 40% | 40% | 36% | 36% | 44% | 44% | |
| | 2 (Waned protection) | 19% | 19% | 17% | 17% | 21% | 21% | |
| | 3 (Full protection) | 40% | 40% | 36% | 36% | 44% | 44% | |
| | 3 (Waned protection) | 19% | 19% | 17% | 17% | 21% | 21% | |

**Table S15:** Sensitivity analysis parameters for vaccine efficacy *vs* the Delta variant. "Infection" refers to vaccine effectiveness protecting an individual from being infected with SARS-CoV-2, whilst "transmission" refers to the vaccine effectiveness at preventing onward transmission by an infected individual.

| | | Central | | Lower VE | | Higher VE | | Informed by |
|---|---|---|---|---|---|---|---|---|
| End point | Dose | AZ | PF/Mod | AZ | PF/Mod | AZ | PF/Mod | |
| Death | 2 (Full protection) | 97% | 97% | 80% | 80% | 97% | 97% | Low and high values from Gov [41] |
| | 2 (Waned protection) | 56% | 56% | 40% | 40% | 70% | 70% | |
| | 3 (Full protection) | 96% | 96% | 85% | 85% | 96% | 96% | |
| | 3 (Waned protection) | 62% | 62% | 50% | 50% | 80% | 80% | |
| Severe disease | 2 (Full protection) | 97% | 97% | 80% | 80% | 97% | 97% | Low and high values from Gov [41] |
| | 2 (Waned protection) | 56% | 56% | 40% | 40% | 65% | 65% | |
| | 3 (Full protection) | 96% | 96% | 85% | 85% | 96% | 96% | |
| | 3 (Waned protection) | 62% | 62% | 40% | 40% | 75% | 75% | |
| Mild disease or infection | 2 (Full protection) | 41% | 60% | 30% | 30% | 50% | 65% | Low and high values from Gov [41] |
| | 2 (Waned protection) | 0% | 0% | 0% | 0% | 5% | 15% | |
| | 3 (Full protection) | 72% | 74% | 60% | 65% | 72% | 74% | |
| | 3 (Waned protection) | 0% | 0% | 0% | 0% | 20% | 20% | |
| Transmission | 2 (Full protection) | 40% | 40% | 29% | 29% | 40% | 40% | Assumed lower than for mild disease |
| | 2 (Waned protection) | 0% | 0% | 0% | 0% | 3% | 8% | |
| | 3 (Full protection) | 40% | 40% | 30% | 30% | 50% | 50% | |
| | 3 (Waned protection) | 0% | 0% | 0% | 0% | 10% | 10% | |

**Table S16:** Sensitivity analysis parameters for vaccine effectiveness against the Omicron variant for AstraZeneca (AZ), Pfizer (PF), and Moderna (Mod) by vaccine dose. "Infection" refers to vaccine effectiveness protecting an individual from being infected with SARS-CoV-2, whilst "transmission" refers to the vaccine effectiveness at preventing onward transmission by an infected individual.

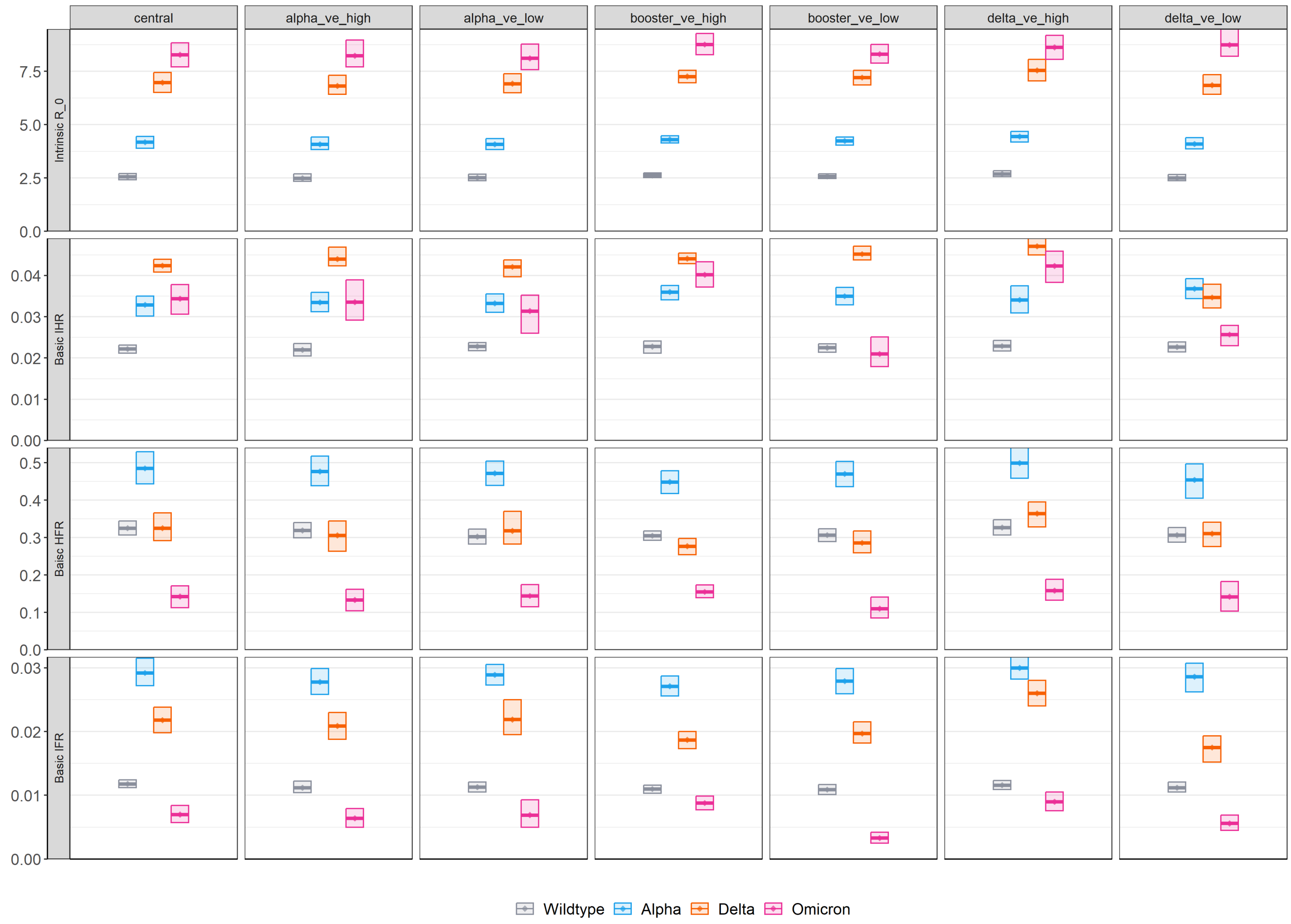

**Figure S8:** Sensitivity analysis of model inferred intrinsic $R_0$ and basic IHR, HFR and IFR of the variants. From left to right: Central, high and low vaccine efficacy (1st and 2nd dose, table S14) against the Alpha variant, high and low vaccine efficacy (1st, 2nd and booster, table S12) against the Delta variant, and high and low vaccine efficacy (2nd and booster dose only table S16) against the Omicron variant. Box plots show mean model-inferred properties and 95% CrI for the Wildtype (grey), Alpha (blue), Delta (orange) and Omicron (pink) variants.

### 5.2 Cross-immunity sensitivity analysis

We varied central cross-immunity parameters of conferred by prior infection with a historic variant *vs* an emerging variant (e.g. prior infection with Alpha or Delta *vs* Omicron variant). Each of cross-immunity against infection, hospitalisation or death was varied in a one-way analysis, simultaniously for all variants. For specific values and literature sources of parameters (where available) see table S13.

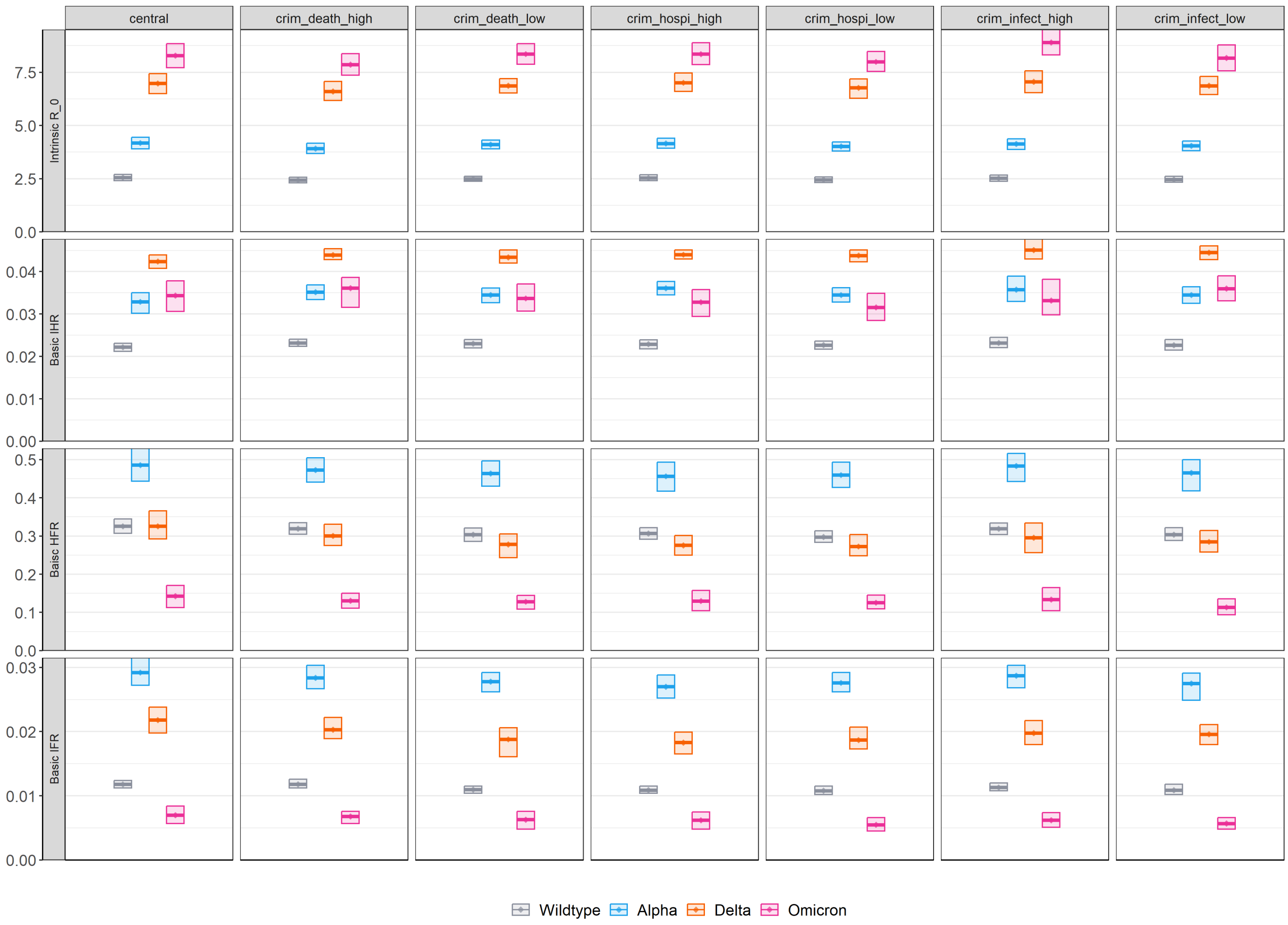

**Figure S9:** Sensitivity analysis for scenario description) of model inferred intrinsic $R_0$ and basic IHR, HFR and IFR of the variants. From left to right: high and low cross-immunity against death, high and low cross-immunity against hospitalisation, and high and low cross-immunity against infection (see values in table S13). Box plots show mean model-inferred properties and 95% CrI for the Wildtype (grey), Alpha (blue), Delta (orange) and Omicron (pink) variants.

#### 5.3 Other sensitivity analysis

Our model allowed fitting time-varying changes in severity, independent of the inferred basic properties of the variants. We explored alternative parameterisations of the piece-wise linear function for changing $\mu_D(t)$ (see details in section 4.4.4. In these scenarios, we specifically sought to infer whether the choice of change points influenced the inferred relative severity between the Alpha and Delta variants. Additionally, we varied our central serial interval parameters, by either fixing it at its highest (i.e. all variants with same value as Wildtype) and lowest (i.e. all variants with same value as Omicron BA.1) values.

| Scenario | Dates | Rationale |
|---|---|---|
| Central | 01-04-2020<br>01-07-2020 to 15-09-2020<br>15-10-2020 to 01-12-2020<br>04-02-2021<br>01-04-2021 to 04-11-2021<br>31-12-2021 | See table S10 |
| $\mu_D(t) winter$ | 01-04-2020<br>01-07-2020 to 15-09-2020<br>01-12-2020<br>04-02-2021<br>01-04-2021 to 04-11-2021<br>31-12-2021 | Explore a linear change in $\mu_D(t)$ from 09-2020 to 12-2020 to infer potential changes in severity given mounting healthcare demands around the date of Alpha emergence. |
| $\mu_D(t) summer$ | 01-04-2020<br>01-07-2020 to 15-09-2020<br>15-10-2020 to 01-12-2020<br>04-02-2021<br>01-04-2021 to 01-06-2021<br>04-11-2021<br>31-12-2021 | Explore flat period in $\mu_D(t)$ between 04-2021 and 06-2021 during the period of Delta emergence, then linear change from 06-2021 to 11-2021 as Delta was dominant during a period of potential evolving hospital admission and triaging thresholds. |

**Table S17:** Sensitivity analysis of fitted change points for the time-varying $\mu_D(t)$.

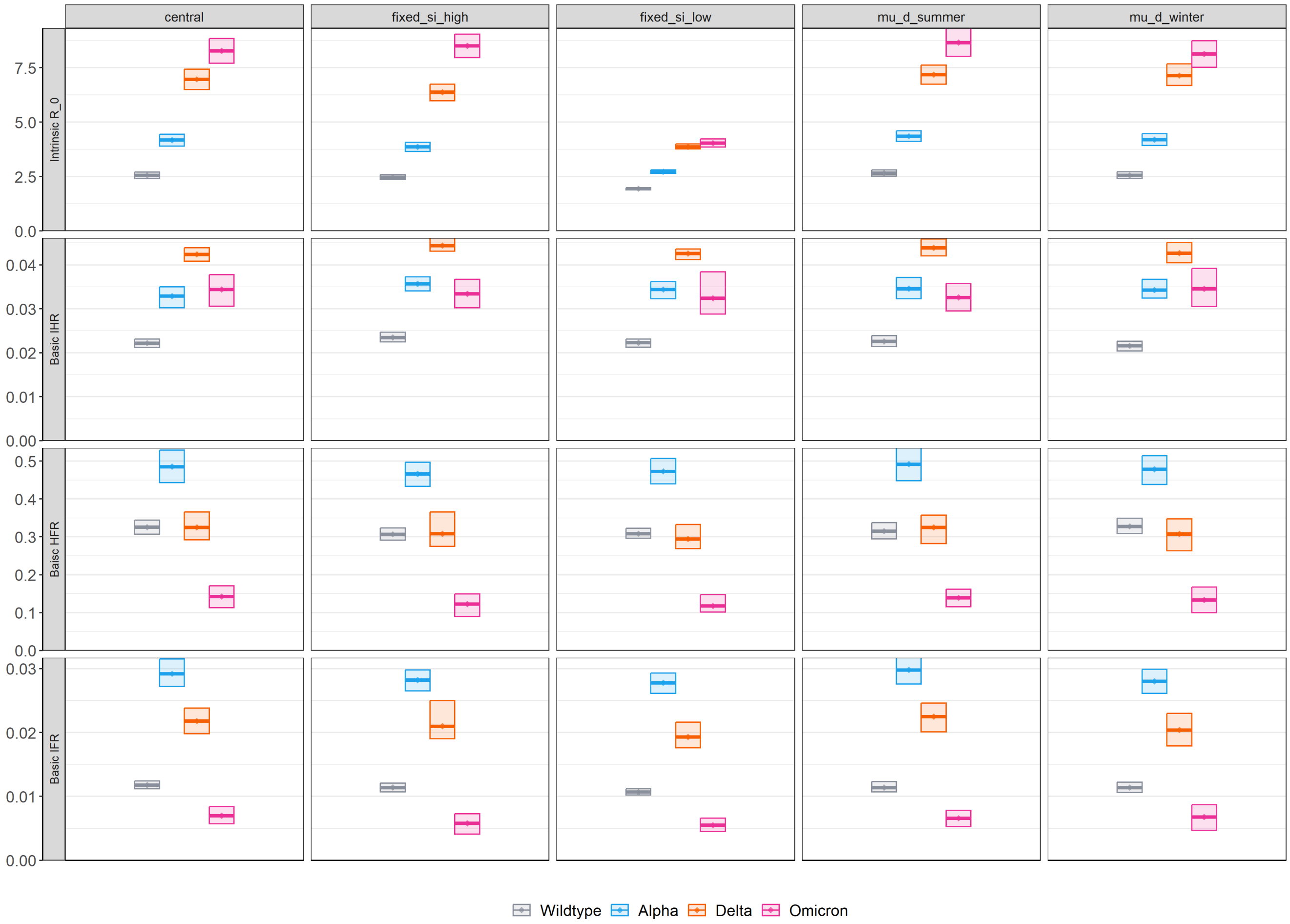

**Figure S10:** Sensitivity analysis of model inferred intrinsic $R_0$ and basic IHR, HFR and IFR of the variants. From left to right: Central, high and low fixed serial interval duration (see values in table S13), and alternative changepoints for $\mu_D(t)$ in the winter of 2020/21 and in the summer of 2021 (see values and rationale in table S17). Box plots show mean model-inferred properties and 95% CrI for the Wildtype (grey), Alpha (blue), Delta (orange) and Omicron (pink) variants.

### 6 Supplementary Results

A limitation of our approach is that there could be identifiability issues in model-inferred parameters of the relative severity of the variants. Contrary to the rich genomic surveillance data representative of cases diagnosed in the community (Pillar 2), which we used to fit our model to variant replacement events and infer differences in the relative transmissibility of the variants, there was not a similarly representative genomic surveillance dataset for hospital admissions and deaths. Therefore, we could not fit to or compare our modelled results to empiric data of hospital admissions and deaths by variant status to further underpin our inference around relative severity of the variants.

In particular, the above could mean that inferred statistically significant differences in the relative severity of one variant compared to another could have been driven by confounding factors. In particular, there could have been underlying differences in vaccine-derived protection against severe disease or death in the population, or given healthcare system performance variations at the time when each circulated, driving changes in severity rather than the variants themselves.

To support the robustness of our results and minimise the risk of wrongly attributing differences in variant severity to such confounding factors, we conducted the below supplementary analyses. Future similar analyses of pandemic emergencies would greatly benefit from representative genomic surveillance performed across all levels of the severity pathways of disease, including hospitalisations and deaths.

#### 6.1 Relative risk of transmissibility and severity of the variants

Tables below summarise the relative risk of transmissibility and severity comparing across all variants. We performed paired sampling of model-inferred relative transmissibility and severity parameters within pMCMC steps. We did this to ensure 95%CrI of results below reflected only uncertainty from parameter uncertainty, void of stochastic variance.

| | Wildtype | Alpha | Delta | Omicron |
|---|---|---|---|---|
| Wildtype | - | 1.63 (1.60 - 1.67) | 2.73 (2.66 - 2.79) | 3.24 (3.14 - 3.35) |
| Alpha | 0.61 (0.60 - 0.62) | - | 1.67 (1.62 - 1.71) | 1.98 (1.91 - 2.05) |
| Delta | 0.37 (0.36 - 0.38) | 0.60 (0.58 - 0.62) | - | 1.19 (1.15 - 1.22) |
| Omicron | 0.31 (0.30 - 0.32) | 0.50 (0.49 - 0.52) | 0.84 (0.82 - 0.87) | - |

**Table S18:** Relative intrinsic transmissibility risk (mean, 95%CrI) across modelled variants, calculated as $R_0^j$ / $R_0^i$. Note here indices $i$ and $j$ refer to table row and column, respectively.

| | Wildtype | Alpha | Delta | Omicron |
|---|---|---|---|---|
| Wildtype | - | 1.48 (1.39 - 1.57) | 1.91 (1.80 - 2.04) | 1.56 (1.36 - 1.75) |
| Alpha | 0.67 (0.64 - 0.72) | - | 1.29 (1.20 - 1.42) | 1.05 (0.92 - 1.21) |
| Delta | 0.52 (0.49 - 0.56) | 0.78 (0.70 - 0.84) | - | 0.81 (0.72 - 0.89) |
| Omicron | 0.65 (0.57 - 0.73) | 0.96 (0.83 - 1.09) | 1.23 (1.12 - 1.38) | - |

**Table S19:** Relative basic IHR risk (mean, 95%CrI) across modelled variants, calculated as $IHR^j$ / $IHR^i$. Note here indices $i$ and $j$ refer to table row and column, respectively.

| | Wildtype | Alpha | Delta | Omicron |
|---|---|---|---|---|
| Wildtype | - | 1.49 (1.40 - 1.60) | 1.00 (0.88 - 1.15) | 0.44 (0.34 - 0.53) |
| Alpha | 0.67 (0.62 - 0.72) | - | 0.67 (0.58 - 0.77) | 0.29 (0.23 - 0.36) |
| Delta | 1.00 (0.87 - 1.14) | 1.50 (1.29 - 1.72) | - | 0.44 (0.36 - 0.52) |
| Omicron | 2.31 (1.88 - 2.92) | 3.44 (2.81 - 4.27) | 2.30 (1.93 - 2.77) | - |

**Table S20:** Relative basic HFR risk (mean, 95%CrI) across modelled variants, calculated as $HFR^j$ / $HFR^i$. Note here indices $i$ and $j$ refer to table row and column, respectively.

| | Wildtype | Alpha | Delta | Omicron |
|---|---|---|---|---|
| Wildtype | - | 2.48 (2.32 - 2.64) | 1.84 (1.65 - 2.03) | 0.59 (0.47 - 0.72) |
| Alpha | 0.40 (0.38 - 0.43) | - | 0.74 (0.65 - 0.84) | 0.24 (0.20 - 0.29) |
| Delta | 0.54 (0.49 - 0.61) | 1.35 (1.20 - 1.53) | - | 0.32 (0.26 - 0.38) |
| Omicron | 1.70 (1.39 - 2.11) | 4.21 (3.48 - 5.12) | 3.13 (2.66 - 3.80) | - |

**Table S21:** Relative basic IFR risk (mean, 95%CrI) across modelled variants, calculated as $IFR^j$ / $IFR^i$. Note here indices $i$ and $j$ refer to table row and column, respectively.

### 6.2 Winter 2020/21 HFR and mechanical ventilation bed occupancy

To further characterise the model inferred increase in effective HFR during the winter of 2020-2021 and the role of Alpha emergence *vs* healthcare performance as contributing factors, we analysed linked patient-level data on PCR-positive COVID-19 cases, hospital admissions and deaths between September 1, 2020 and February 28, 2021. We counted hospitalisations for patients staying in hospital for 24 hours after their date of admission, which was defined as either the date of first hospital attendance within 14 days of a positive PCR test in the community or the date of PCR-positive diagnosis for patients already in hospital. We then calculated the a time series of daily HFR as the number of hospital admissions that (prospectively) had an outcome of death over the total number of admissions for that day. Lastly, we used daily mechanical ventilation (MV) bed occupancy as a proxy of hospital pressures, defined as the number of MV beds occupied, divided by the total number of such beds.

We logit transformed occupancy and HFR and assessed the correlation between them in simple linear regression, adjusting for a potential interaction between occupancy and variant dominance. The latter was defined as a binary variable for a given day, as either "Alpha dominant" day or not, if the frequency of this variant in community positive PCR was greater than 60%. Results are presented below in figure S11 and table S22.

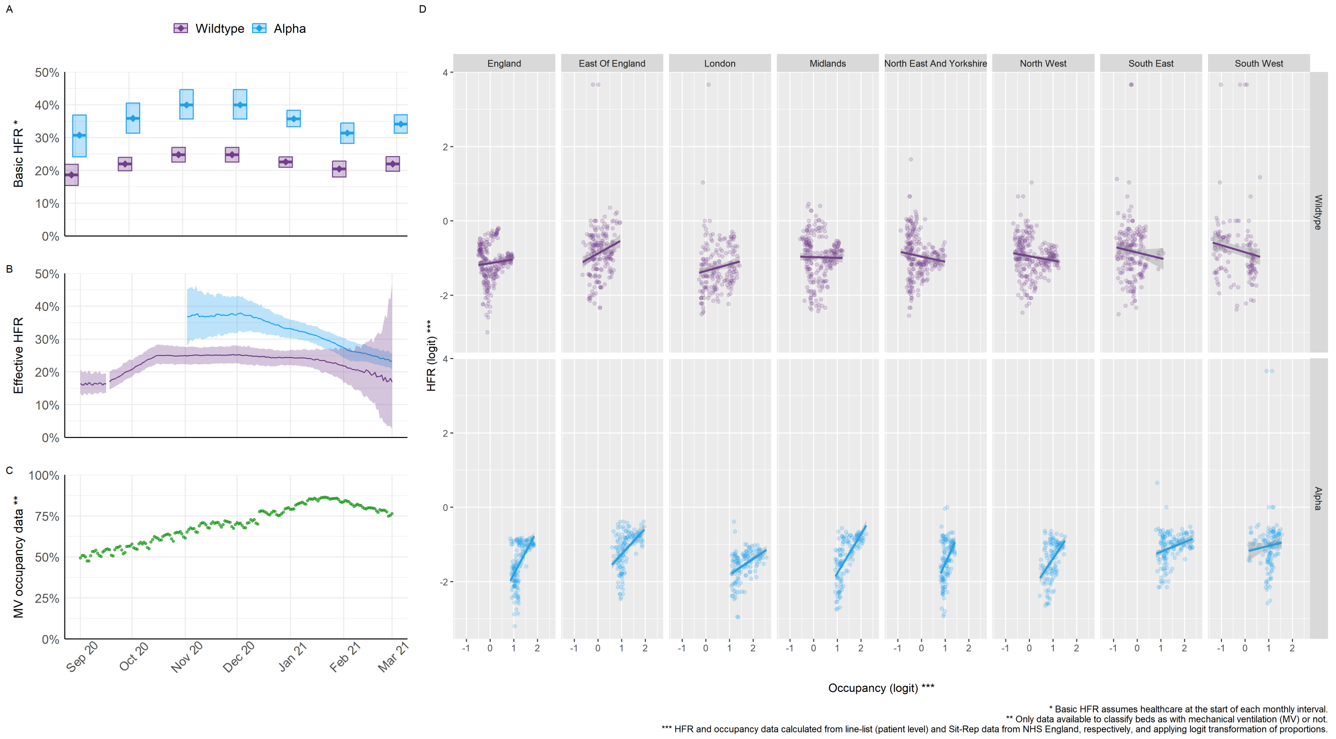

**Figure S11:** Correlation between hospital fatality ratio and mechanical ventilation beds occupancy. A) Basic HFR (model inferred) of the Alpha and Wildtype variants assuming healthcare characteristics at the start of each month. B) Daily (model inferred) effective severity by variant. C) Data on mechanical ventilation beds occupancy. D) Correlation plot between occupancy (logit scale) and HFR (logit scale) by NHS England region and variant dominance.

| | Estimate | S.E. | t value | p |
|---|---|---|---|---|
| **Intercept** | -1.14 | 0.03 | -42.30 | $<$0.01 |
| Occupancy (logit) | 0.10 | 0.06 | 1.59 | 0.11 |
| Alpha dominant | -1.84 | 0.16 | -11.45 | $<$0.01 |
| Occupancy (logit) * Alpha dominant | 1.09 | 0.14 | 7.78 | $<$0.01 |

**Table S22:** Correlation between hospital fatality ratio and mechanical ventilation beds occupancy. Ordinary least square linear regression model, with dependent and independent variables in logit scale, and variant dominance included as an interaction term. Adjusted $R^2 = 0.18$.

#### 6.3 Severity outputs by vaccination status

We compared modelled *vs* empiric hospital admissions and deaths by vaccine status over time, aggregated from national patient-level linked records of vaccination status, hospitalisations and deaths. The model performed well in capturing these trends, which we did not explicitly fit to, indicating it was able to correctly infer the effect of vaccine-derived immunity against hospitalisation and death at the population level.

It should be further noted that our model includes for time-varying patterns of severity mechanisms (section 4.4.4), as proxy of healthcare performance variations over time. Results below thus support there was an overall low risk of the model wrongly attributing differences in variant basic severity to factors driven by the evolving profiles of vaccine-derived immunity or healthcare performance.

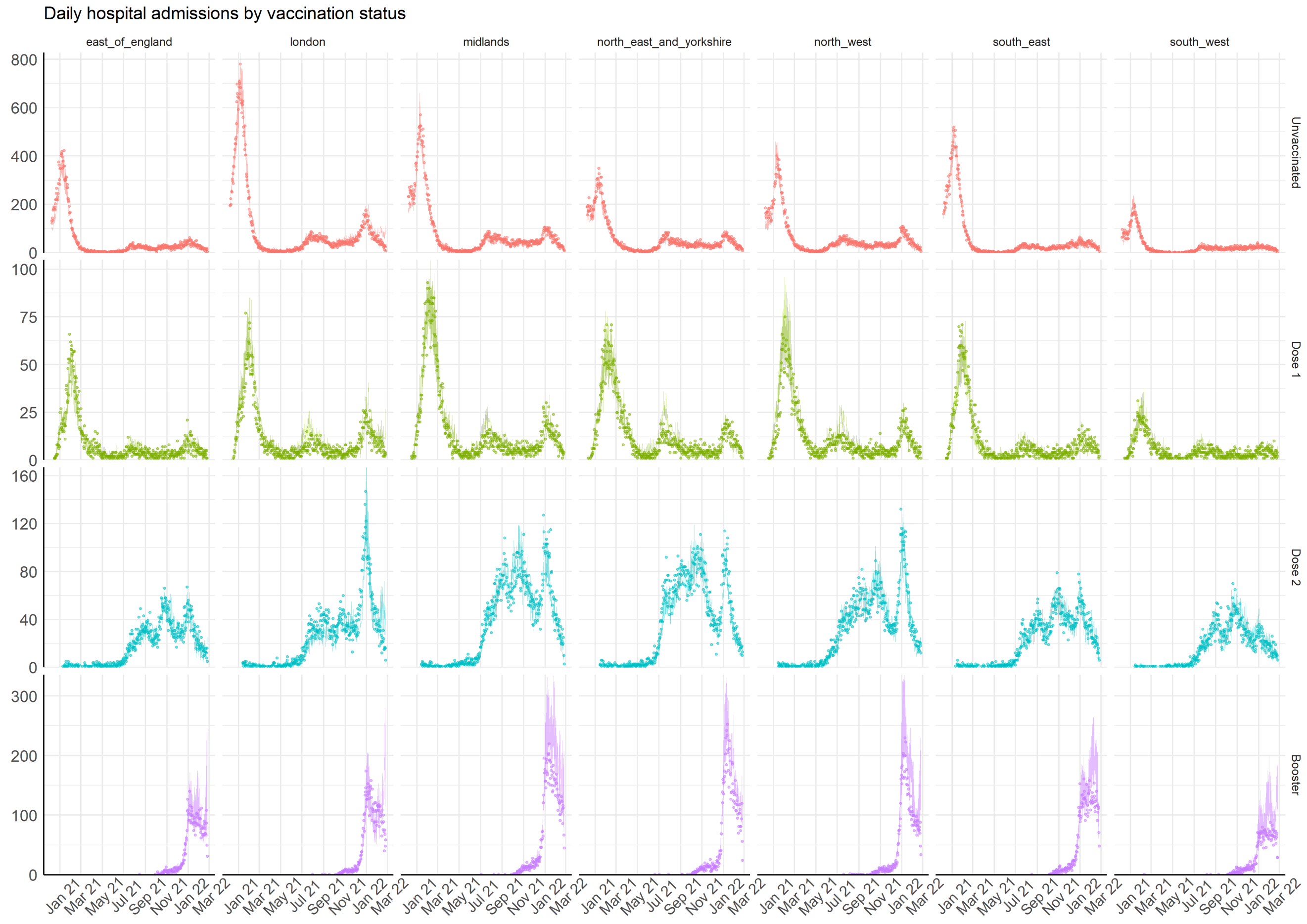

**Figure S12:** Points represent data on daily admissions by vaccination status (see section 6.2) for details of data aggregation from linked patient-level line lists) and shaded areas 95%CrI of model inferred trajectories by NHS England region.

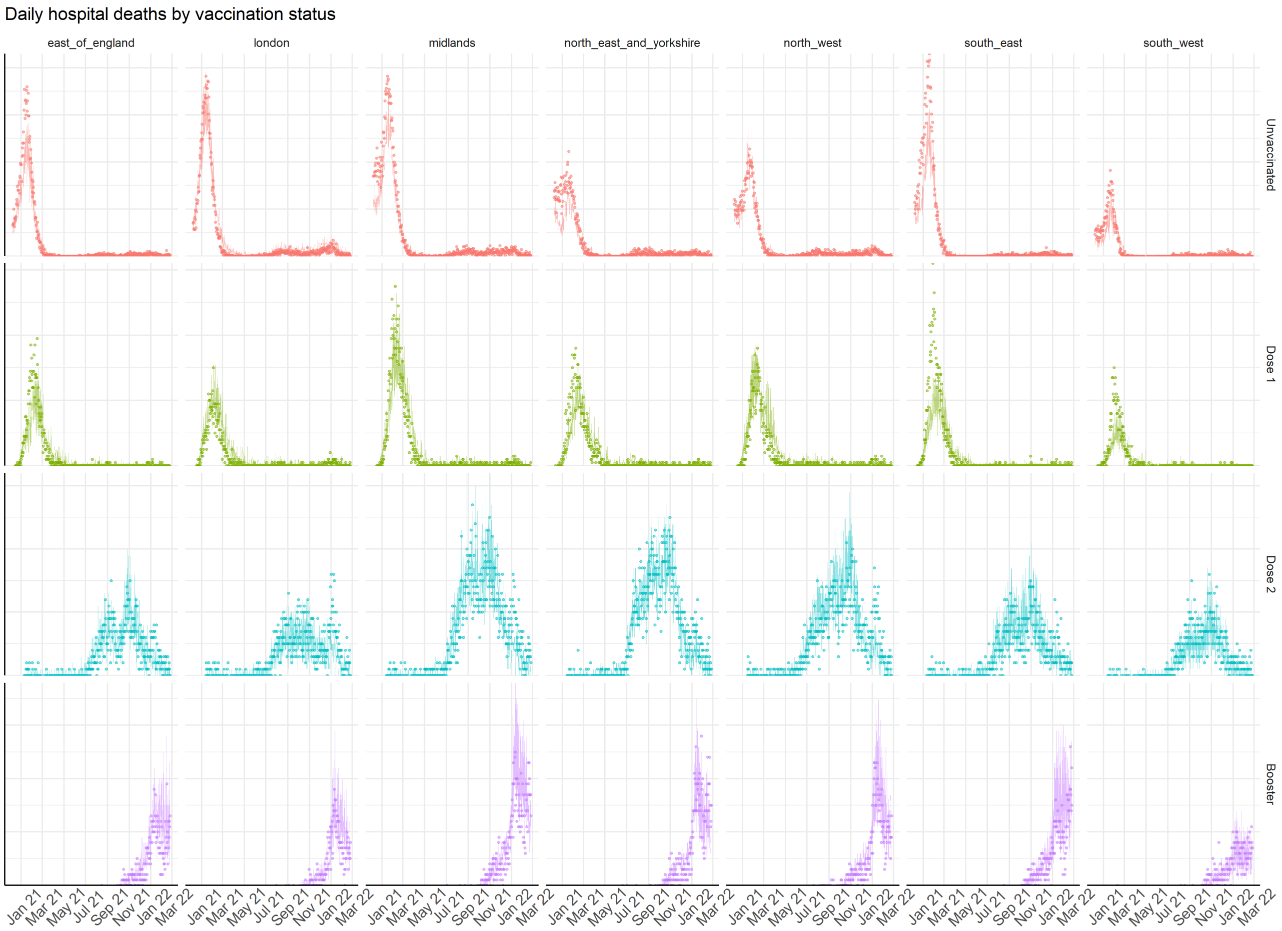

**Figure S13:** Points represent data on daily hospital deaths by vaccination status (see section 6.2) for details of data aggregation from linked patient-level line lists) and shaded areas 95%CrI of model inferred trajectories by NHS England region.

#### 6.4 Model comparison to other empiric severity outputs over time

There are additional nuances around variations in patient cohorting practices within the healthcare system over time we could not account for in our model. These include changes in the clinical threshold for admitting patients or for triaging them to critical care. This could have implied that the model could have still suffered from parameter identifiability issues when attributing severity to basic properties of the variants *vs* the above factors.

To reduce this risk, we conducted additional checks to ensure the model was able to reproduce the overall HFR by England region, and the age distribution of hospitalisations and deaths over time. Lastly, we also present inferred IHR, HFR and IFR by age over time and the relation in uncertainty (and overlaps) between the intrinsic $R_0$ and the basic severity properties of the variants by NHS England region.

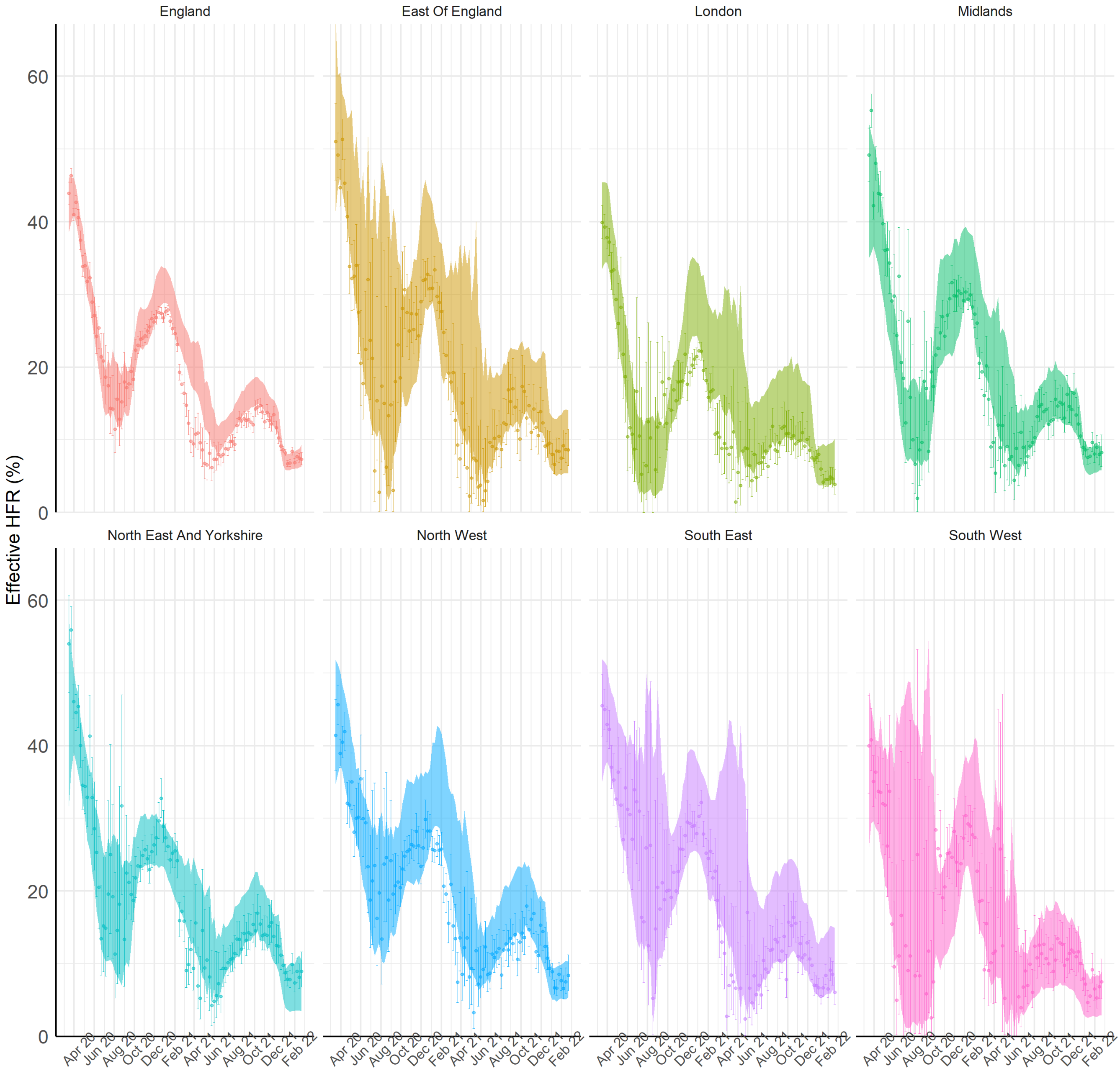

**Figure S14:** Point-range represents data and binomial confidence interval (see section 6.2) for details of data aggregation from linked patient-level line lists) and shaded areas 95%CrI of model inferred trajectories by NHS England region.

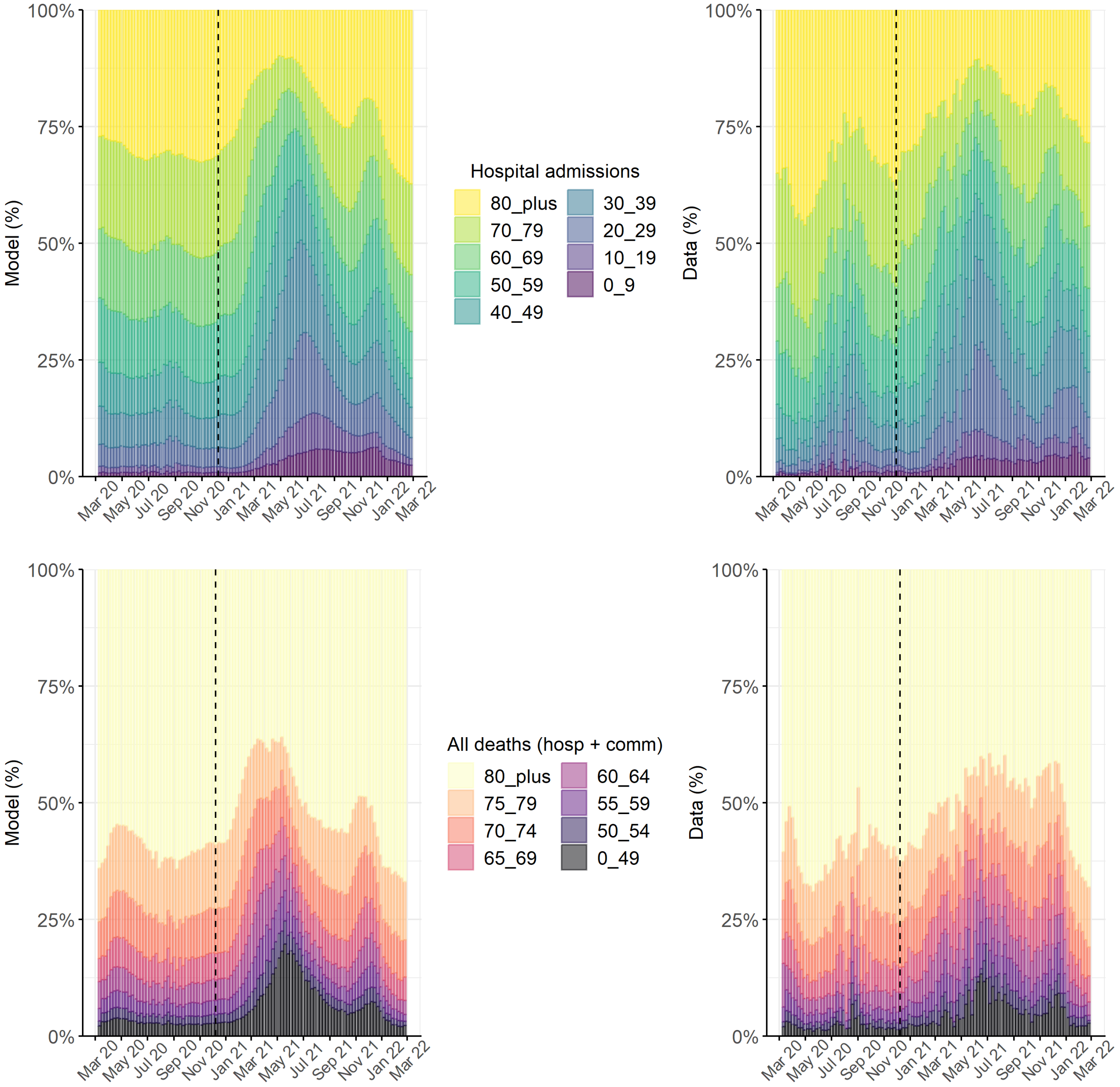

**Figure S15:** Comparison of model (left column) and data (right column) age distribution of weekly hospital admissions (top row) and deaths (bottom row, in hospital and in the community).

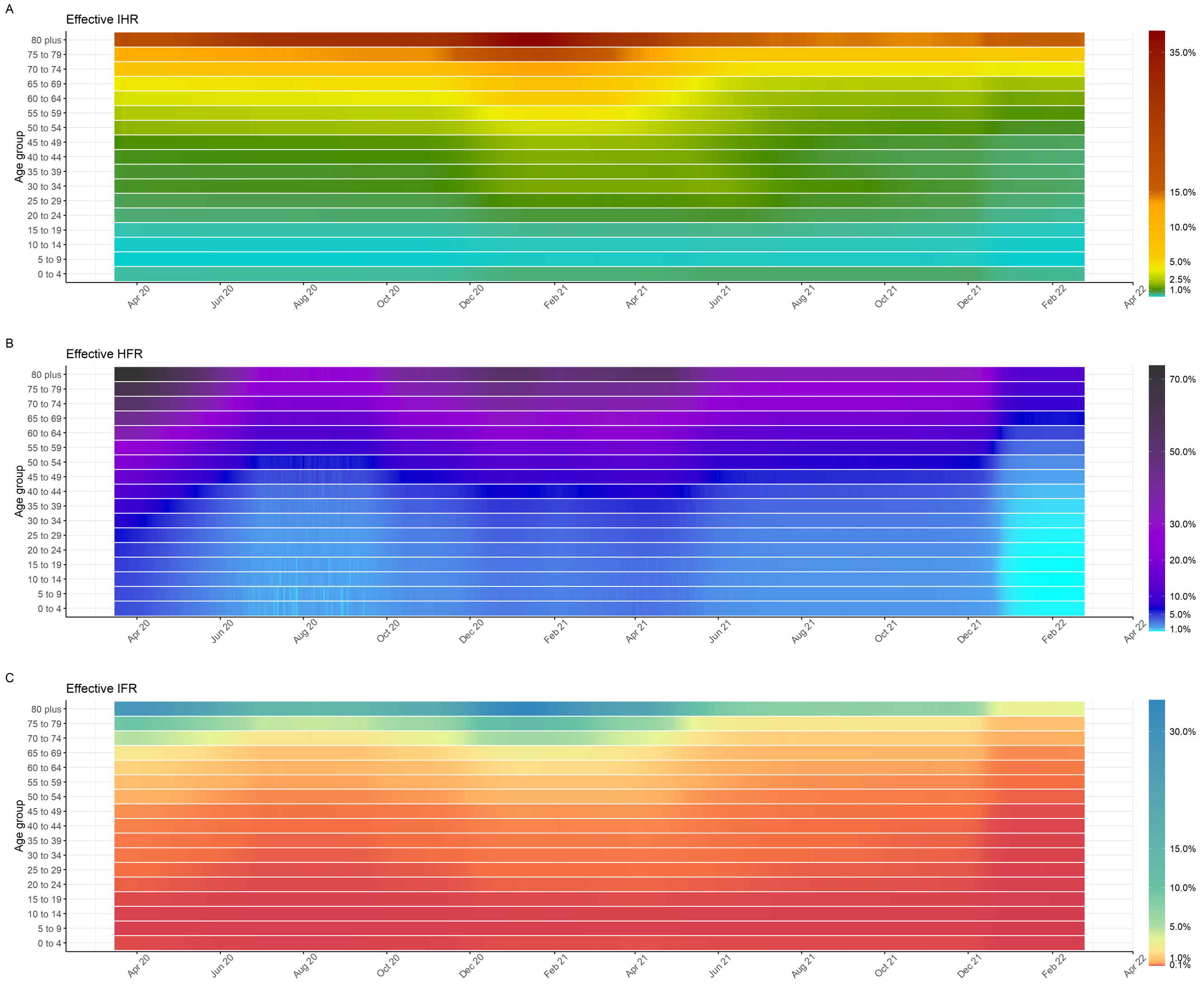

**Figure S16:** Effective infection hospitalisation ratio (IHR), hospital fatality ration (HFR) and infection fatality ration (IFR) by model age compartments over time, accounting for immunity by age class. Please note different colour gradient scales for each heatmap.

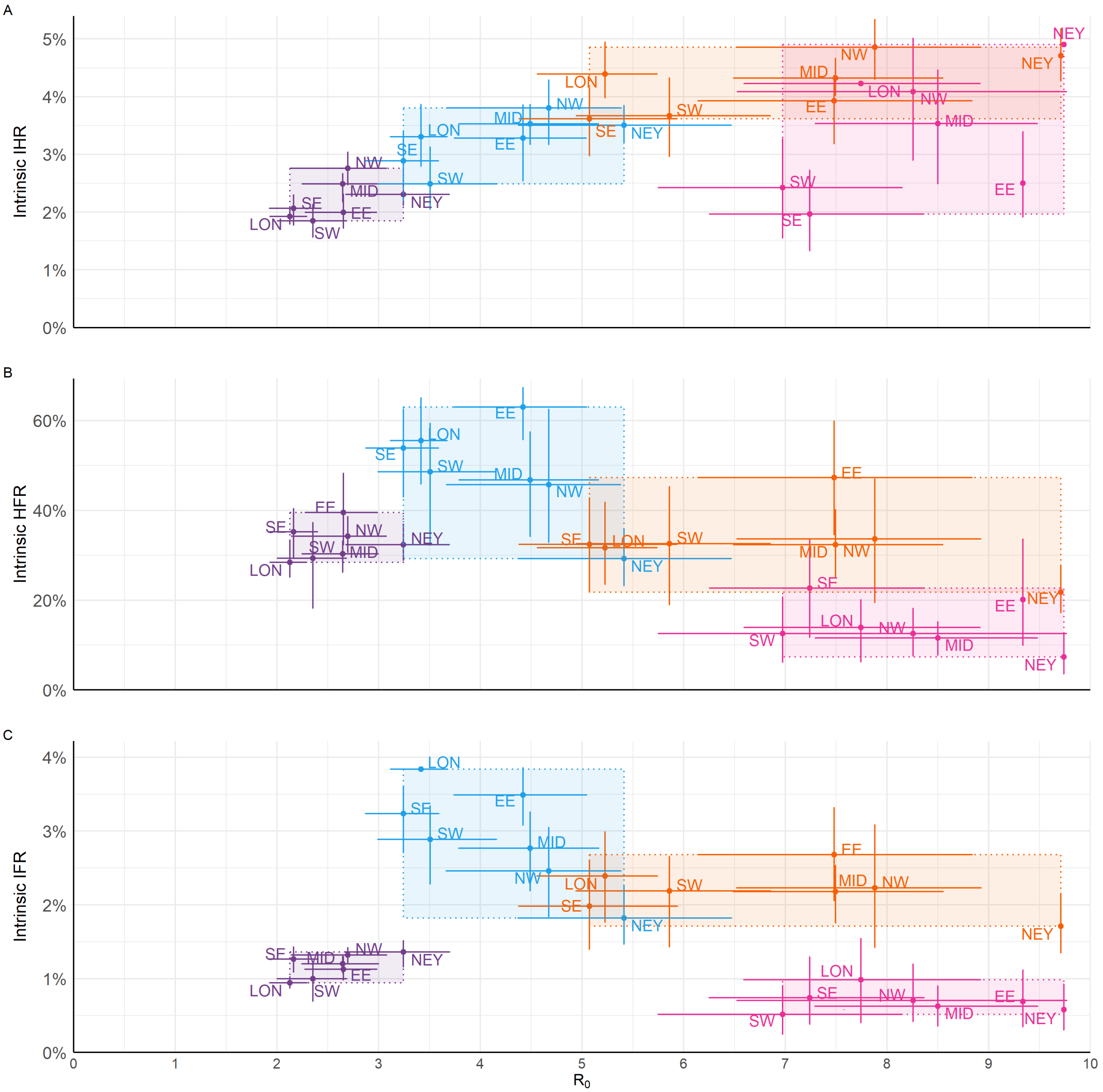

**Figure S17:** Scatter plot of intrinsic $R_0$ and basic infection hospitalisation ratio (IHR), hospital fatality ration (HFR) and infection fatality ration (IFR) by variant and NHS England region. EE, East of England; LON, London; MID, Midlands; NEY, North East and Yorkshire; NW, North West; SE, South East; and SW, South West.

#### 6.5 Number of particles check

To check on the robustness of our results given selection of number of particles for the pMCMC, we compare inferred values of the intrinsic $R_0$ and basic severity with central parameters using 192 (default) and 384 (double) pMCMC particles. We ran these from the same initial (pre-tuned) parameter values and VCV proposal matrix (see section 4.11.

| | Intrinsic $R_0$ | Basic $IHR$ | Basic $HFR$ | Basic $IFR$ |
|---|---|---|---|---|
| **192 particles (default)** | | | | |
| Wildtype | 2.6 (2.4-2.7) | 2.2 (2.1-2.3) | 32.6 (30.7-34.4) | 1.2 (1.1-1.2) |
| Alpha | 4.2 (3.9-4.4) | 3.3 (3.0-3.5) | 48.5 (44.3-52.9) | 2.9 (2.7-3.2) |
| Delta | 7.0 (6.5-7.4) | 4.2 (4.1-4.4) | 32.6 (29.2-36.6) | 2.2 (2.0-2.4) |
| Omicron | 8.3 (7.7-8.8) | 3.4 (3.1-3.8) | 14.3 (11.3-17.1) | 0.7 (0.6-0.8) |
| **384 particles** | | | | |
| Wildtype | 2.5 (2.3-2.6) | 2.3 (2.2-2.4) | 31.0 (29.4-32.4) | 1.1 (1.0-1.2) |
| Alpha | 4.0 (3.8-4.2) | 3.5 (3.4-3.7) | 46.4 (43.2-49.5) | 2.7 (2.5-2.9) |
| Delta | 6.8 (6.5-7.2) | 4.4 (4.2-4.5) | 29.0 (26.0-31.2) | 1.9 (1.7-2.1) |
| Omicron | 8.1 (7.8-8.5) | 3.5 (3.2-3.7) | 12.9 (9.9-15.4) | 0.6 (0.5-0.7) |

**Table S23:** Main model results (see in main manuscript) with default of 192 pMCMC particles *vs* 384.

### 6.6 Model fit to healthcare data

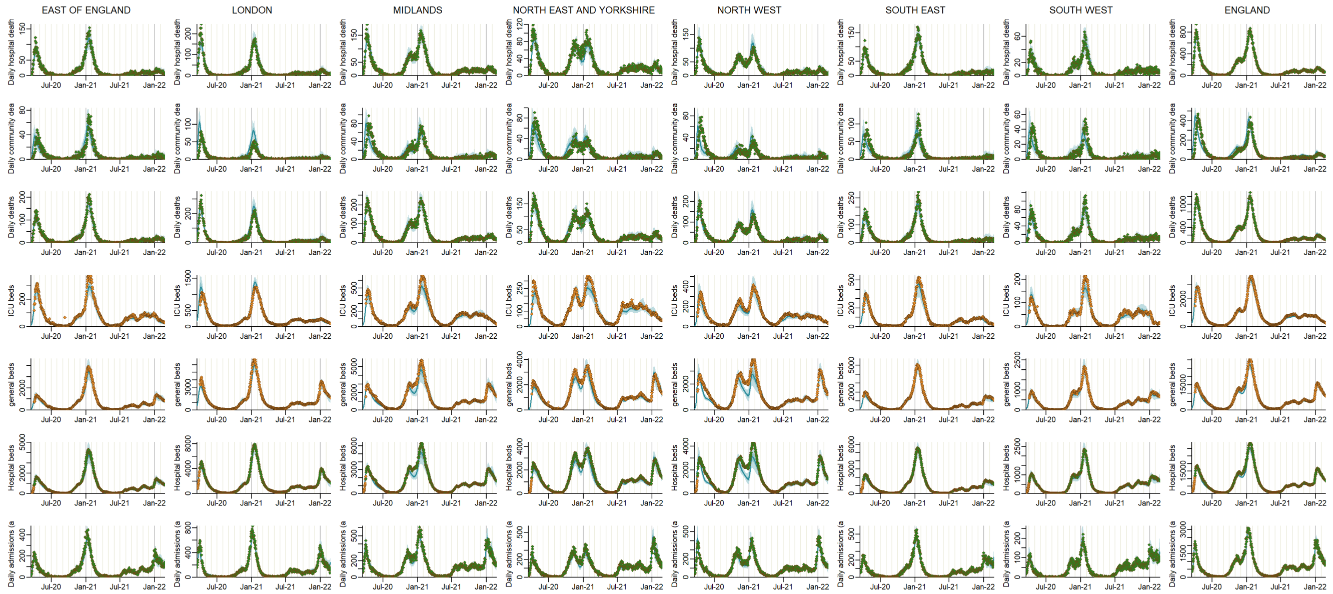

**Figure S18:** Model fits to NHS England Regions (columns); from top row to bottom: Deaths in hospital, Deaths in the community, all deaths, ICU beds occupancy, general beds occupancy and all daily admissions. Points show the data, solid line the median model fit and the shaded area the 95% CrI. Green data points indicate data streams where the model was fitted to age-disaggregated data (see section 6.7), and orange where it was fitted to aggregated data as shown.

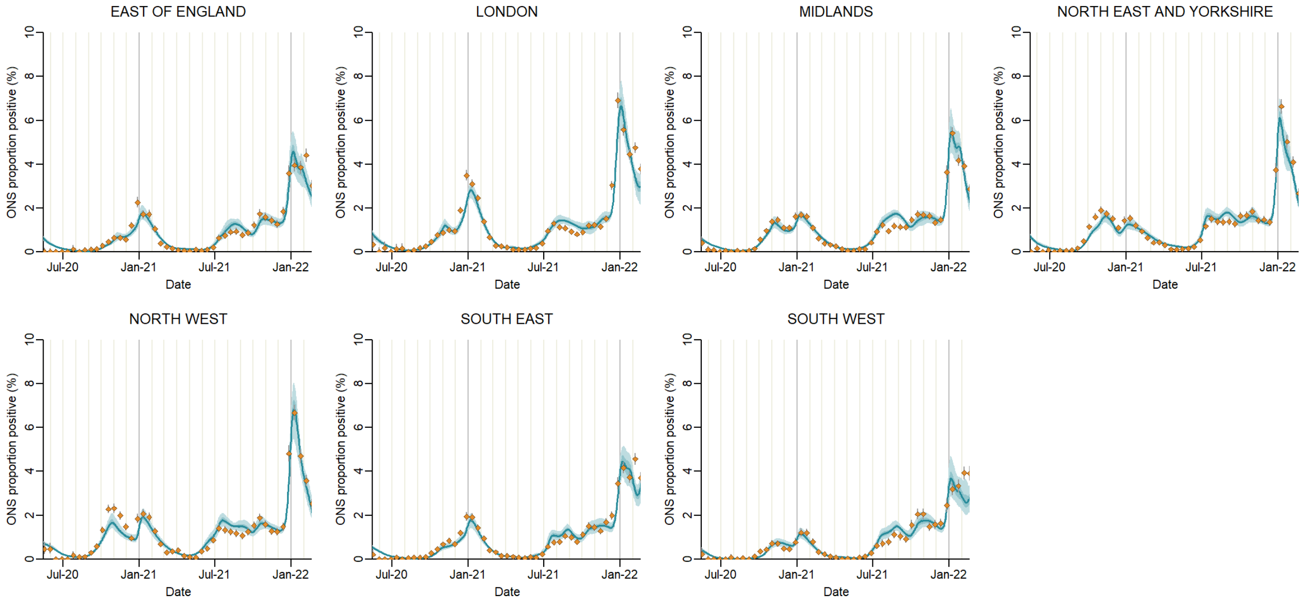

**Figure S19:** Model fits to NHS England Regions (columns): infection prevalence in the community from ONS infection survey. Points show the data, the solid line the median model fit and the shaded area the 95% CrI.

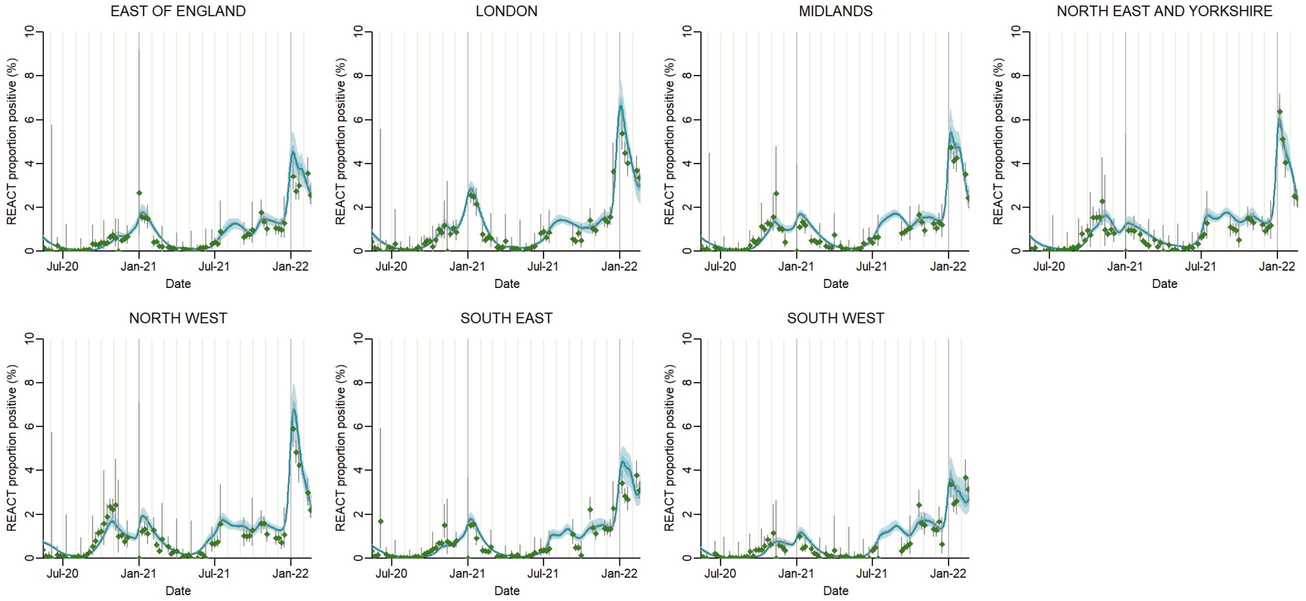

**Figure S20:** Model fits to NHS England Regions (columns): infection prevalence in the community from the REal-time Assessment of Community Transmission (REACT) Study. Points show the data, the solid line the median model fit and the shaded area the 95% CrI. Geen colour points indicate the model was fitted to age-disaggregated data (see section 6.7).

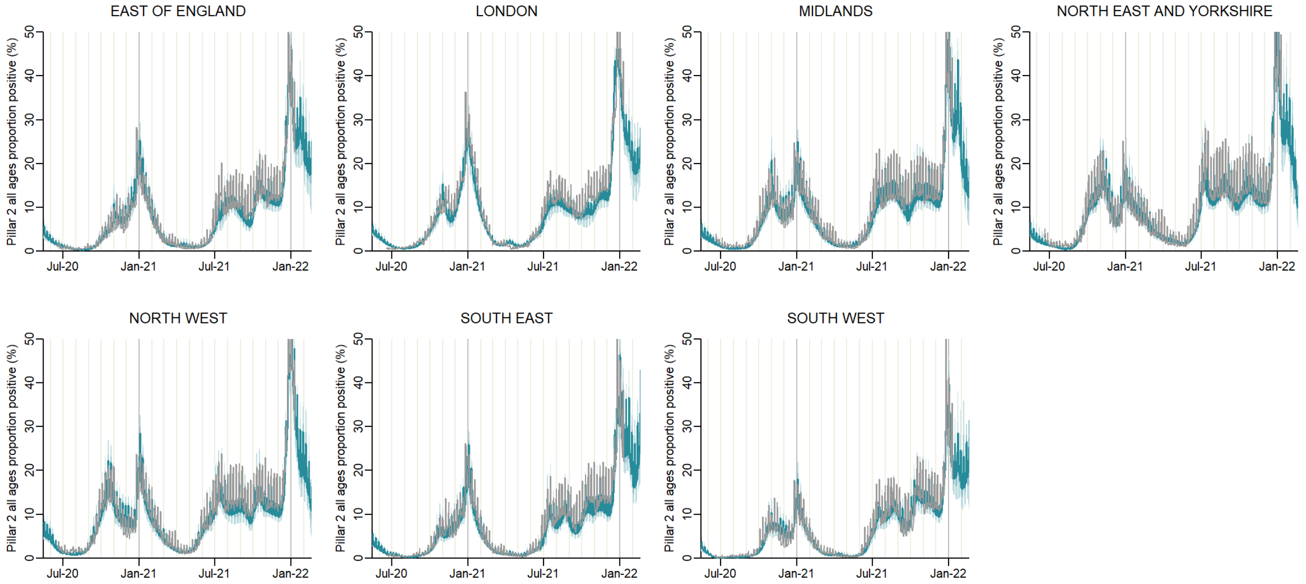

**Figure S21:** Model fits to NHS England Regions (columns): PCR positivity in the community from the national community testing programme (Pillar 2). Grey line shows the data, aggregated for all ages, and the blue line the median model fit and the shaded area the 95% CrI. Grey colour indicates the model was fitted to age-disaggregated data (see section 6.7). PCR positivity data was removed before 2020-06-18, as the programme had not been scaled up nationally before this date, and after 2021-11-01.

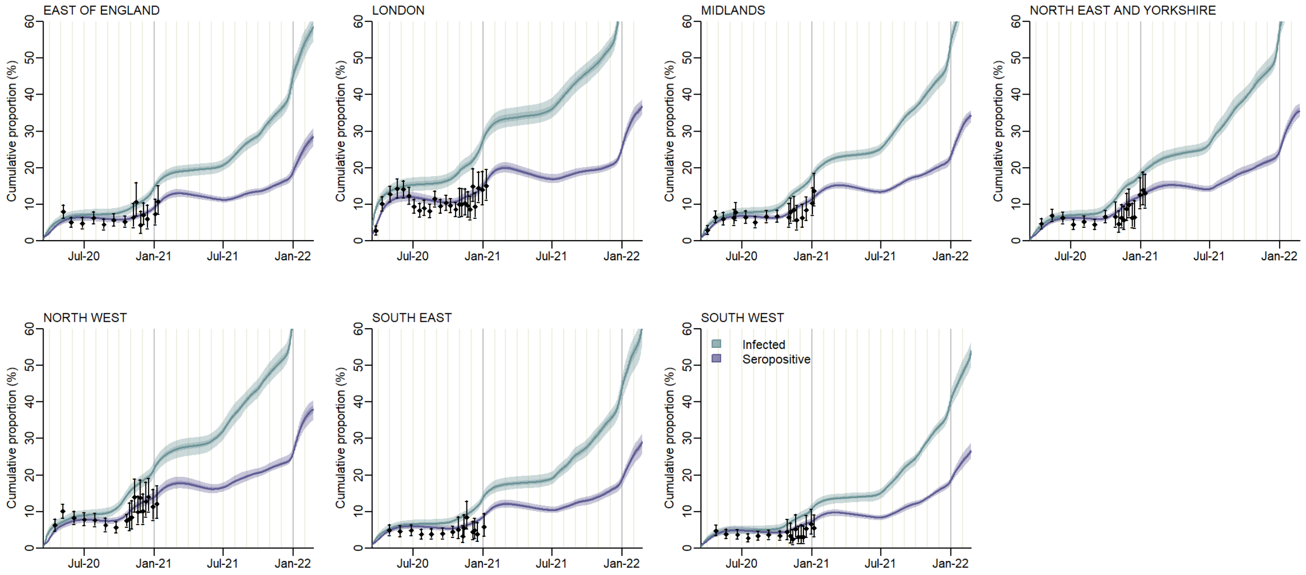

**Figure S22:** Model fits to NHS England Regions (columns): seropositivity by Euroimmun essay among blood donors. Black point-range shows the data and binomial confidence interval, and the blue and purple lines the median model fit to proportion seropositive and infected, respectively, and shaded area the 95% CrI.

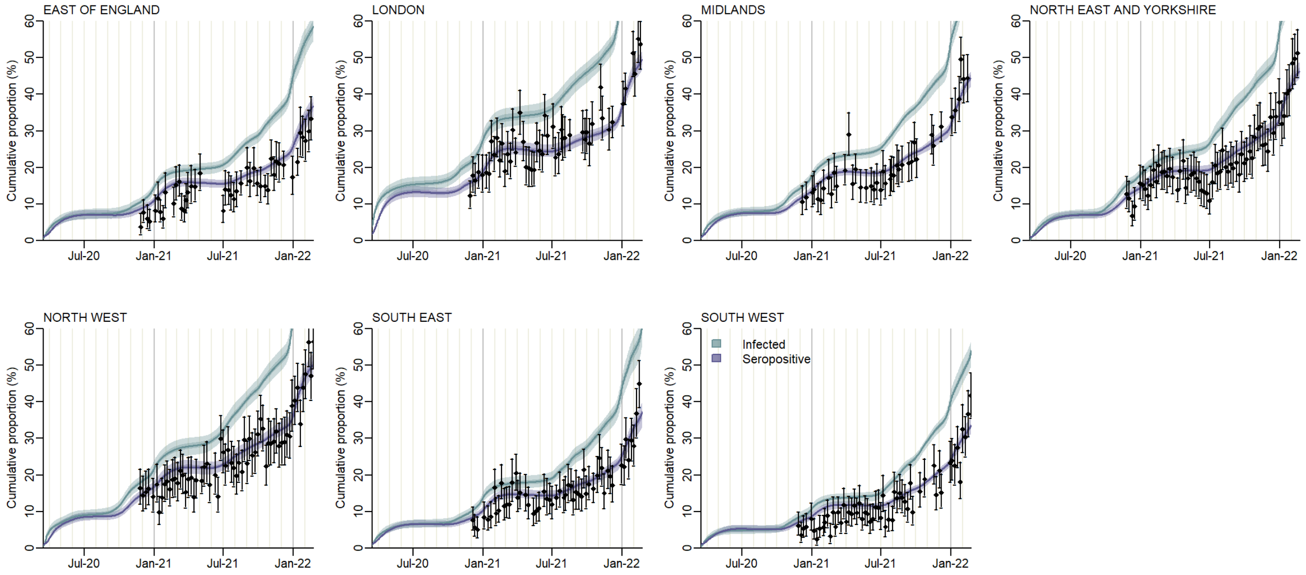

**Figure S23:** Model fits to NHS England Regions (columns): seropositivity by Roche N essay among blood donors. Black point-range shows the data and binomial confidence interval, and the blue and purple lines the median model fit to proportion seropositive and infected, respectively, and shaded area the 95% CrI.

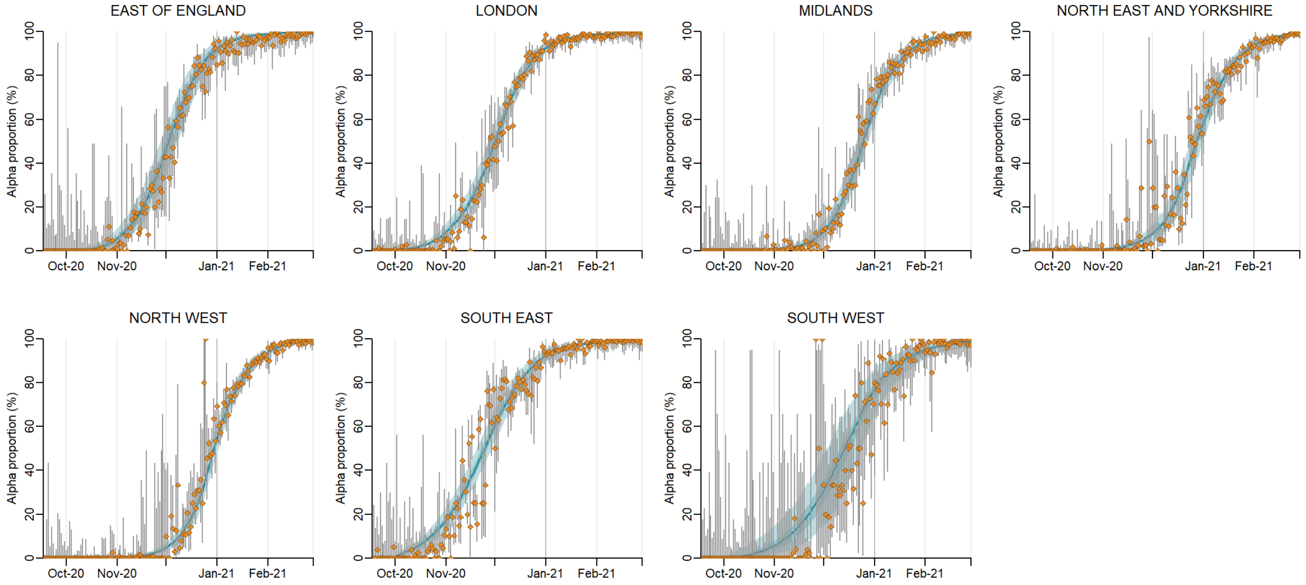

**Figure S24:** Model fits to NHS England Regions (columns): Alpha variant frequency among symptomatic Pillar 2 PCR-positive cases. Points-range shows the data and binomial confidence interval, solid line the median model fit and the shaded area the 95% CrI.

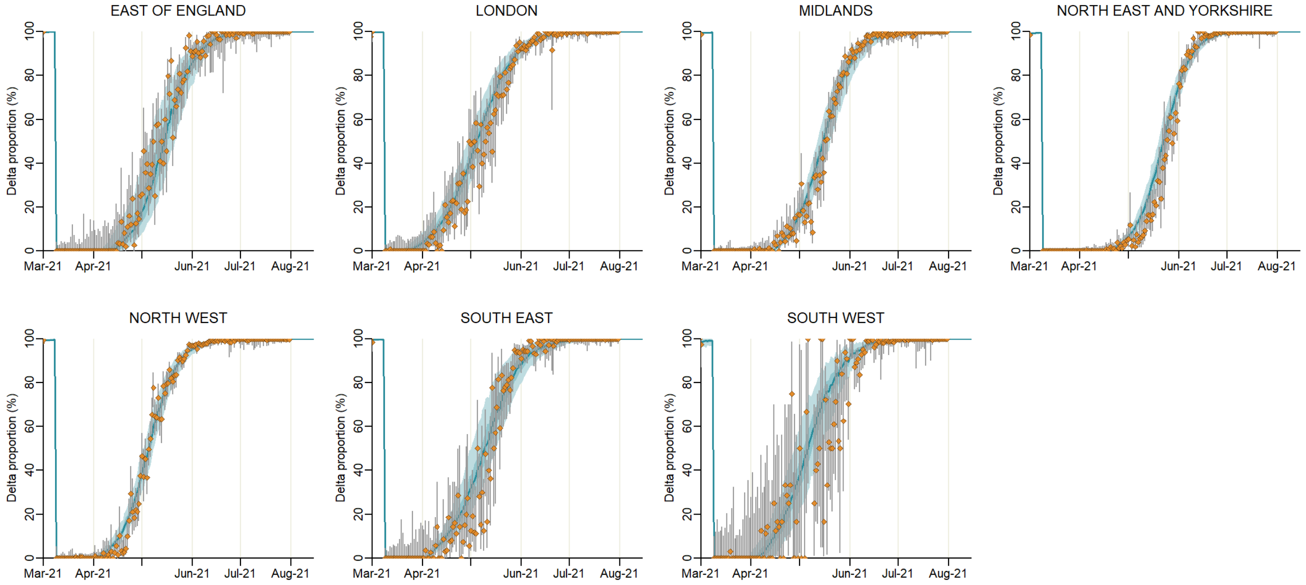

**Figure S25:** Model fits to NHS England Regions (columns): Delta variant frequency among symptomatic Pillar 2 PCR-positive cases. Points-range shows the data and binomial confidence interval, solid line the median model fit and the shaded area the 95% CrI.

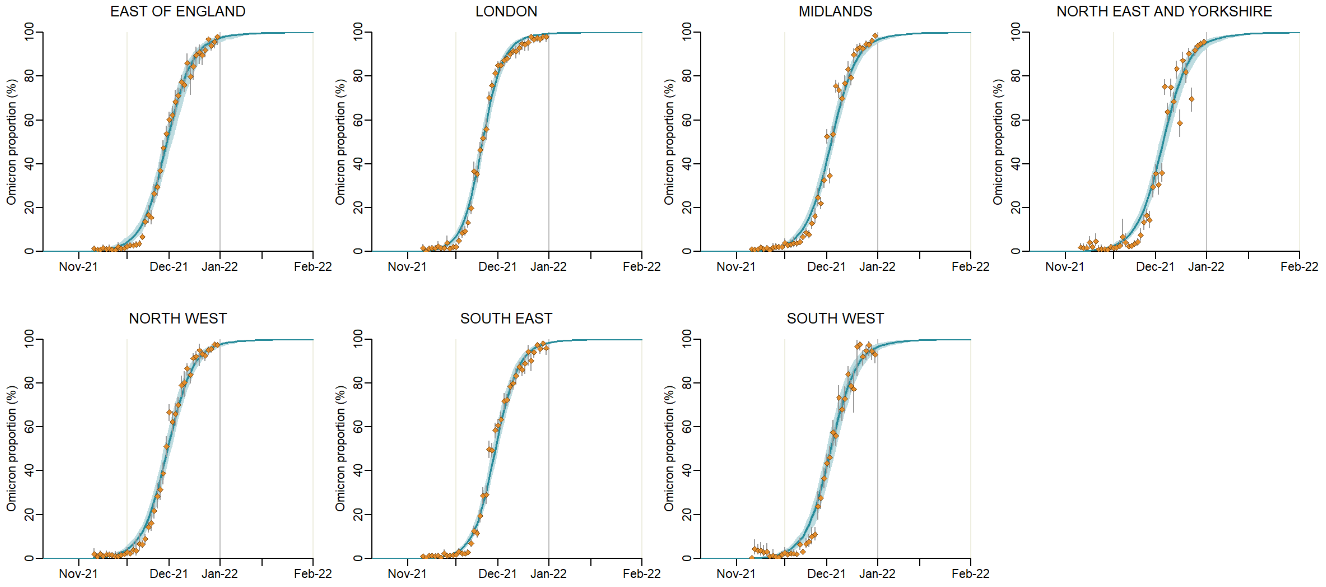

**Figure S26:** Model fits to NHS England Regions (columns): Omicron variant frequency among symptomatic Pillar 2 PCR-positive cases. Points-range shows the data and binomial confidence interval, solid line the median model fit and the shaded area the 95% CrI.

#### 6.7 Model fit to age-disaggregated data

The model was fitted to age-specific data by NHS England region for Pillar 2 PCR positivity, infection prevalence from the REACT study, hospital admissions and deaths, both in hospital and in the community. Age bands were defined according to data availability for each data stream. Here we present the model fits compared to these age-disaggregated data streams, aggregating across regions to the national (England) level.

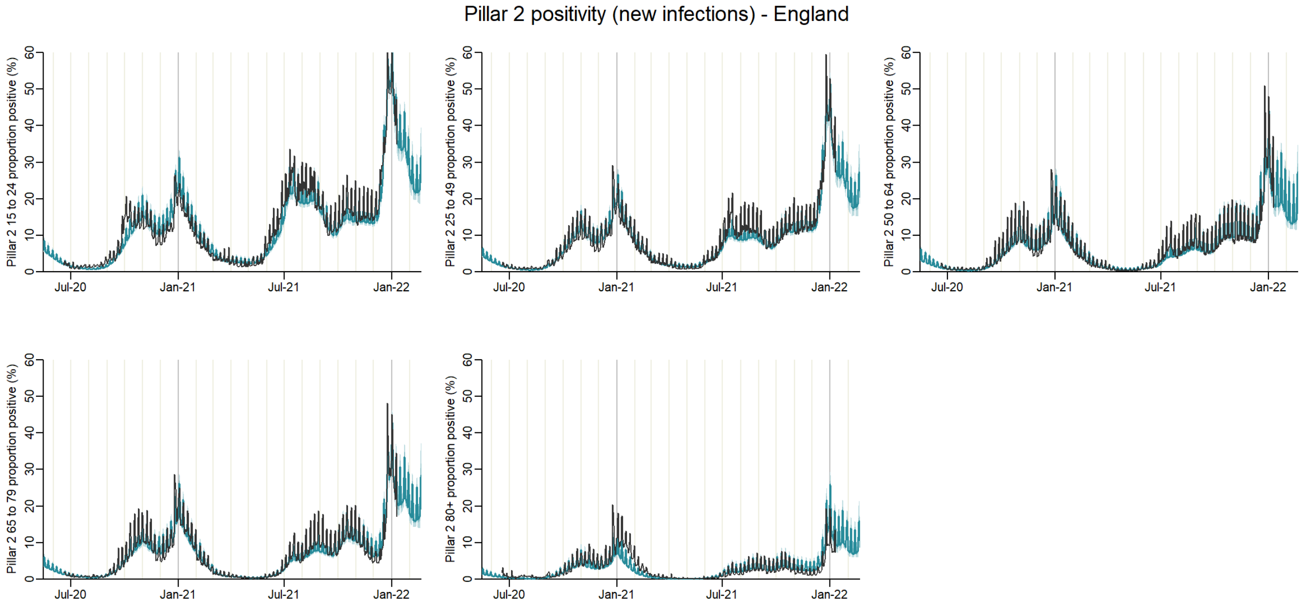

**Figure S27:** Model fits to data by age aggregated to national-level: PCR positivity in the community from the national community testing programme (Pillar 2). Black line shows the data, and the blue line the median model fit and the shaded area the 95% CrI.

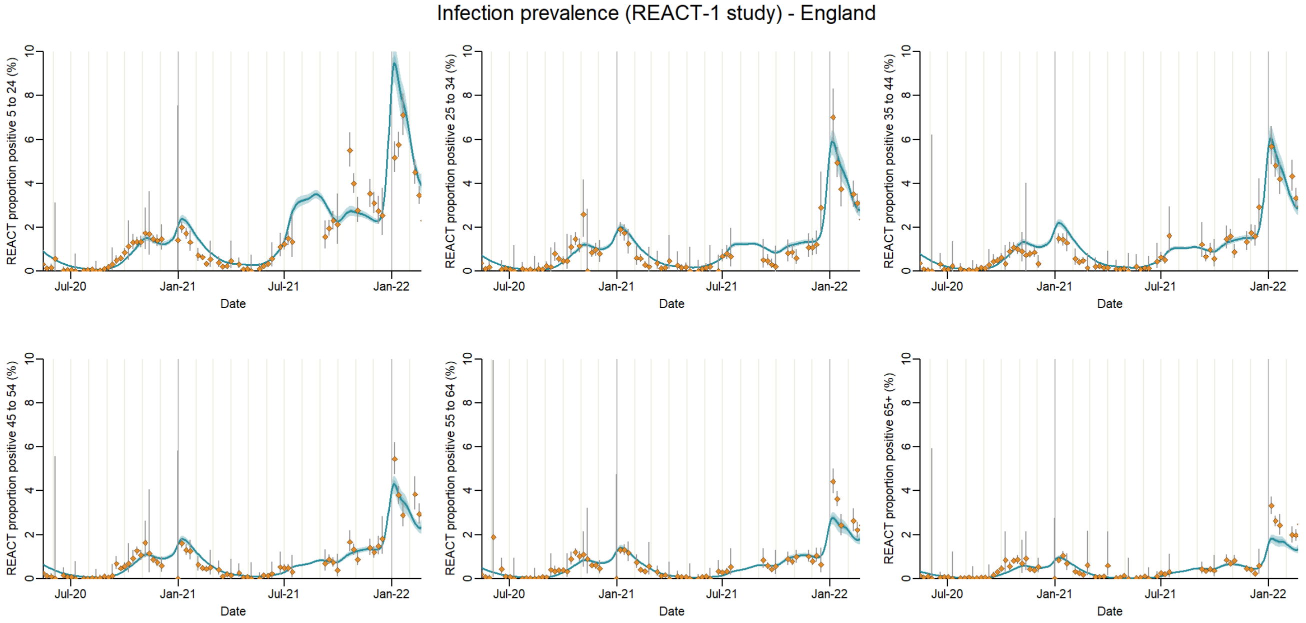

**Figure S28:** Model fits to age-disaggregated data: infection prevalence in the community from the REal-time Assessment of Community Transmission (REACT) Study. Grey line shows the data, aggregated to England-level, and the blue line the median model fit and the shaded area the 95% CrI.

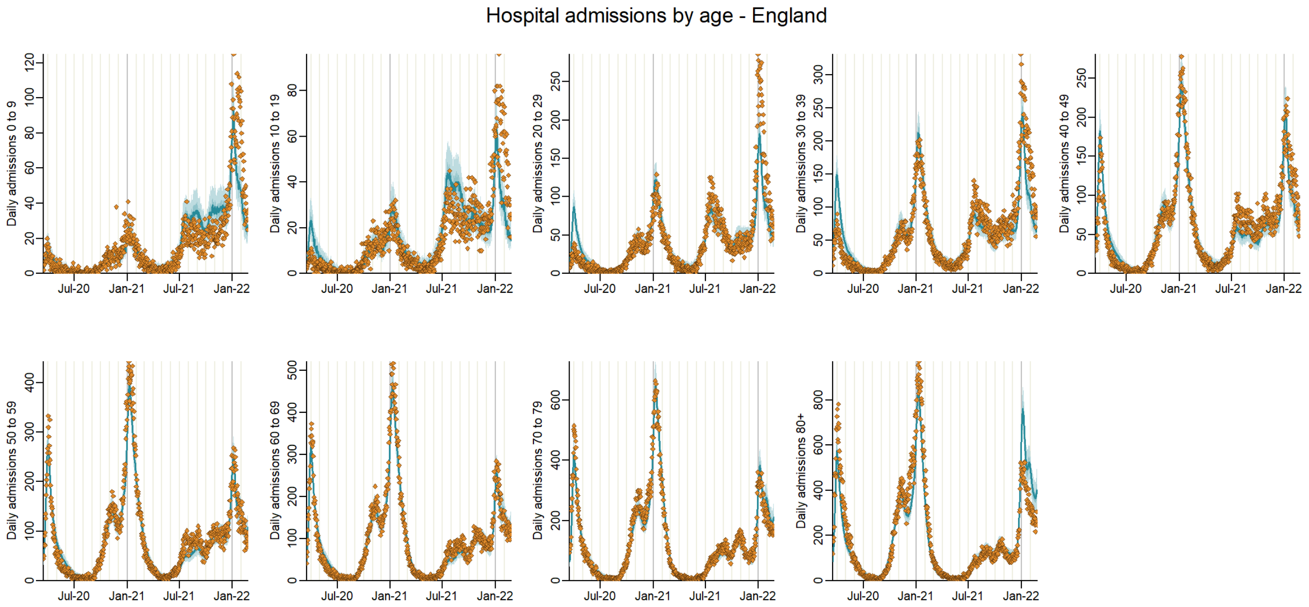

**Figure S29:** Model fits to age-disaggregated data: hospital admissions. Points-range shows the data and binomial confidence interval, aggregated to England-level, and the blue line the median model fit and the shaded area the 95% CrI.

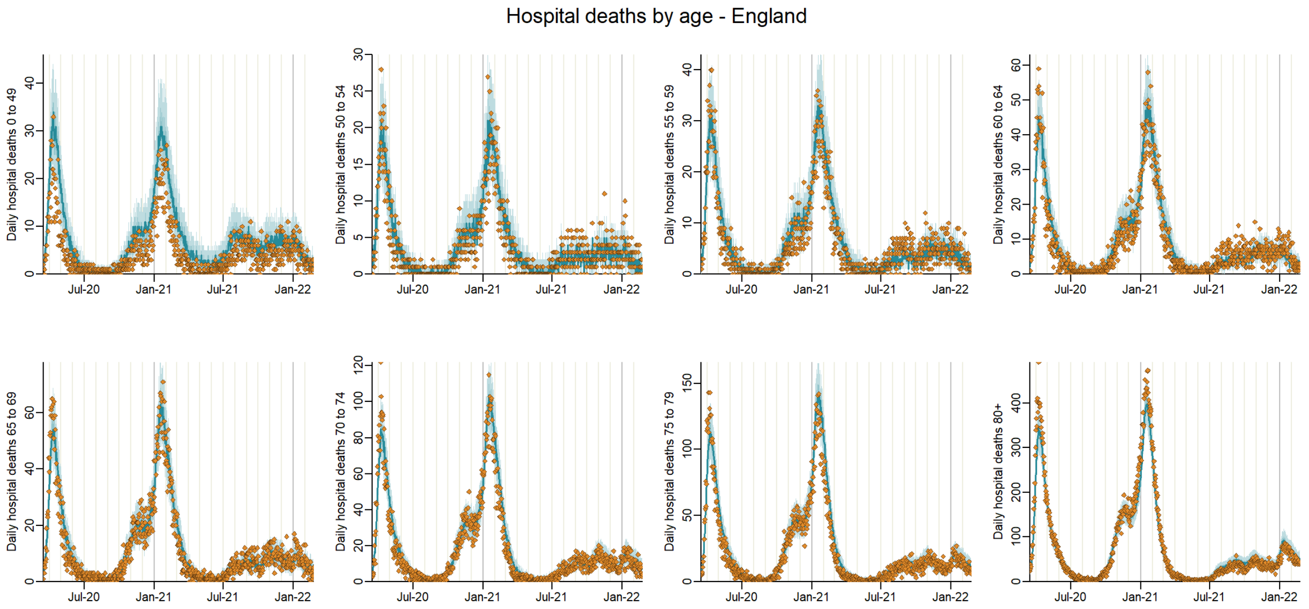

**Figure S30:** Model fits to age-disaggregated data: hospital deaths. Orange points show the data, aggregated to England-level, and the blue line the median model fit and the shaded area the 95% CrI.

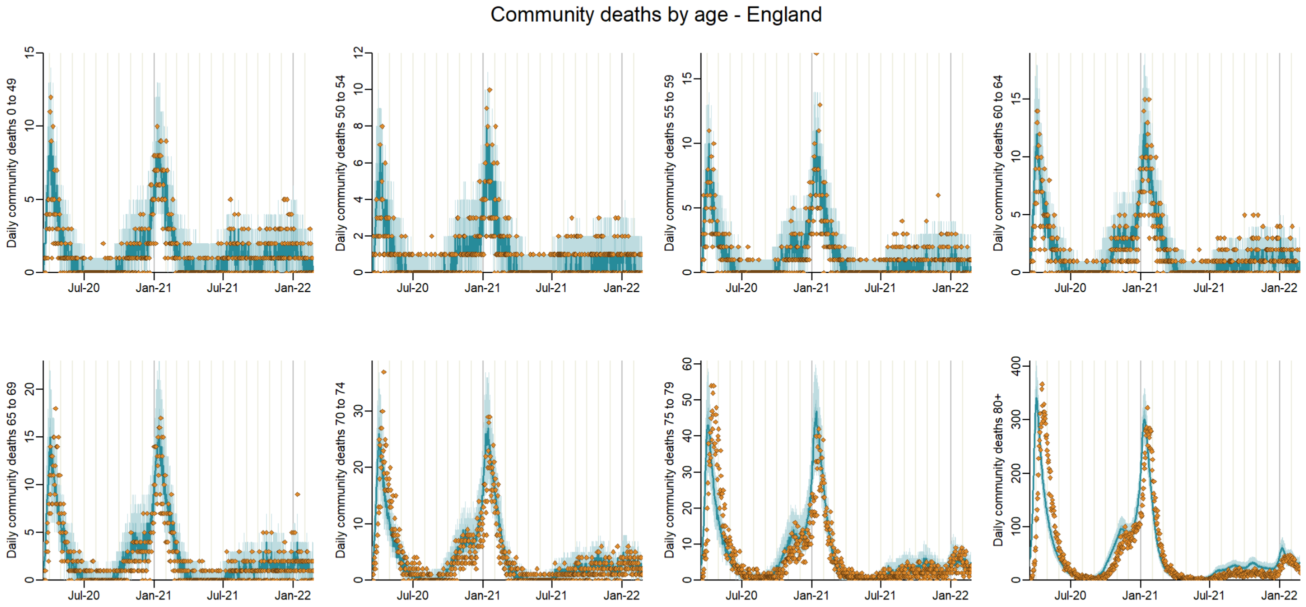

**Figure S31:** Model fits to age-disaggregated data: deaths in the community, including care homes. Orange points show the data, aggregated to England-level, and the blue line the median model fit and the shaded area the 95% CrI.

### 6.8 Model inferred (fitted) parameters

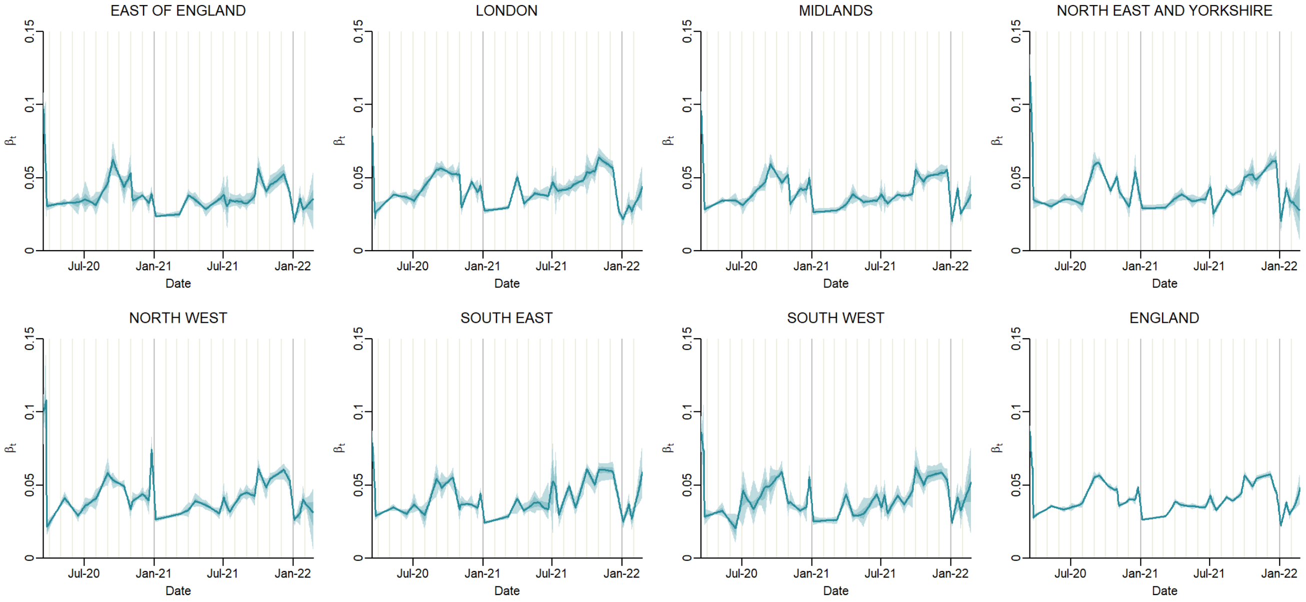

**Figure S32:** Estimated trajectory of beta over time by region. Solid lines show the median model trajectory and the shaded areas the 95% CrI.

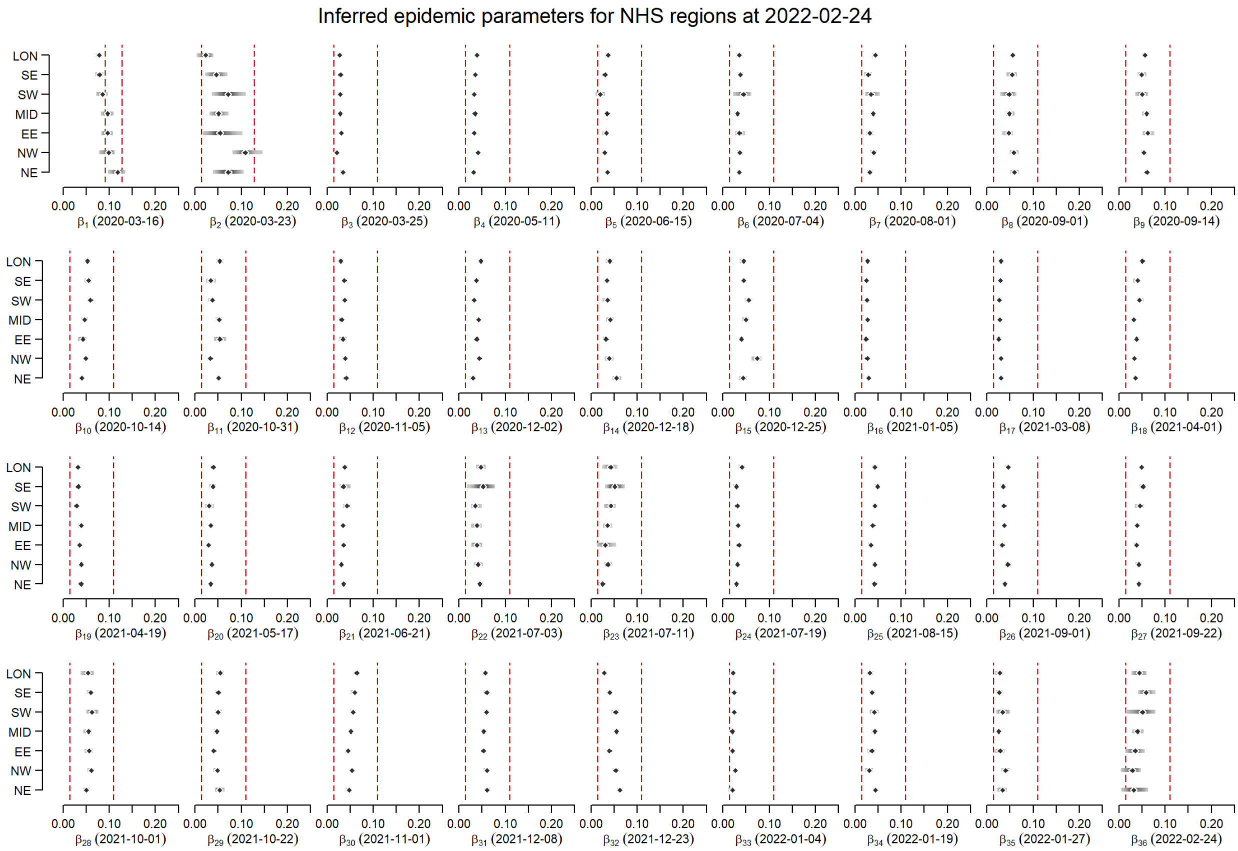

**Figure S33:** Estimated value of beta over time by region. The points show the median model fit and the bars the 95% CrI. The vertical dashed red lines show the bounds of the prior distribution.

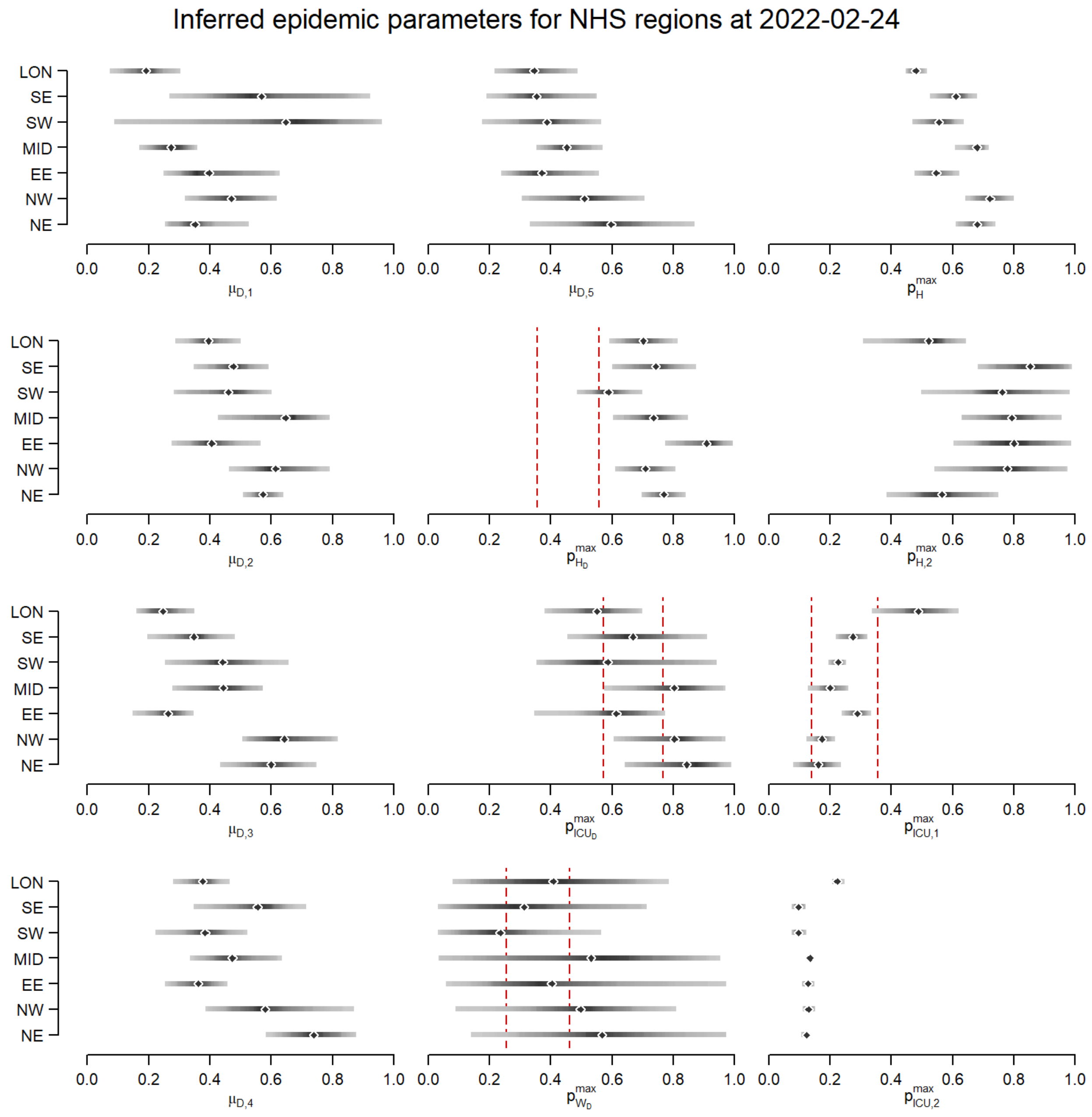

**Figure S34:** Model parameter posterior distributions for time-varying severity modifiers. The points show the median model fit and the bars the 95% CrI. Where shown, vertical dashed red lines show the bounds of the prior distribution, else we assumed a uniform (flat) prior.

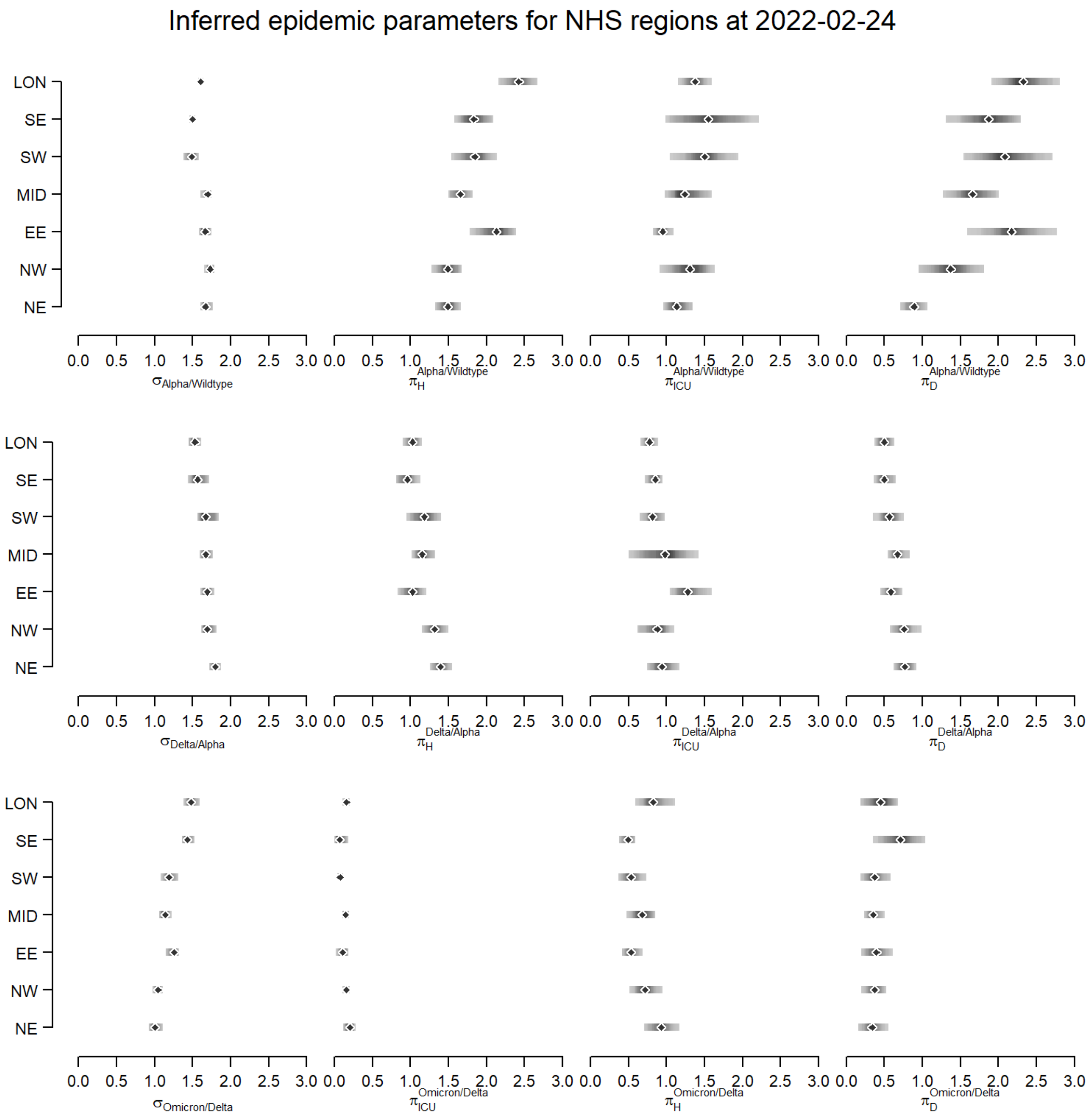

**Figure S35:** Model parameter posterior distributions for variant-driven severity modifiers. The points show the median model fit and the bars the 95% CrI. We assumed a uniform (flat) prior, hence no prior bounds are shown.

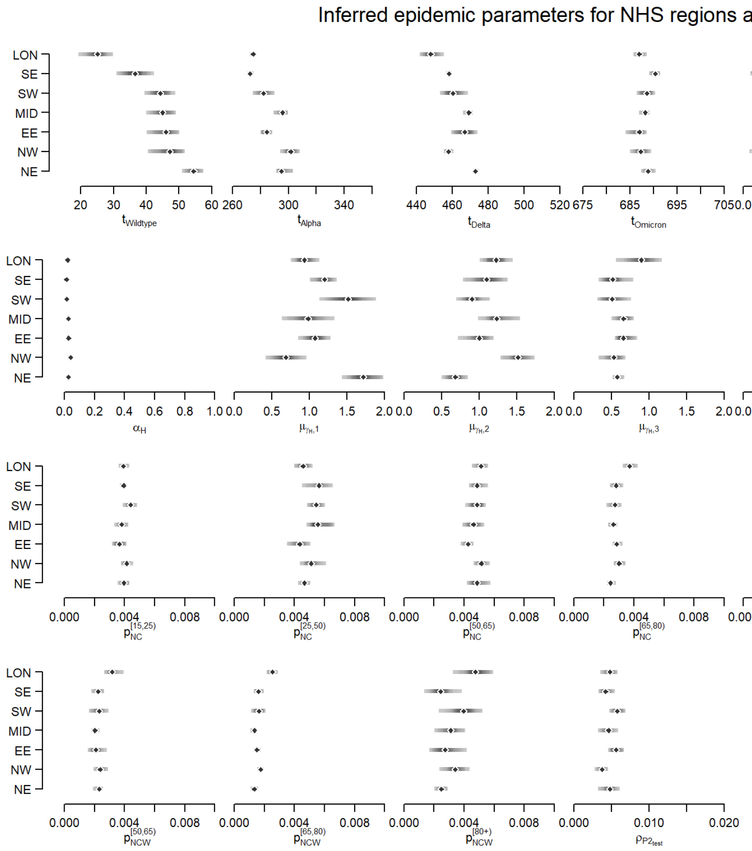

**Figure S36:** Model parameter posterior distributions. The points show the median model fit and the bars the 95% CrI. We assumed a uniform (flat) prior, hence no prior bounds are shown.

#### 6.9 Assessing convergence

As previously described (section 4.11), our framework followed a two-step fitting process whereby we first ran eight parameter pre-tunning MCMC chains ("expectation" model), followed by four pMCMC chains (stochastic). Note both the "expectation" and stochastic steps are the same model and share the same parameter space.

Each of the eight MCMC chains were started from different places of the 85-dimensional parameter space. To assess convergence of the parameter samples produced by the MCMC chains we performed:

- Visual inspection of the MCMC chains (Figures S37 to S43). The caterpillar-like appearance of the chains (8) for each parameters plus the shape of the log-likelihood curve suggests convergence and good coverage of the target area of the parameter space.
- Gelman-Rubin statistic (Table S24) associated with the full parameter chains (30,000 MCMC steps). All chains show $\hat{R}$ statistics below the recommended 1.1 threshold.
- Effective Sample Size (ESS) for the different parameters (Table S24) for the full parameter chains (30,000 MCMC steps).

Indeed, the above diagnostics suggest the MCMC chains reached the right area of the parameter space and that, therefore, the scales of the proposal distribution for the random walk were appropriate for the following pMCMCM algorithm. That is, MCMC chains reduced auto-correlation between successive samples. Subsequently, we used a re-scaled version of the VCV proposal matrix generated by the MCMC in the pMCMC algorithm. This was done in order to capture the main correlation structure and the "scale" of the different parameters, where diagonal elements of the VCV matrix represent the variances of the different parameters.

It should be noted that there are no existing diagnostics to guarantee MCMC convergence, but only necessary tests. Nevertheless, by relying on a group of diagnostics rather than a single one, we thoroughly tested the validity of our fitting process, as at least one of these diagnostics would have failed otherwise. Please see the posterior distributions of the fitted parameters after the pMCMC in figures S33 to S36. Where informative priors were used, their distributions are shown (red lines).

**Table S24:** Gelman-Rubin and effective sample size (ESS) of pre-tunned model parameters after MCMC ("expectation" model). For each NHS England region, we present the highest Gelman-Rubin and lowest ESS values across the 85-dimensional parameter space. See corresponding convergence plots for each parameter and region below (figure S37 to figure S43).

| Diagnostic | East of England | London | Midlands | North East and Yorkshire | North West | South East | South West |
|---|---|---|---|---|---|---|---|
| $Gelman-Rubin$ | 1.09 | 1.09 | 1.05 | 1.06 | 1.08 | 1.08 | 1.09 |
| $ESS$ | 1433 | 1412 | 1479 | 1282 | 1475 | 1214 | 1272 |

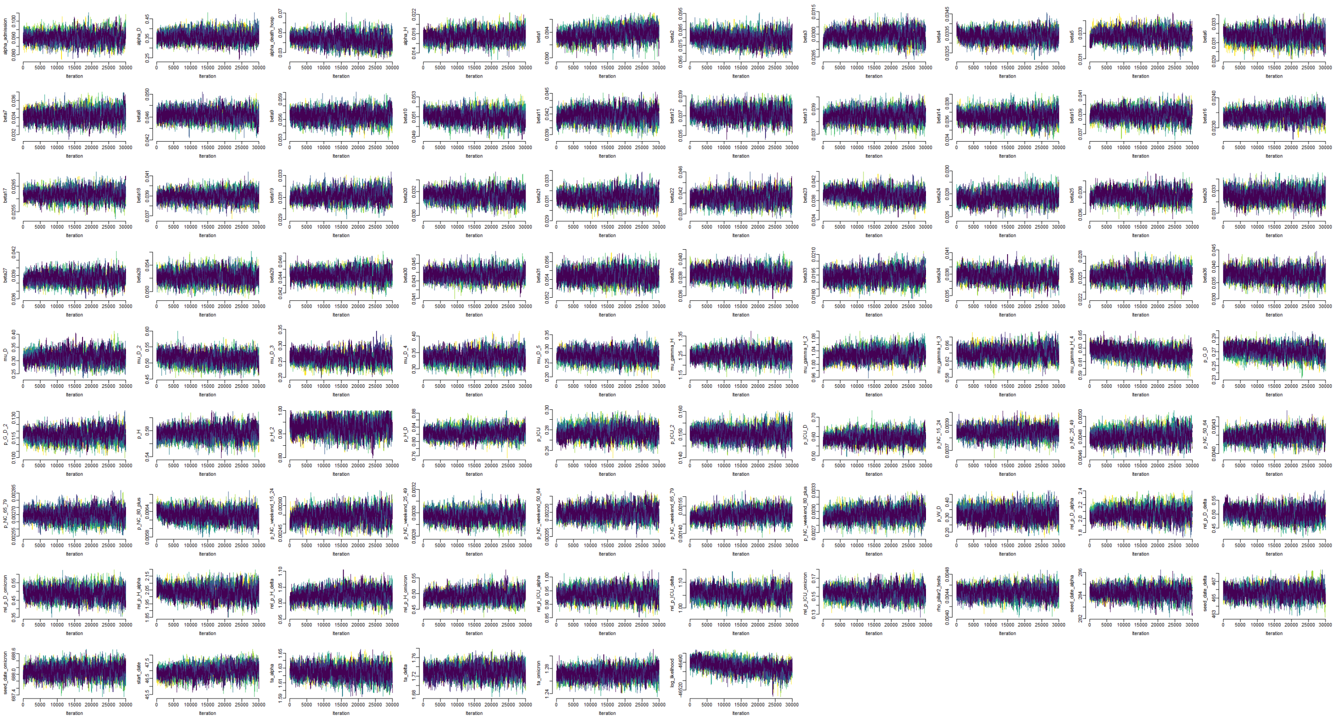

**Figure S37:** Traceplots of inferred initial parameters from MCMC pre-tuning for East of England.

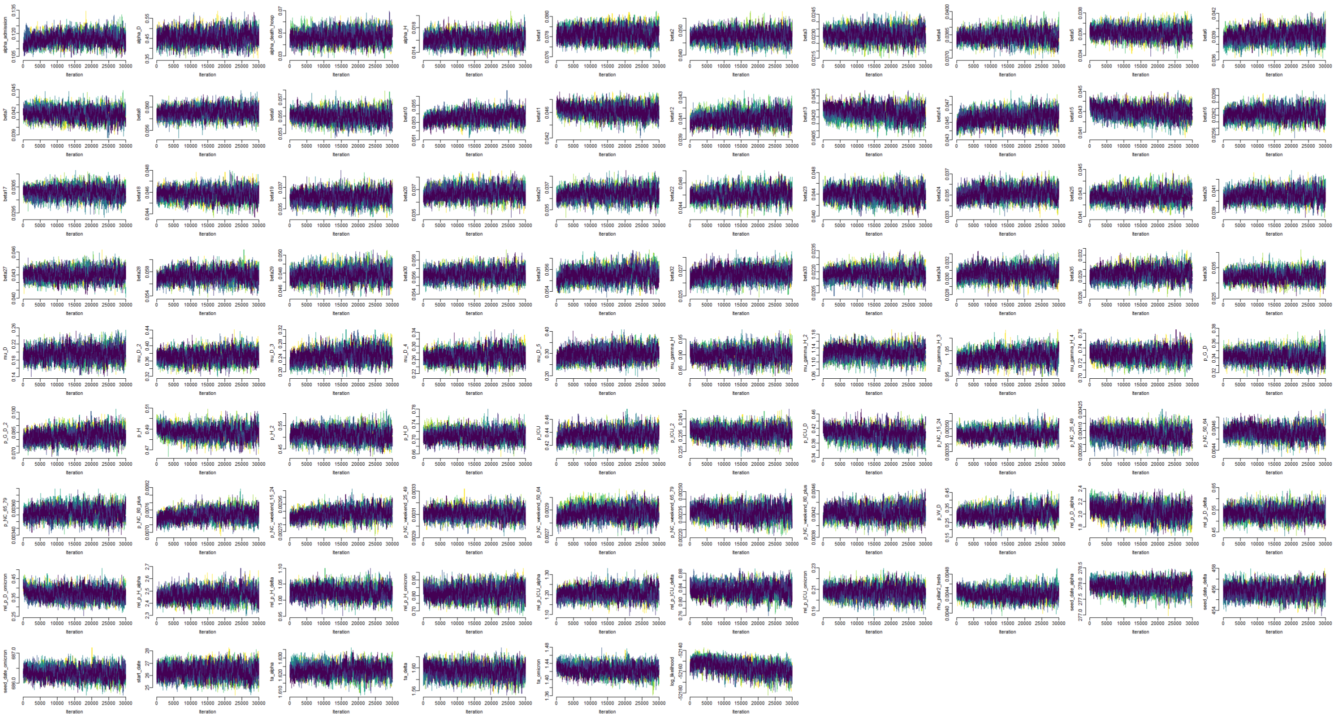

**Figure S38:** Traceplots of inferred initial parameters from MCMC pre-tuning for London.

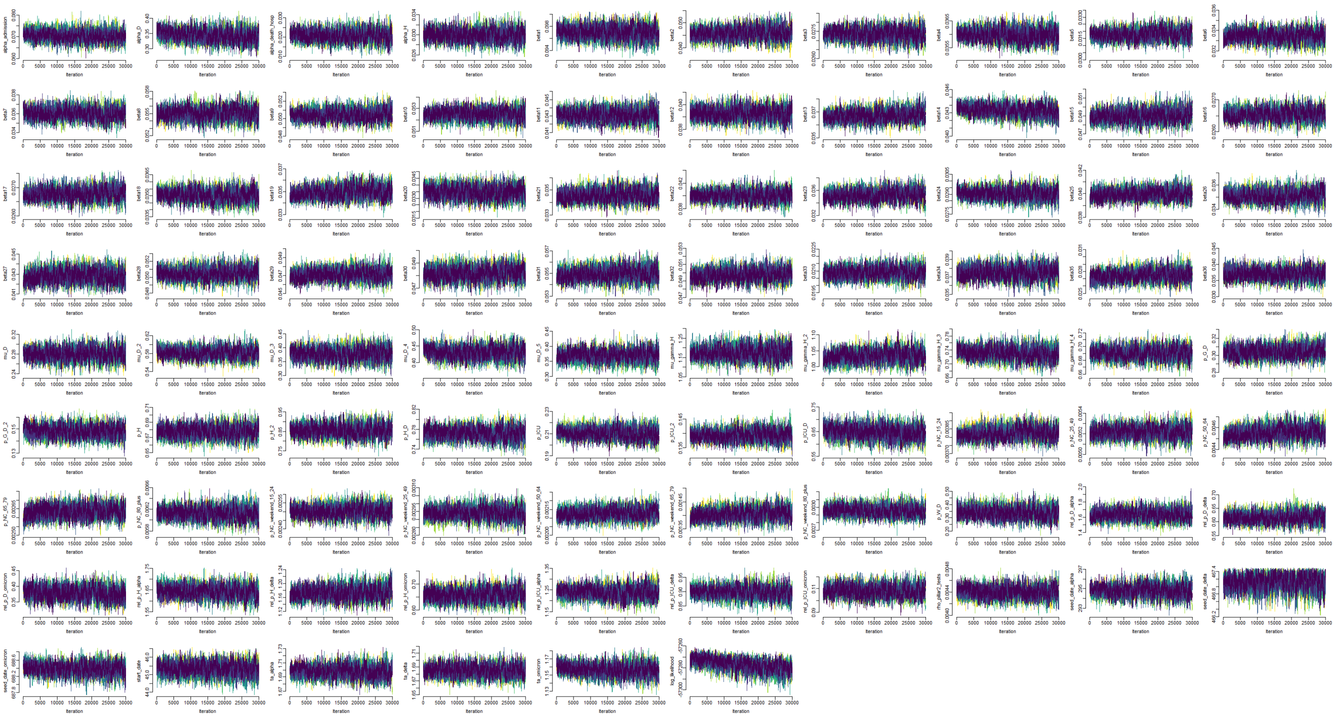

**Figure S39:** Traceplots of inferred initial parameters from MCMC pre-tuning for Midlands.

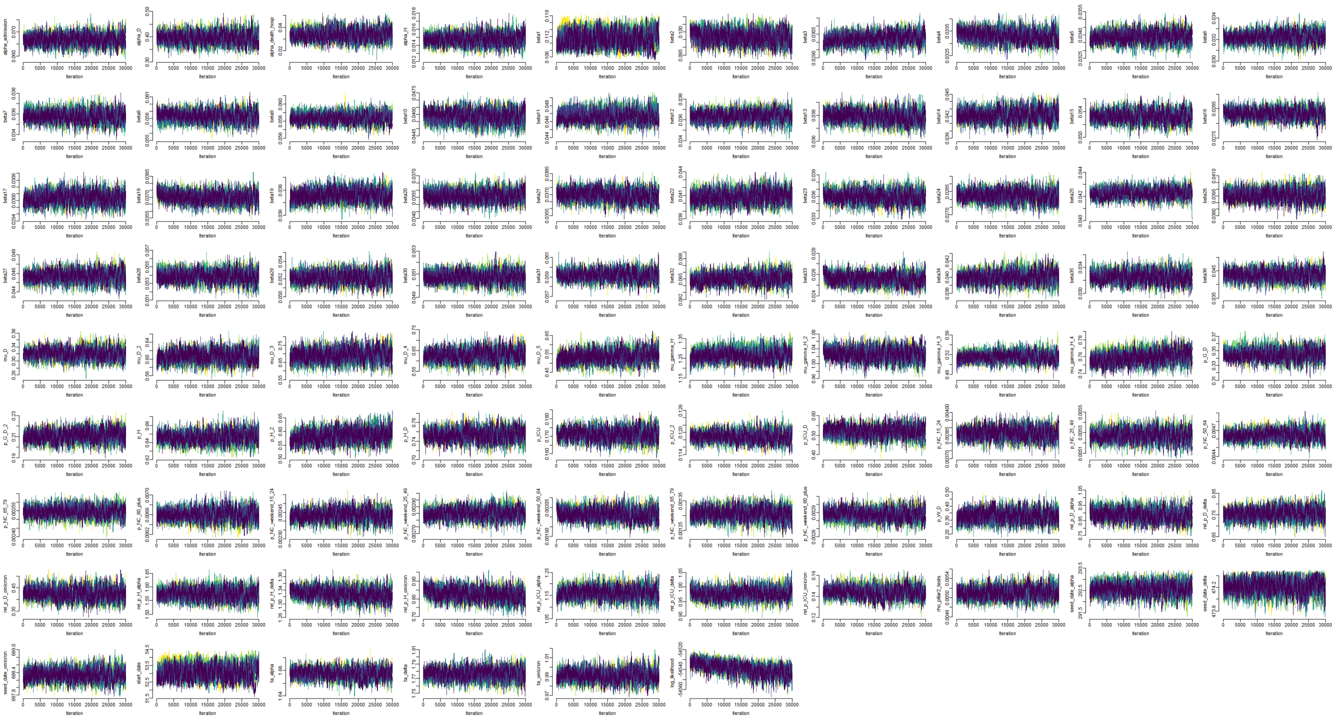

**Figure S40:** Traceplots of inferred initial parameters from MCMC pre-tuning for North East and Yorkshire.

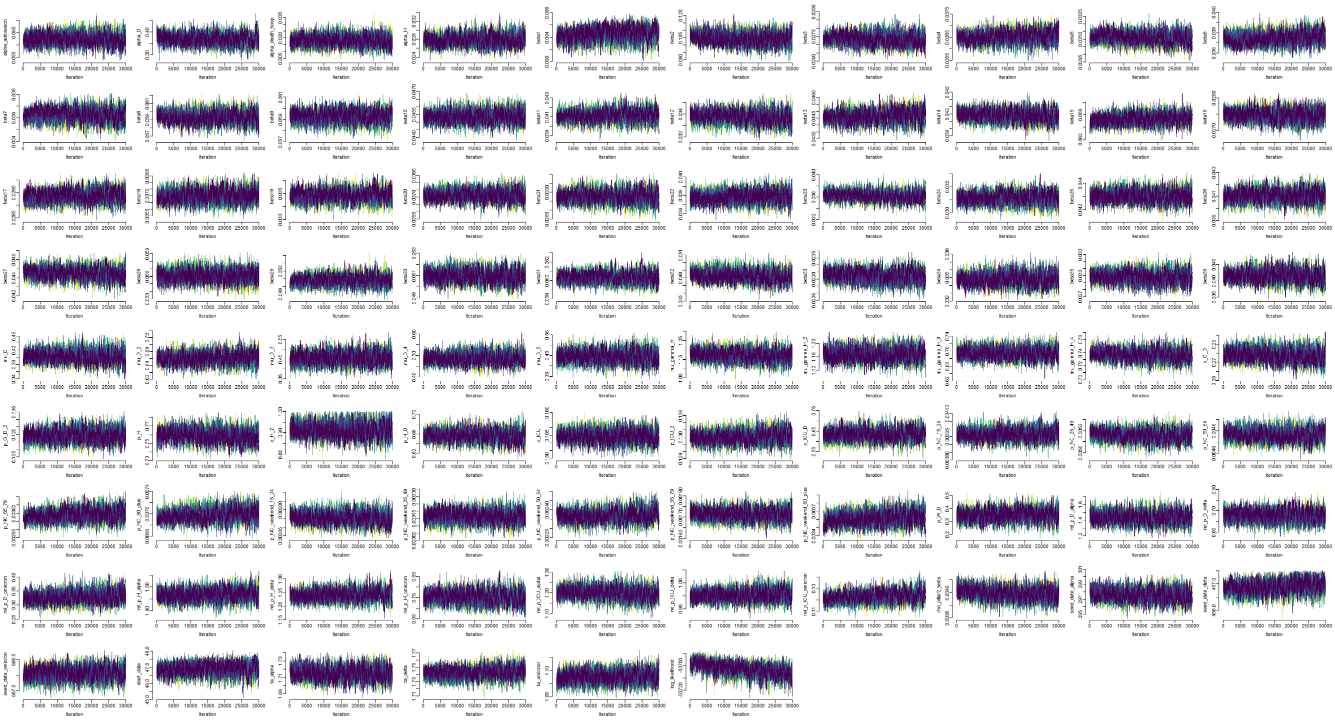

**Figure S41:** Traceplots of inferred initial parameters from MCMC pre-tuning for North West.

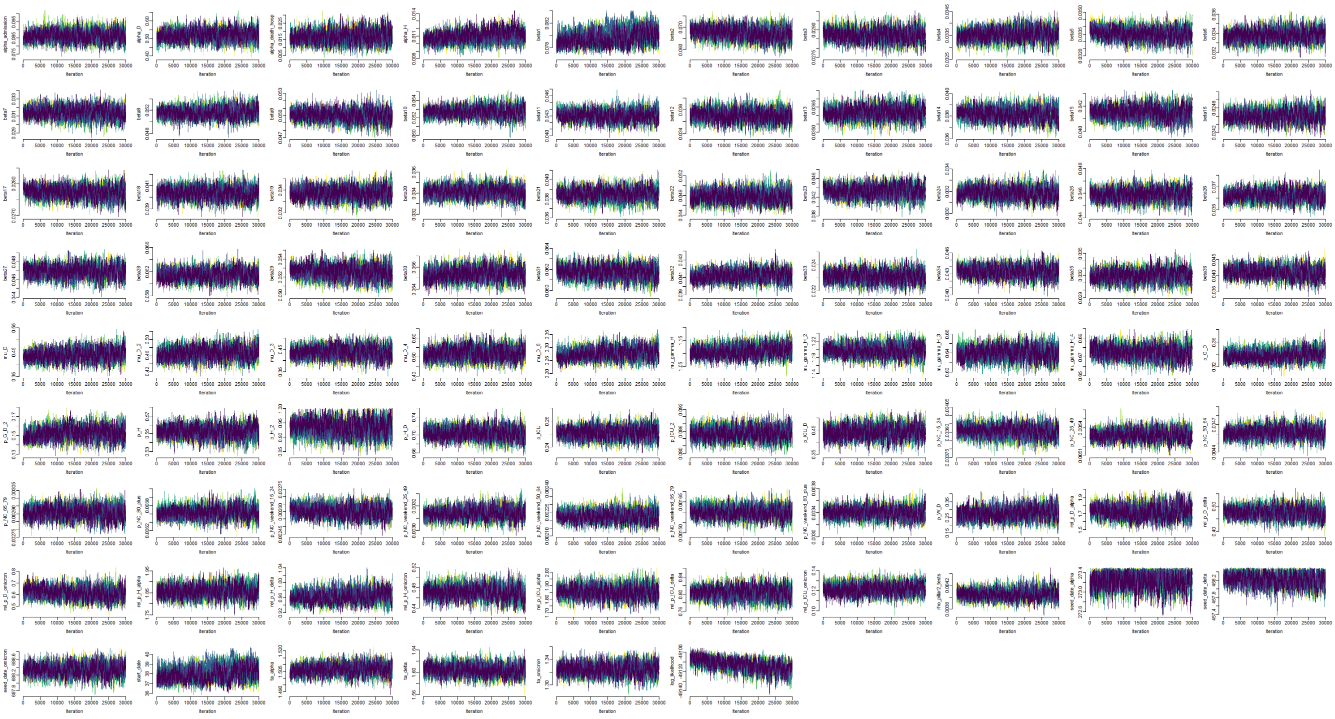

**Figure S42:** Traceplots of inferred initial parameters from MCMC pre-tuning for South East.

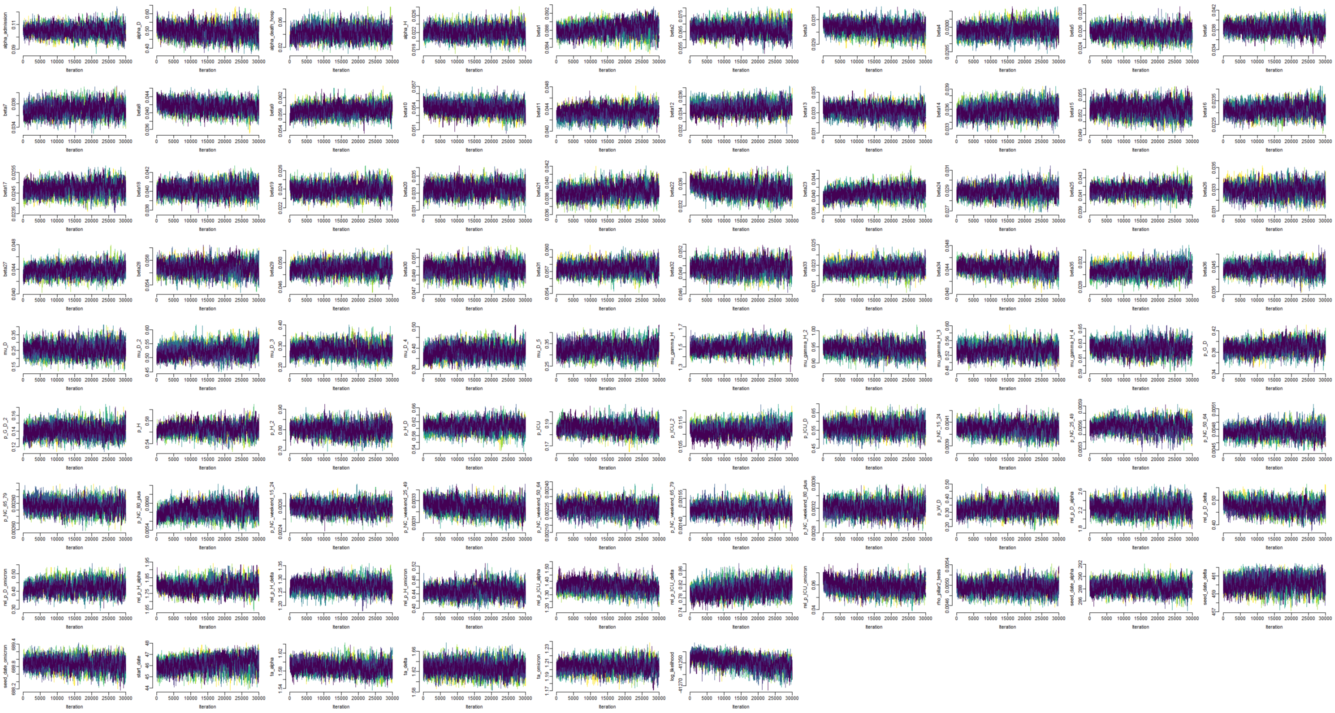

**Figure S43:** Traceplots of inferred initial parameters from MCMC pre-tuning for SouthWest.

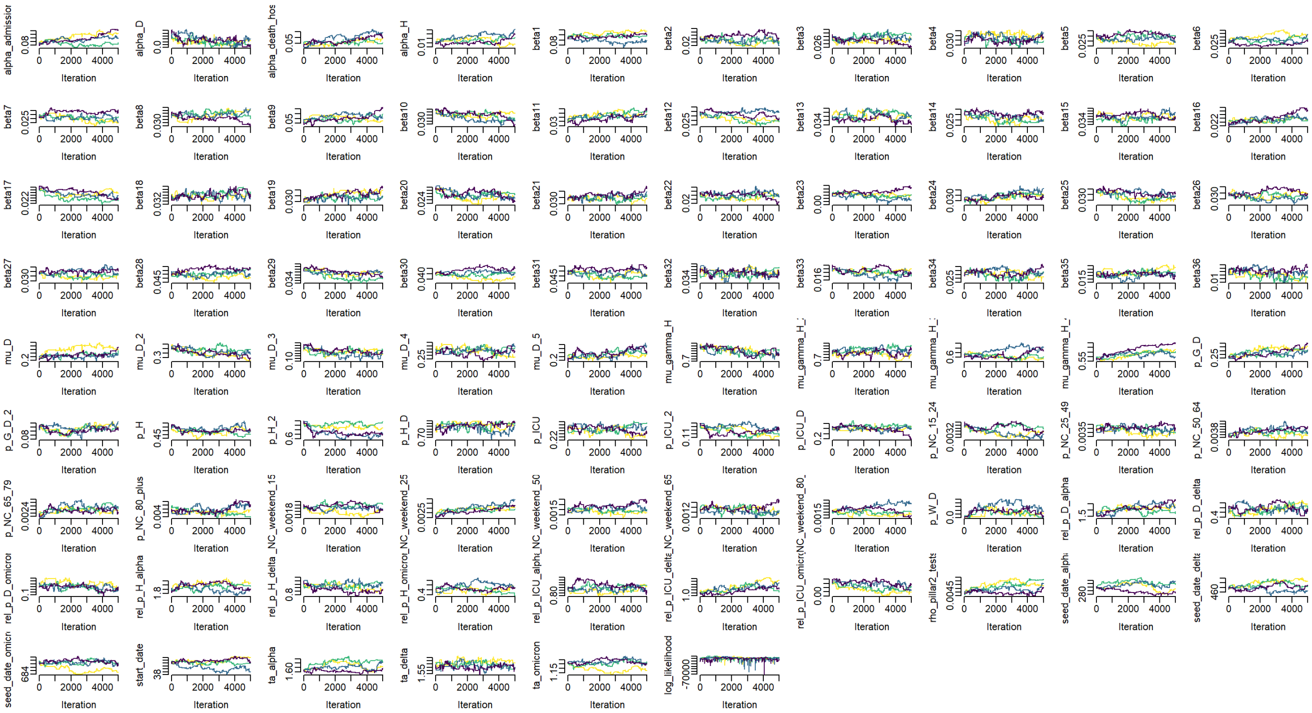

**Figure S44:** Traceplots of inferred parameters from pMCMC for East of England.

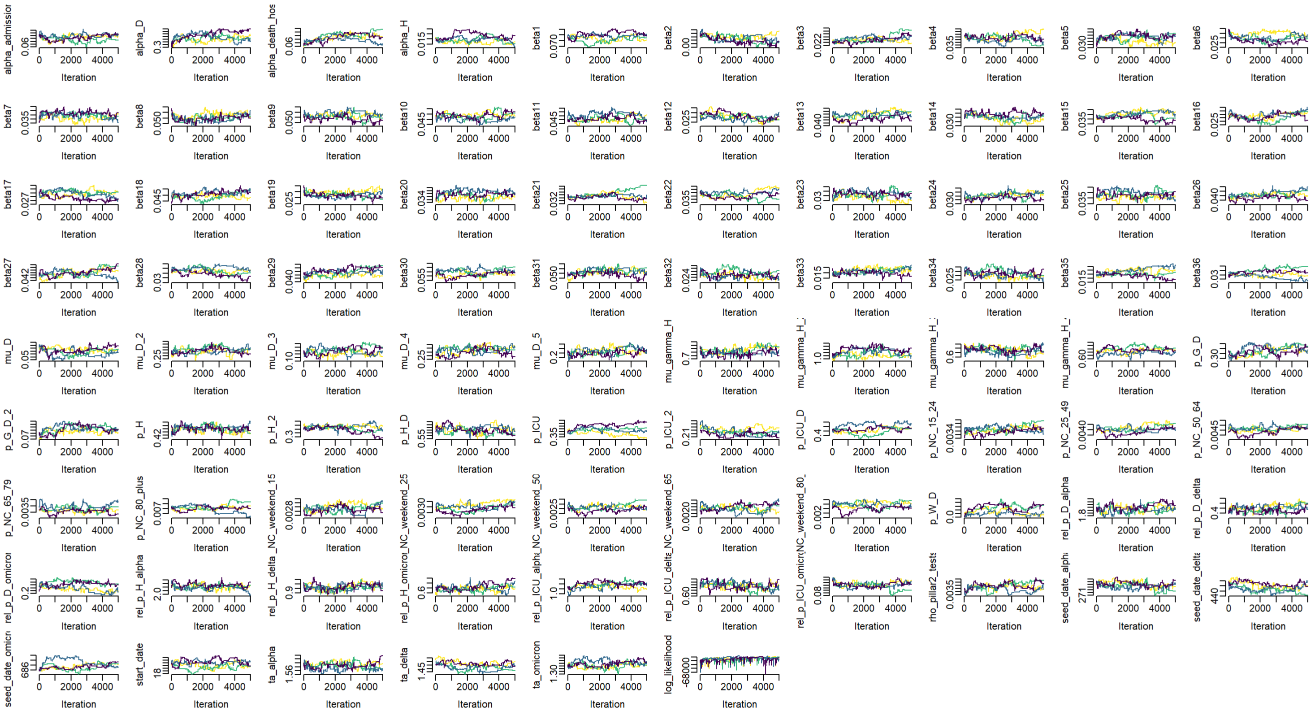

**Figure S45:** Traceplots of inferred parameters from pMCMC for London.

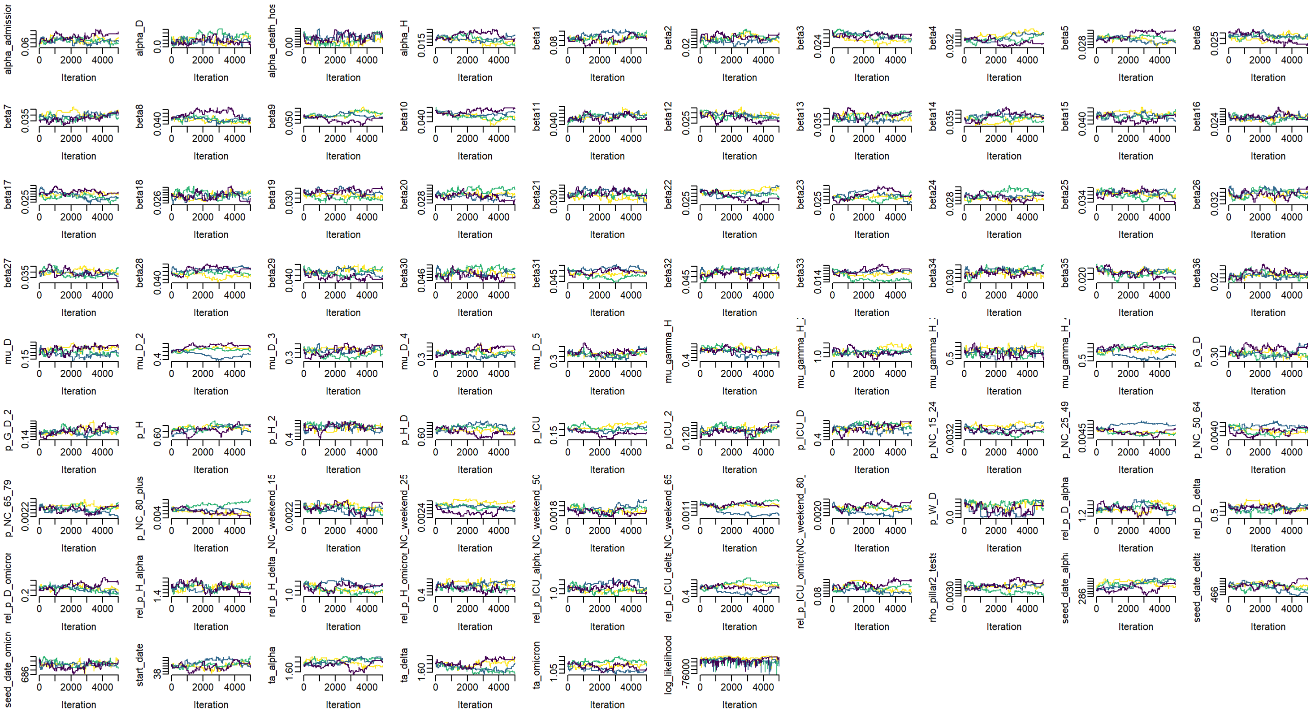

**Figure S46:** Traceplots of inferred parameters from pMCMC for Midlands.

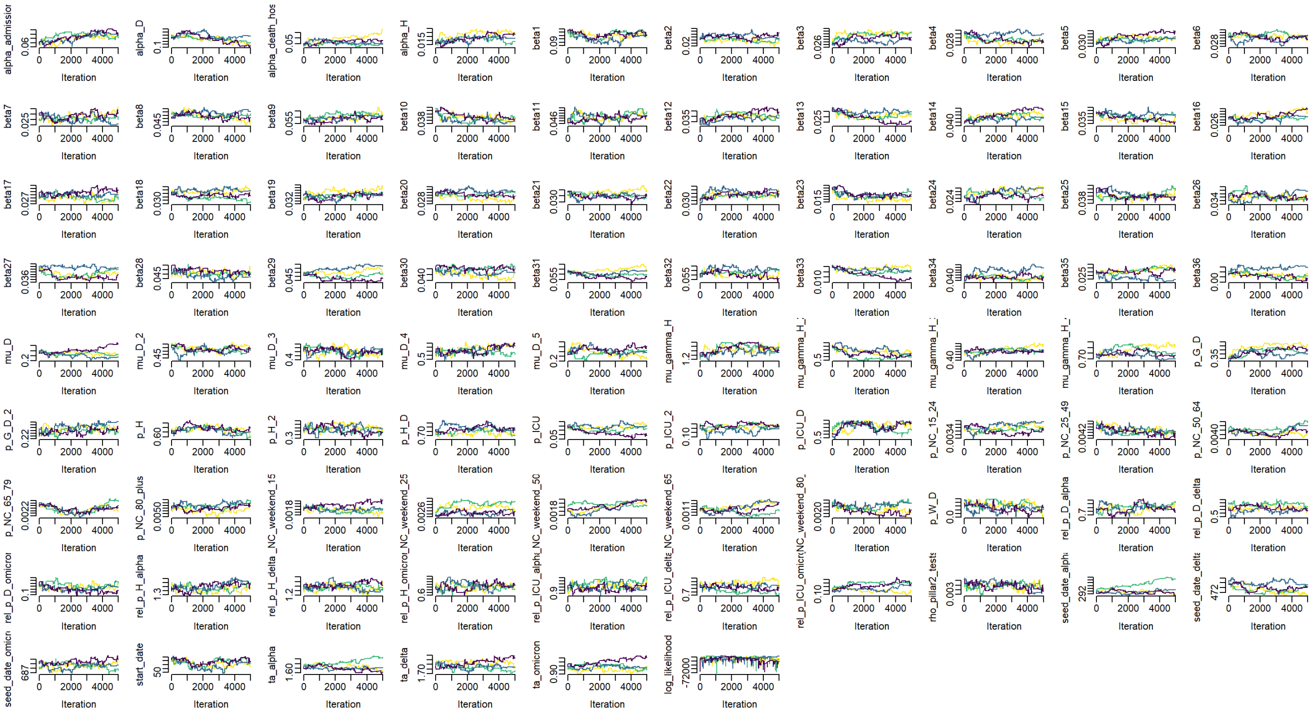

**Figure S47:** Traceplots of inferred parameters from pMCMC for North East and Yorkshire.

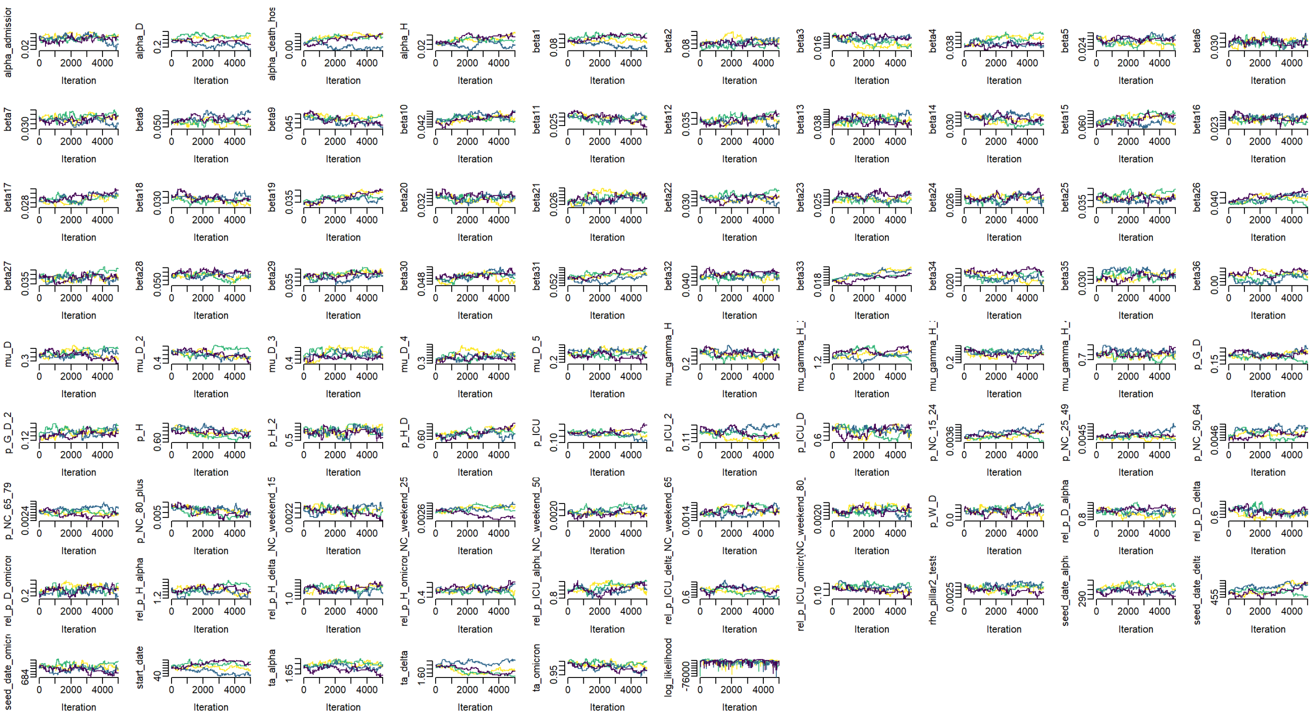

**Figure S48:** Traceplots of inferred parameters from pMCMC for North West.

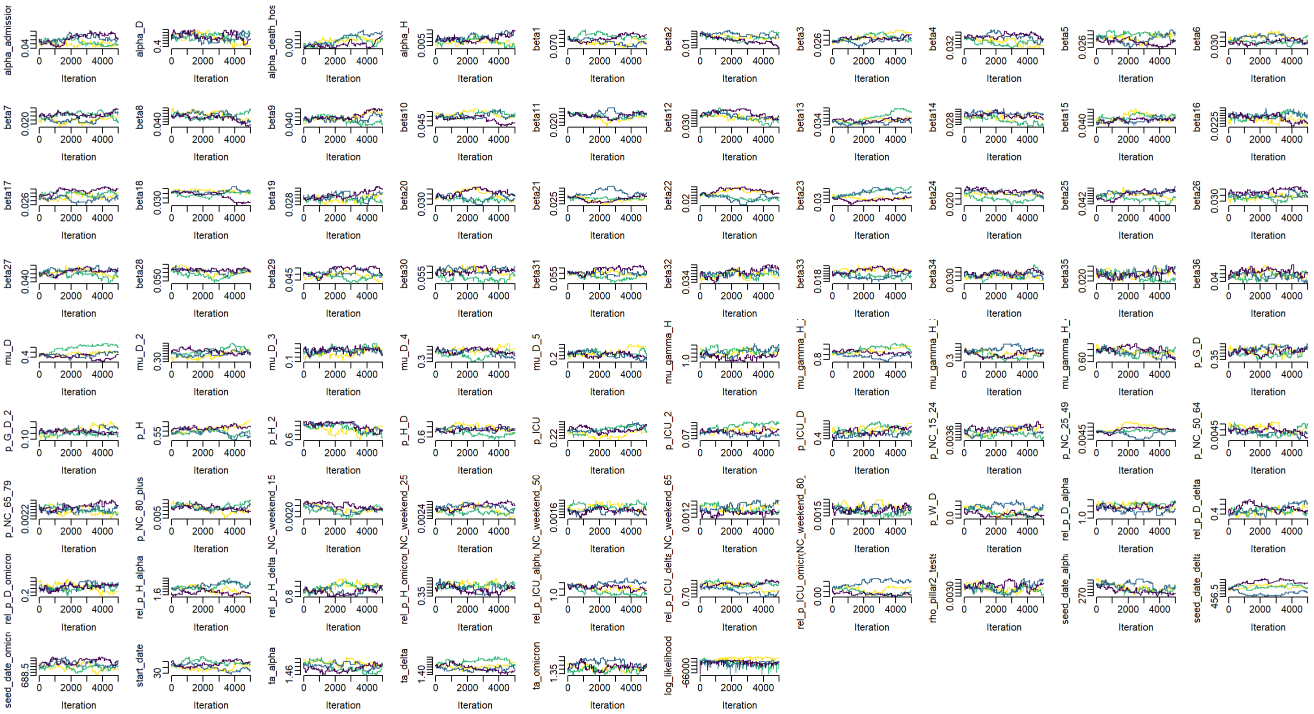

**Figure S49:** Traceplots of inferred parameters from pMCMC for South East.

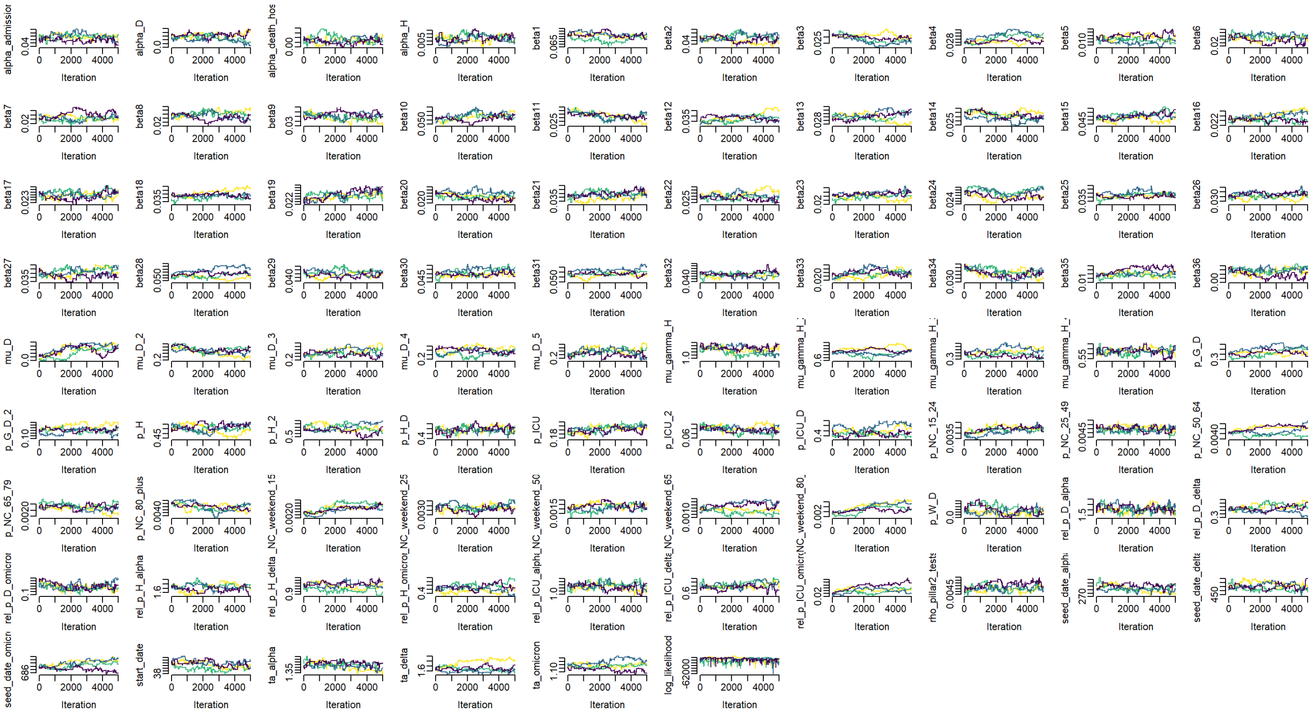

**Figure S50:** Traceplots of inferred parameters from pMCMC for South West.

**Symbol Glossary**

| Symbol | Definition |
|---|---|
| | **Abbreviations** |
| ICU | Intensive care unit |
| VE | Vaccine effectiveness |
| AZ | AstraZeneca ChAdOx1 nCoV-19 (AZD1222) vaccine |
| PF | Pfizer-BioNTech COVID-19 Vaccine BNT162b2 |
| Mod | Moderna mRNA-1273 vaccine |
| | **Model Compartments** |
| $S^{i,k}$ | Susceptible |
| $E^{i,j,k}$ | Exposed |
| $I_P^{i,j,k}$ | Infected pre-symptomatic |
| $I_A^{i,j,k}$ | Infected asymptomatic |
| $I_{C_1}^{i,j,k}$ | Symptomatic infected (infectious) |
| $I_{C_2}^{i,j,k}$ | Symptomatic infected (not infectious) |
| $G_D^{i,j,k}$ | Severe disease, not hospitalised |
| $D^{i,j,k}$ | Deceased (as a result of COVID-19) |
| $R^{i,j,k}$ | Recovered |
| $V_k$ | Vaccination strata |
| $ICU_{pre}^{i,j,k}$ | Awaiting admission to ICU |
| $ICU_{W_R}^{i,j,k}$ | Hospitalised in ICU, leading to recovery |
| $ICU_{W_D}^{i,j,k}$ | Hospitalised in ICU, leading to death following step-down from ICU |
| $ICU_D^{i,j,k}$ | Hospitalised in ICU, leading to death |
| $W_D^{i,j,k}$ | Step-down post-ICU period, leading to death |
| $W_R^{i,j,k}$ | Step-down post-ICU recovery period |
| $H_D^{i,j,k}$ | Hospitalised on general ward leading to death |
| $H_R^{i,j,k}$ | Hospitalised on general ward leading to recovery |
| | **Model Parameters** |
| $p_H^{i,j,k}(t)$ | Probability of hospitalisation given symptomatic |
| $p_{G_D}^{i,j,k}(t)$ | Probability of dying in the community/care home given severe disease requiring hospitalisation |
| $p_{ICU}^{i,j}(t)$ | Probability of ICU admission given hospitalised |
| $p_{H_D}^{i,j,k}(t)$ | Probability of death given hospitalised and not in ICU |
| $p_{ICU_D}^{i,j,k}(t)$ | Probability of death given ICU |
| $p_{W_D}^{i,j,k}(t)$ | Probability of death after discharge |
| $\chi^{i,j,k}(t)$ | Susceptibility of an individual to variant $j$ given vaccine stratum $k$ |
| $\xi^{i,j,k}(t)$ | Infectivity of an individual infected with variant $j$ given vaccine stratum $k$ |
| $\lambda^{i,j,k}(t)$ | Variant-specific force of infection |
| $\Lambda^{i,k}(t)$ | Combined force of infection (both variants) |
| $\zeta^{i,k}(t)$ | Rate of progression from vaccine strata $k$ to $k+1$ |
| $\gamma_x$ | Rate of progression from compartment $x$ |
| $R_t^j$ | Reproduction number for variant $j$ at time $t$ |
| $R_t^{j,eff}$ | Effective reproduction number for variant $j$ at time $t$ |
| $t_{Wiltype}$ | Region specific outbreak start time |
| $t_{Alpha}$ | Region specific Alpha seeding time |
| $t_{Delta}$ | Region specific Delta seeding time |
| $t_{Omicron}$ | Region specific Omicron seeding time |
| $\nu_j$ | Duration of seeding period for variant $j$ |
| $\phi_j$ | Daily seeding rate for variant $j$ |
| $\delta^{i,j,k}(t)$ | Daily seeding rate of variant $j$ (stratified by age and vaccination strata) |
| $\sigma$ | Delta transmission advantage |
| $m_{i,i'}(t)$ | Person-to-person transmission rate |
| $c_{i,i'}$ | Person-to-person contact rate |
| Continued on next page | |

| **Table S25 – continued from previous page** | |
|---|---|
| Symbol | Definition |
| $\beta(t)$ | Transmission rate |
| $\beta_i$ | Transmission rate at change-point $t_i$ |
| $\Theta_{i,j,k}(t)$ | Weighted number of infectious individuals |
| $\Delta_I^{i,k}$ | Mean duration of infectiousness weighted by infectivity |
| | **Vaccine Effectiveness vs.** |
| $e_{inf}$ | Infection |
| $e_{SD}$ | Severe disease |
| $e_{death}$ | Death |
| $e_{SD\|sympt}$ | Severe disease given symptoms |
| $e_{death\|SD}$ | Death given severe disease |
| $e_{ins}$ | Infectiousness |
| | **Fixed Parameters** |
| $p_C^i$ | Probability of being symptomatic given infected |
| $p^*(t)$ | Probability of COVID-19 diagnosis confirmed prior to hospital admission |
| $\gamma_U$ | Rate at which unconfirmed hospital patients are confirmed as infected |
| $\gamma_x$ | Rate at which individuals move out of compartment x |
| $p_{sero_{pos}}$ | Probability of seroconversion following infection |
| $p_{sero_{spec}}$ | Specificity of serology test |
| $p_{sero_{sens}}$ | Sensitivity of serology test |
| $1/\gamma_{sero_{pre}}$ | Mean time to seroconversion from onset of infectiousness |
| $1/\gamma_{sero^1_{pos}}$ | Mean duration of seropositivity (Euroimmun assay) |
| $1/\gamma_{sero^2_{pos}}$ | Mean duration of seropositivity (Roche N) |
| $\eta_j$ | Probability of cross-immunity to variant $j$ following infection from a variant predating variant $j$ |
| $\theta_{I_A}$ | Infectivity of an asymptomatic individual, relative to a symptomatic one |

### References

[1] Knock ES, Whittles LK, Lees JA, Perez-Guzman PN, Verity R, FitzJohn RG, et al. Key epidemiological drivers and impact of interventions in the 2020 SARS-CoV-2 epidemic in England;13(602). Publisher: American Association for the Advancement of Science _eprint: https://stm.sciencemag.org/content/13/602/eabg4262.full.pdf. Available from: https://stm.sciencemag.org/content/13/602/eabg4262.

[2] Sonabend R, Whittles LK, Imai N, Perez-Guzman PN, Knock ES, Rawson T, et al. Non-pharmaceutical interventions, vaccination, and the SARS-CoV-2 delta variant in England: a mathematical modelling study;398(10313):1825-35. Publisher: Elsevier B.V. Available from: http://www.thelancet.com/article/S0140673621022765/fulltexthttp://www.thelancet.com/article/S0140673621022765/abstracthttps://www.thelancet.com/journals/lancet/article/PIIS0140-6736(21)02276-5/abstract.

[3] Imai N, Rawson T, Knock ES, Sonabend R, Elmaci Y, Perez-Guzman PN, et al. Quantifying the impact of delaying the second COVID-19 vaccine dose in England: a mathematical modelling study;8. Available from: https://www.thelancet.com/journals/lanpub/article/PIIS2468-2667(22)00337-1/fulltext.

[4] Mossong J, Hens N, Jit M, Beutels P, Auranen K, Mikolajczyk R, et al. Social contacts and mixing patterns relevant to the spread of infectious diseases;5(3):e74. Publisher: Public Library of Science.

[5] Whittles LK, Imai N, Knock ES, Perez-Guzman PN, Ghani A, Ferguson NM, et al.. Potential profile of the COVID-19 epidemic in the UK under different vaccination roll out strategies [Ver 2.];. Available from: https://assets.publishing.service.gov.uk/government/uploads/system/uploads/attachment_data/file/958913/S1024_SPI-M_vaccination_ask_Imperial_College.pdf.

[6] Whittles LK, Imai N, Sonabend R, Knock ES, Perez-Guzman PN, Hogan AB, et al.. Evaluating England's Roadmap out of Lockdown;. Available from: https://assets.publishing.service.gov.uk/government/uploads/system/uploads/attachment_data/file/1000511/S1183_SPI-M_Imperial.pdf.

[7] Sonabend R, Imai N, Knock ES, Perez-Guzman PN, Whittles LK, Rawson T, et al.. Evaluating the Roadmap out of Lockdown for England: modelling the delayed step 4 of the roadmap in the context of the Delta variant;. Available from: https://assets.publishing.service.gov.uk/government/uploads/system/uploads/attachment_data/file/1001177/S1303_Imperial_College_London_Evaluating_the_Roadmap_out_of_Lockdown_for_England_modelling_the_delayed_step_4.2_of_the_roadmap_in_the_context_of_the_Delta_variant__7_July_2021__1_.pdf.

[8] Perez-Guzman PN, Imai N, Knock ES, Rawson T, Sonabend R, Kanapram DT, et al.. Autumn and Winter 2021-2022: potential COVID-19 epidemic trajectories;. Available from: https://assets.publishing.service.gov.uk/government/uploads/system/uploads/attachment_data/file/1027920/S1386_SPI-M_potential_winter_trajectories_Imperial_College.pdf.

[9] UK Government. Regulatory approval of COVID-19 Vaccine AstraZeneca;. Available from: https://www.gov.uk/government/publications/regulatory-approval-of-covid-19-vaccine-astrazeneca.

[10] UK Government. Regulatory approval of Pfizer/BioNTech vaccine for COVID-19;. Available from: https://www.gov.uk/government/publications/regulatory-approval-of-pfizer-biontech-vaccine-for-covid-19.

[11] UK Government. Moderna vaccine becomes third COVID-19 vaccine approved by UK regulator;. Available from: https://www.gov.uk/government/news/moderna-vaccine-becomes-third-covid-19-vaccine-approved-by-uk-regulator.

[12] Kojima N, Shrestha N, Klausner JD. A systematic review of the protective effect of prior SARS-CoV-2 infection on repeat infection;44(4):327-32. Publisher: SAGE Publications Sage CA: Los Angeles, CA.

[13] Coronavirus (COVID-19) in the UK;. Available from: https://coronavirus.data.gov.uk.

[14] Riley S, Walters CE, Wang H, Eales O, Ainslie KE, Atchinson C, et al. REACT-1 round 7 updated report: regional heterogeneity in changes in prevalence of SARS-CoV-2 infection during the second national COVID-19 lockdown in England. Publisher: Cold Spring Harbor Laboratory Press.

[15] Office for National Statistics. Coronavirus (COVID-19) latest insights: Infections;. Available from: https://www.ons.gov.uk/peoplepopulationandcommunity/healthandsocialcare/conditionsanddiseases/articles/coronaviruscovid19latestinsights/infections.

[16] UK Government. Sero-surveillance of COVID-19;. Available from: https://www.gov.uk/government/publications/national-covid-19-surveillance-reports/sero-surveillance-of-covid-19.

[17] UK Health Security Agency. Variants: distribution of cases data, 20 May 2021 - GOV.UK;. Available from: https://www.gov.uk/government/publications/covid-19-variants-genomically-confirmed-case-numbers/variants-distribution-of-cases-data.

[18] Svensson A note on generation times in epidemic models;208(1):300-11. Available from: https://linkinghub.elsevier.com/retrieve/pii/S0025556406002094.

[19] Park SW, Bolker BM, Funk S, Metcalf CJE, Weitz JS, Grenfell BT, et al. The importance of the generation interval in investigating dynamics and control of new SARS-CoV-2 variants;19(191):20220173. Available from: https://royalsocietypublishing.org/doi/10.1098/rsif.2022.0173.

[20] Challen R, Brooks-Pollock E, Tsaneva-Atanasova K, Danon L. Meta-analysis of the severe acute respiratory syndrome coronavirus 2 serial intervals and the impact of parameter uncertainty on the coronavirus disease 2019 reproduction number;31(9):1686-703. Available from: http://journals.sagepub.com/doi/10.1177/09622802211065159.

[21] an der Heiden M, Buchholz U. Serial interval in households infected with SARS-CoV-2 variant B.1.1.529 (Omicron) is even shorter compared to Delta;150:e165. Available from: https://www.cambridge.org/core/product/identifier/S0950268822001248/type/journal_article.

[22] Geismar C, Fragaszy E, Nguyen V, Fong WLE, Shrotri M, Beale S, et al. Household serial interval of COVID-19 and the effect of Variant B.1.1.7: analyses from prospective community cohort study (Virus Watch);6:224. Available from: https://wellcomeopenresearch.org/articles/6-224/v2.

[23] Hart WS, Miller E, Andrews NJ, Waight P, Maini PK, Funk S, et al. Generation time of the alpha and delta SARS-CoV-2 variants: an epidemiological analysis;22(5):603-10. Available from: https://linkinghub.elsevier.com/retrieve/pii/S1473309922000019.

[24] Edara VV, Lai L, Sahoo M, Floyd K, Sibai M, Solis D, et al. Infection and vaccine-induced neutralizing antibody responses to the SARS-CoV-2 B. 1.617. 1 variant. Publisher: Cold Spring Harbor Laboratory.

[25] Mulligan MJ, Lyke KE, Kitchin N, Absalon J, Gurtman A, Lockhart S, et al. Phase I/II study of COVID-19 RNA vaccine BNT162b1 in adults;586(7830):589-93. Publisher: Nature Publishing Group.

[26] Ramasamy MN, Minassian AM, Ewer KJ, Flaxman AL, Folegatti PM, Owens DR, et al. Safety and immunogenicity of ChAdOx1 nCoV-19 vaccine administered in a prime-boost regimen in young and old adults (COV002): a single-blind, randomised, controlled, phase 2/3 trial;396(10267):1979-93. Publisher: Elsevier.

[27] COVID-19 vaccination programme: FAQs on second doses;. Accessed: 2021-05-20. https://www.england.nhs.uk/coronavirus/publication/covid-19-vaccination-programme-faqs-on-second-doses/.

[28] Voysey M, Clemens SAC, Madhi SA, Weckx LY, Folegatti PM, Aley PK, et al. Safety and efficacy of the ChAdOx1 nCoV-19 vaccine (AZD1222) against SARS-CoV-2: an interim analysis of four randomised controlled trials in Brazil, South Africa, and the UK;397(10269):99-111. Publisher: Elsevier.

[29] Polack FP, Thomas SJ, Kitchin N, Absalon J, Gurtman A, Lockhart S, et al. Safety and efficacy of the BNT162b2 mRNA Covid-19 vaccine;383(27):2603-15. Publisher: Mass Medical Soc.

[30] Harris RJ, Hall JA, Zaidi A, Andrews NJ, Dunbar JK, Dabrera G. Impact of vaccination on household transmission of SARS-COV-2 in England.

[31] Bernal JL, Andrews N, Gower C, Robertson C, Stowe J, Tessier E, et al. Effectiveness of the Pfizer-BioNTech and Oxford-AstraZeneca vaccines on covid-19 related symptoms, hospital admissions, and mortality in older adults in England: test negative case-control study;373. Publisher: British Medical Journal Publishing Group.

[32] Ismail S, Vilaplana T, Elgohari S, others. Effectiveness of BNT162b2 mRNA and ChAdOx1 adenovirus vector COVID-19 vaccines on risk of hospitalisation among older adults in England: an observational study using surveillance data;.

[33] Hogan AB, Wu SL, Doohan P, Watson OJ, Winskill P, Charles G, et al. The value of vaccine booster doses to mitigate the global impact of the Omicron SARS-CoV-2 variant:2022.01.17.22269222. Publisher: Cold Spring Harbor Laboratory Press. Available from: https://www.medrxiv.org/content/10.1101/2022.01.17.22269222v1https://www.medrxiv.org/content/10.1101/2022.01.17.22269222v1.abstract.

[34] Khoury DS, Cromer D, Reynaldi A, Schlub TE, Wheatley AK, Juno JA, et al. Neutralizing antibody levels are highly predictive of immune protection from symptomatic SARS-CoV-2 infection;27(7):1205-11. Publisher: Nature Publishing Group. Available from: https://www.nature.com/articles/s41591-021-01377-8.

[35] Andrews N, Tessier E, Stowe J, Gower C, Kirsebom F, Simmons R, et al. Vaccine effectiveness and duration of protection of Comirnaty, Vaxzevria and Spikevax against mild and severe COVID-19 in the UK. Publisher: Cold Spring Harbor Laboratory Press.

[36] Vasileiou E, Simpson CR, Shi T, Kerr S, Agrawal U, Akbari A, et al. Interim findings from first-dose mass COVID-19 vaccination roll-out and COVID-19 hospital admissions in Scotland: a national prospective cohort study. Publisher: Elsevier.

[37] Public Health England vaccine effectiveness report;. Accessed: 2021-05-20. https://assets.publishing.service.gov.uk/government/uploads/system/uploads/attachment_data/file/971017/SP_PH__VE_report_20210317_CC_JLB.pdf.

[38] Hyams C, Marlow R, Maseko Z, King J, Ward L, Fox K, et al. Assessing the effectiveness of bnt162b2 and chadox1ncov-19 covid-19 vaccination in prevention of hospitalisations in elderly and frail adults: A single centre test negative case-control study.

[39] Stowe J, Andrews N, Gower C, Gallagher E, Utsi L, Simmons R. Effectiveness of COVID-19 vaccines against hospital admission with the Delta (B. 1.617. 2) variant.

[40] Hall VJ, Foulkes S, Saei A, Andrews N, Oguti B, Charlett A, et al. Effectiveness of BNT162b2 mRNA vaccine against infection and COVID-19 vaccine coverage in healthcare workers in England, multicentre prospective cohort study (the SIREN Study).

[41] UK Government. COVID-19 vaccine surveillance report: week 48;. Available from: https://assets.publishing.service.gov.uk/government/uploads/system/uploads/attachment_data/file/1121345/vaccine-surveillance-report-week-48-2022.pdf.

[42] Sheikh A, McMenamin J, Taylor B, Robertson C. SARS-CoV-2 Delta VOC in Scotland: demographics, risk of hospital admission, and vaccine effectiveness. Publisher: Elsevier.

[43] Bernal JL, Andrews N, Gower C, Gallagher E, Simmons R, Thelwall S, et al. Effectiveness of Covid-19 vaccines against the B. 1.617. 2 (Delta) variant;385(7):385-94. Publisher: Mass Medical Soc.

[44] Voysey M, Clemens SAC, Madhi SA, Weckx LY, Folegatti PM, Aley PK, et al. Single-dose administration and the influence of the timing of the booster dose on immunogenicity and efficacy of ChAdOx1 nCoV-19 (AZD1222) vaccine: a pooled analysis of four randomised trials;397(10277):881-91. Publisher: Elsevier.

[45] Nasreen S, Chung H, He S, Brown KA, Gubbay JB, Buchan SA, et al. Effectiveness of COVID-19 vaccines against symptomatic SARS-CoV-2 infection and severe outcomes with variants of concern in Ontario;7(3):379-85. Publisher: Nature Publishing Group. Available from: https://www.nature.com/articles/s41564-021-01053-0.

[46] Andrews N, Stowe J, Kirsebom F, Toffa S, Rickeard T, Gallagher E, et al. Covid-19 vaccine effectiveness against the Omicron (B. 1.1. 529) variant;386(16):1532-46. Publisher: Mass Medical Soc.

[47] Institute for Government. Coronavirus vaccine rollout;. Available from: https://www.instituteforgovernment.org.uk/explainers/coronavirus-vaccine-rollout.

[48] UK Government. COVID-19 Greenbook chapter 14a;. Available from: https://assets.publishing.service.gov.uk/government/uploads/system/uploads/attachment_data/file/984310/Greenbook_chapter_14a_7May2021.pdf.

[49] Funk S. Socialmixr: Social mixing matrices for infectious disease modelling. Publisher: The Comprehensive R Archive Network.

[50] COVID-19 Hospital Activity;. Available from: https://www.england.nhs.uk/statistics/statistical-work-areas/covid-19-hospital-activity/.

[51] SGSS and CHESS data;. Available from: https://digital.nhs.uk/about-nhs-digital/corporate-information-and-documents/directions-and-data-provision-notices/data-provision-notices-dpns/sgss-and-chess-data.

[52] Regulatory approval of Ronapreve;.

[53] UK Government. Regulatory approval of Lagrevio (molnupiravir);. Available from: https://www.gov.uk/government/publications/regulatory-approval-of-lagevrio-molnupiravir.

[54] UK Government. Regulatory approval of Paxlovid;. Available from: https://www.gov.uk/government/publications/regulatory-approval-of-paxlovid.

[55] UK Government. Regulatory approval of Xevudy (sotrovimab);. Available from: https://www.gov.uk/government/publications/regulatory-approval-of-xevudy-sotrovimab.

[56] Docherty AB, Mulholland RH, Lone NI, Cheyne CP, De Angelis D, et al. Changes in in-hospital mortality in the first wave of COVID-19: a multicentre prospective observational cohort study using the WHO Clinical Characterisation Protocol UK. The Lancet. 2021;9.

[57] UK Government. Dexamethasone in the treatment of COVID-19: Implementation and management of supply for treatment in hospitals;. Available from: https://www.cas.mhra.gov.uk/ViewandAcknowledgment/ViewAlert.aspx?AlertID=103054.

[58] Lauer SA, Grantz KH, Bi Q, Jones FK, Zheng Q, Meredith HR, et al. The incubation period of coronavirus disease 2019 (COVID-19) from publicly reported confirmed cases: estimation and application;172(9):577-82. Publisher: American College of Physicians.

[59] Gillespie DT. Approximate accelerated stochastic simulation of chemically reacting systems;115(4):1716-33. Publisher: American Institute of Physics.

[60] Diekmann O, Heesterbeek JAP, Metz JA. On the definition and the computation of the basic reproduction ratio R 0 in models for infectious diseases in heterogeneous populations;28(4):365-82. Publisher: Springer.

[61] Brazeau N, Verity R, Jenks S, Fu H, Whittaker C, Winskill P, et al. Report 34: COVID-19 infection fatality ratio: estimates from seroprevalence.

[62] Borremans B, Gamble A, Prager K, Helman SK, McClain AM, Cox C, et al. Quantifying antibody kinetics and RNA detection during early-phase SARS-CoV-2 infection by time since symptom onset;9:e60122. Publisher: eLife Sciences Publications Limited.

[63] Harris RJ, Whitaker HJ, Andrews NJ, Aiano F, Amin-Chowdhury Z, Flood J, et al. Serological surveillance of SARS-CoV-2: Six-month trends and antibody response in a cohort of public health workers;82(5):162-9. Publisher: Elsevier.

[64] Chen S, Flegg JA, White LJ, Aguas R. Levels of SARS-CoV-2 population exposure are considerably higher than suggested by seroprevalence surveys. Publisher: Cold Spring Harbor Laboratory Press.

[65] Planas D, Veyer D, Baidaliuk A, Staropoli I, Guivel-Benhassine F, Rajah MM, et al. Reduced sensitivity of SARS-CoV-2 variant Delta to antibody neutralization:1-7. Publisher: Nature Publishing Group.

[66] Kim P, Gordon SM, Sheehan MM, Rothberg MB. Duration of SARS-CoV-2 Natural Immunity and Protection against the Delta Variant: A Retrospective Cohort Study.

[67] Planas D, Saunders N, Maes P, Guivel-Benhassine F, Planchais C, others. Considerable escape of SARS-CoV-2 Omicron to antibody neutralization. Publisher: Nature Publishing Group.

[68] Chen J, Wang R, Gilby NB, Wei GW. Omicron Variant (B.1.1.529): Infectivity, Vaccine Breakthrough, and Antibody Resistance;62:412-22. Publisher: J Chem Inf Model.

[69] Wu JT, Leung K, Leung GM. Nowcasting and forecasting the potential domestic and international spread of the 2019-nCoV outbreak originating in Wuhan, China: a modelling study;395(10225):689-97. Available from: https://www.sciencedirect.com/science/article/pii/S0140673620302609.

[70] Flaxman S, Mishra S, Gandy A, Unwin HJT, Mellan TA, Coupland H, et al. Estimating the effects of non-pharmaceutical interventions on COVID-19 in Europe;584(7820):257-61. Publisher: Nature Research. Available from: https://doi.org/10.1038/s41586-020-2405-7.

[71] Public Health Scotland COVID-19 Statistical Report As at 28 June 2021;. Published: https://www.publichealthscotland.scot/media/8268/21-06-30-covid19-publication_report.pdf.

[72] UK Government. COVID-19 Response - Living with COVID-19;. Available from: https://assets.publishing.service.gov.uk/government/uploads/system/uploads/attachment_data/file/1056229/COVID-19_Response_-_Living_with_COVID-19.pdf.

[73] Andrieu C, Doucet A, Holstein R. Particle Markov Chain Monte Carlo Methods;72:269-342. Available from: https://academic.oup.com/jrsssb/article/72/3/269/7076437.

[74] Andrieu C, Roberts GO. The pseudo-marginal approach for efficient Monte Carlo computations;37. Available from: https://projecteuclid.org/journals/annals-of-statistics/volume-37/issue-2/The-pseudo-marginal-approach-for-efficient-Monte-Carlo-computations/10.1214/07-AOS574.full.

[75] FitzJohn R, Knock E, Whittles L, Perez-Guzman P, Bhatia S, Guntoro F, et al. Reproducible parallel inference and simulation of stochastic state space models using odin, dust, and mcstate [version 2; peer review: 1 approved, 1 approved with reservations];5(288).

[76] R Core Team. R: A Language and Environment for Statistical Computing. R Foundation for Statistical Computing;. Available from: https://www.R-project.org/.

[77] Nyberg T, Ferguson NM, Nash SG, Webster HH, Flaxman S, Andrews N, et al. Comparative analysis of the risks of hospitalisation and death associated with SARS-CoV-2 omicron (B.1.1.529) and delta (B.1.617.2) variants in England: a cohort study;399(10332):1303-12. Available from: https://linkinghub.elsevier.com/retrieve/pii/S0140673622004627.
